# Two-dose SARS-CoV-2 vaccine effectiveness with mixed schedules and extended dosing intervals: test-negative design studies from British Columbia and Quebec, Canada

**DOI:** 10.1101/2021.10.26.21265397

**Authors:** Danuta M Skowronski, Solmaz Setayeshgar, Yossi Febriani, Manale Ouakki, Macy Zou, Denis Talbot, Natalie Prystajecky, John R Tyson, Rodica Gilca, Nicholas Brousseau, Geneviève Deceuninck, Eleni Galanis, Chris D Fjell, Hind Sbihi, Elise Fortin, Sapha Barkati, Chantal Sauvageau, Monika Naus, David M Patrick, Bonnie Henry, Linda M N Hoang, Philippe De Wals, Christophe Garenc, Alex Carignan, Mélanie Drolet, Manish Sadarangani, Marc Brisson, Mel Krajden, Gaston De Serres

**Affiliations:** BC Centre for Disease Control, Communicable Diseases and Immunization Services, Vancouver, British Columbia, Canada; University of British Columbia, School of Population and Public Health, Vancouver, British Columbia, Canada; Centre Hospitalier Universitaire (CHU) de Québec-Université Laval Research Center, Quebec City, Quebec, Canada; Institut national de sante publique du Québec, Biological and Occupational Risks, Quebec City, Quebec, Canada; BC Centre for Disease Control, Data and Analytics Services, Vancouver, British Columbia, Canada; Laval University, Department of Social and Preventive Medicine, Faculty of Medicine, Quebec City, Quebec, Canada; BC Centre for Disease Control, Public Health Laboratory, Vancouver, British Columbia, Canada; University of British, Department of Pathology and Laboratory Medicine, Vancouver, British Columbia, Canada; McGill University, Department of Medicine, Division of Infectious Diseases, McGill University Health Center, Montreal, Quebec, Canada; Office of the Provincial Health Officer, Ministry of Health, Victoria, British Columbia, Canada; Sherbrooke University, Department of Microbiology and Infectious Diseases, Sherbrooke, Quebec, Canada; BC Children’s Hospital Research Institute, Vaccine Evaluation Center, Vancouver, British Columbia, Canada; University of British Columbia, Department of Pediatrics, Vancouver, British Columbia, Canada

## Abstract

**Background:** The Canadian COVID-19 immunization strategy deferred second doses and allowed mixed schedules. We compared two-dose vaccine effectiveness (VE) by vaccine type (mRNA and/or ChAdOx1), interval between doses, and time since second dose in two of Canada’s larger provinces.

**Methods:** Two-dose VE against infections and hospitalizations due to SARS-CoV-2, including variants of concern, was assessed between May 30 and October 2, 2021 using test-negative designs separately conducted among community-dwelling adults ≥18-years-old in British Columbia (BC) and Quebec, Canada.

**Findings:** In both provinces, two doses of homologous or heterologous SARS-CoV-2 vaccines were associated with ∼95% reduction in the risk of hospitalization. VE exceeded 90% against SARS-CoV-2 infection when at least one dose was an mRNA vaccine, but was lower at ∼70% when both doses were ChAdOx1. Estimates were similar by age group (including adults ≥70-years-old) and for Delta-variant outcomes. VE was significantly higher against both infection and hospitalization with longer 7-8-week vs. manufacturer-specified 3-4-week interval between doses. Two-dose mRNA VE was maintained against hospitalization for the 5-7-month monitoring period and while showing some decline against infection, remained ≥80%.

**Interpretation:** Two doses of mRNA and/or ChAdOx1 vaccines gave excellent protection against hospitalization, with no sign of decline by 5-7 months post-vaccination. A 7-8-week interval between doses improved VE and may be optimal in most circumstances. Findings indicate prolonged two-dose protection and support the use of mixed schedules and longer intervals between doses, with global health, equity and access implications in the context of recent third-dose proposals.

## BACKGROUND

In Canada, two mRNA vaccines were approved in December 2020 according to a two-dose schedule with interval between doses of three weeks for BNT162b2 (Pfizer-BioNTech, Comirnaty) and four weeks for mRNA-1273 (Moderna, Spikevax).^1^ On February 26, 2021, a chimpanzee adenoviral vectored (ChAdOx1) vaccine (AstraZeneca, Vaxzevria and equivalent COVISHIELD) was authorized with an interval of 4-12 weeks between doses.^1^ In randomized controlled trials (RCTs), the efficacy of a single dose exceeded 90% for mRNA and 75% for ChAdOx1 vaccines.^2–5^ In early January 2021, confronted with elevated pandemic activity and constrained vaccine supplies, two provinces, British Columbia (BC) and Quebec, opted to extend the interval between doses (to six weeks and 12 weeks, respectively), to provide the benefits of substantial single-dose protection to as many people as possible, as soon as possible.

In early March 2021 (epidemiological week 9), Canada’s National Advisory Committee on Immunization (NACI) endorsed a second dose-deferral approach, recommending that the interval between first and second doses of all SARS-CoV-2 vaccines be extended up to 16 weeks.^6^ On March 29 (week 13), in response to emerging reports of ChAdOx1-associated thrombosis with thrombocytopenia, NACI recommended ChAdOx1 be used for adults ≥55-years-old.^7^ On April 23 (week 16), NACI lowered this age limit to ≥30 years.^7^ After assessing interchangeability, NACI further recommended on June 1 (week 22), that first-dose recipients of ChAdOx1 could be offered either the same product or an mRNA vaccine, and that first-dose mRNA recipients could complete the series with the alternate mRNA vaccine if the same product was not available.^8^

The first SARS-CoV-2 vaccines in BC and Quebec were prioritized for long-term care facility (LTCF) residents and healthcare workers. Vaccination of community-dwelling adults began with the oldest in March 2021, progressing sequentially to younger age groups. With the recommended deferral, second-dose coverage started to increase among elderly adults in May. As vaccine supply improved, the interval between first and second doses was shortened to eight weeks in late-May, by which time (week 21) about 70% of all adults ≥18-years-old in BC and Quebec had received at least one dose and <10% had received two doses. The interval between doses was again shortened in August to four weeks to maximize the number of fully-vaccinated individuals before autumn, with >80% of adults ≥18 years in both provinces having received two doses by early-October 2021.

We report two-dose vaccine effectiveness (VE) against infection and hospitalization, including due to the Delta variant of concern (VOC), among adults ≥18-years-old in BC and Quebec. Adjustments to the COVID-19 vaccination program in response to changing pandemic conditions in BC and Quebec provided further unique opportunity to compare two-dose VE by vaccine type, both homologous and heterologous; by interval between doses; and by time since the second dose.

## METHODS

### Source population

There are ∼4 million adults ≥18-years-old in BC, the westernmost province of Canada, and ∼7 million in Quebec, located in eastern Canada ∼5000 km apart. About half the adult population in BC and Quebec are women with similar age distributions 18–49, 50–69 and ≥70 years: 51% and 49%, 32% and 33%, and 16% and 17%, respectively. A publicly-funded, mostly symptom-based approach for PCR-based SARS-CoV-2 diagnostic testing is broadly accessible in both provinces. Each province experienced a third pandemic wave that peaked in mid-April 2021 (weeks 14–15) then subsided to stable low levels from early-June (weeks 22–23) before gradually increasing again with the start of a fourth pandemic wave in late-July/early-August (weeks 29-31).^9,10^ Whereas during the third wave the Alpha variant predominated in Quebec, and co-dominated with Gamma in BC, during the fourth wave the Delta variant became dominant in both provinces.^9,10^

### Study design

Two-dose VE was estimated by test-negative design, using multivariable logistic regression to derive the adjusted odds ratio (AOR) for vaccination among SARS-CoV-2 test-positive cases versus test-negative controls. VE and 95% confidence intervals (CI) were computed as (1-AOR) x 100%. Unless otherwise specified, all adjusted models included age group (18–49 years, 50–69 years, 70–79 years and ≥80 years), sex (men/women), epidemiological week (categorical) and region. The latter includes the five health authorities in BC,^9^ with the 18 administrative regions of Quebec also regrouped into five categories (Greater Montreal, Greater Quebec City, Central Quebec, Northern Quebec and others).^10^

### Case and control selection

Specimens with collection dates between weeks 22–39 (May 30 to October 2) were eligible. Cases included any RT-PCR-confirmed SARS-CoV-2 infection. Hospitalized cases were admitted on or ≤30 days after specimen collection. Individuals could contribute a single test-positive specimen and were censored from any contribution thereafter. Controls included all specimens that were RT-PCR-negative for SARS-CoV-2 and met inclusion/exclusion criteria.

In variant-specific analyses, cases were categorized as Alpha, Gamma, or Delta VOC. Methods and sampling frame for genetic characterization evolved in response to changing epidemic conditions, case load and laboratory capacity, as described in Supplementary Table 1.

### Vaccine status definition

Clients with record of having received two doses of BNT162b2, mRNA-1273 or ChAdOx1 on or before the specimen collection date were considered vaccinated; those with no record of vaccination prior to specimen collection were considered unvaccinated. Among respiratory specimens collected for SARS-CoV-2 testing that had both dates available, the median interval between illness onset and respiratory specimen collection was two days with interquartile range of 1–4 days in both provinces. We based primary two-dose VE analyses on second-dose receipt ≥14 days before specimen collection, excluding those vaccinated 0-13 days prior.

### Data sources and exclusions

Specimens were sampled from respective provincial databases capturing all RT-PCR testing for SARS-CoV-2 along with client, collection and testing details. Hospitalized cases were identified through linkage with notifiable disease lists, supplemented in Quebec by the administrative hospitalization database. Vaccination information was obtained from provincial immunization registries capturing all SARS-CoV-2 vaccinations along with client and vaccination details. Individual-level database linkages were achieved through unique personal identifiers.

Specimens with invalid or missing information were excluded as were specimens collected from individuals: identified as cases before the analysis period; residents of LTCFs, assisted-living or independent-living facilities; vaccinated with a single dose or product other than BNT162b2, mRNA-1273 or ChAdOx1; or when tested outside of the public-funding scheme owing to systematically lower likelihood of test-positivity.^9^ The latter criterion also excludes individuals routinely screened for travel.

### Ethics statement

Data linkages and analyses were authorized by the Provincial Health Officer (BC) and National Director of Public Health (Quebec) under respective provincial public health legislation without requirement for research ethics board review.

## FINDINGS

### Case and control profiles

In total, 380,532 specimens contributed to VE analyses in BC including 27,439 (7%) test-positive cases of whom 1582 (6%) were hospitalized (Table 1). In Quebec, 854,915 specimens contributed with 17,234 (2%) test-positive cases of whom 878 (5%) were hospitalized. More than 85% of all cases in both provinces accrued during the fourth wave between weeks 31–39. About two-thirds of participating case viruses in each province were genetically-characterized overall, and of those, 91% in BC and 85% in Quebec were the Delta variant (Supplementary Table 1). Cases and controls by age and sex were similar in both provinces. Hospitalized cases were older and more often male (Table 1). Compared to their share of the population, younger adults and females contributed disproportionately to controls.

**Table 1.** Profile of participants ≥18-year-olds, by case and vaccine status (regardless of time since vaccination), British Columbia and Quebec, Canada.

|  | British Columbia |  |  |  |  |  | Quebec |  |  |  |  |  |
| --- | --- | --- | --- | --- | --- | --- | --- | --- | --- | --- | --- | --- |
|  | Overall [N=380532] (column %) |  |  | Vaccinated [N=285102] (row %)<sup>1</sup> |  |  | Overall [N=854915] (column %) |  |  | Vaccinated [N=630631] (row %)<sup>1</sup> |  |  |
|  | Cases | Hosp | Controls | Cases | Hosp | Controls | Cases | Hosp | Controls | Cases | Hosp | Controls |
| **Total** | 27439 (7) | 1582 (6)<sup>2</sup> | 353093 (93) | 8630 (31) | 217 (14) | 276472 (78) | 17234 (2) | 878 (5)<sup>2</sup> | 837681 (98) | 5868 (34) | 176 (20) | 624763 (75) |
| **Age group (years)** |  |  |  |  |  |  |  |  |  |  |  |  |
| 18-49 | 20085 (73) | 588 (37) | 217157 (62) | 5509 (27) | 30 (5) | 159310 (73) | 13070 (76) | 348 (40) | 478750 (57) | 3606 (28) | 31 (9) | 319674 (67) |
| 50-69 | 5792 (21) | 598 (38) | 87688 (25) | 2329 (40) | 75 (13) | 73864 (84) | 3267 (19) | 311 (35) | 232291 (28) | 1622 (50) | 40 (13) | 188911 (81) |
| 70-79 | 1116 (4) | 233 (15) | 29538 (8) | 551 (49) | 47 (20) | 26680 (90) | 610 (4) | 122 (14) | 80884 (10) | 429 (70) | 47 (39) | 74956 (93) |
| 80+ | 446 (2) | 163 (10) | 18710 (5) | 241 (54) | 65 (40) | 16618 (89) | 287 (2) | 97 (11) | 45756 (6) | 211 (74) | 58 (60) | 41222 (90) |
| Median (Interquartile range) | 37 (28-51) | 57 (41-70) | 42 (31-60) | 42 (31-57) | 71 (60-82) | 44 (32-62) | 36 (27-49) | 56 (42-70) | 45 (33-62) | 44 (33-58) | 73 (60-82) | 49 (35-66) |
| **Sex** |  |  |  |  |  |  |  |  |  |  |  |  |
| Female | 13313 (49) | 647 (41) | 198686 (56) | 4595 (35) | 92 (14) | 159390 (80) | 8756 (51) | 372 (42) | 502864 (60) | 3195 (36) | 77 (21) | 369338 (73) |
| Male | 14126 (51) | 935 (59) | 154407 (44) | 4035 (29) | 125 (13) | 117082 (76) | 8478 (49) | 506 (58) | 334817 (40) | 2673 (32) | 99 (20) | 255425 (76) |
| **Epidemiological week** |  |  |  |  |  |  |  |  |  |  |  |  |
| 22-25 (May 30-June 26) | 1493 (5) | 119 (8) | 29093 (8) | 97 (6) | 5 (4) | 11068 (38) | 1227 (7) | 62 (7) | 111687 (13) | 76 (6) | 6 (10) | 46541 (42) |
| 26-30 (June 27-July 31) | 1611 (6) | 84 (5) | 49434 (14) | 346 (21) | 13 (15) | 35026 (71) | 1118 (7) | 50 (6) | 166516 (20) | 234 (21) | 6 (12) | 112998 (68) |
| 31-35 (August 1-Sept 4) | 12588 (46) | 642 (41) | 131403 (37) | 3527 (28) | 75 (12) | 106647 (81) | 6551 (38) | 312 (36) | 250102 (29) | 2037 (31) | 58 (19) | 197160 (79) |
| 36-39 (Sept 5-Oct 2) | 11747 (43) | 737 (47) | 143163 (41) | 4660 (40) | 124 (17) | 123731 (86) | 8338 (48) | 454 (51) | 309376 (37) | 3521 (42) | 106 (23) | 268064 (87) |
| **Vaccinated<sup>3</sup>** |  |  |  |  |  |  |  |  |  |  |  |  |
| Two any mRNA vaccines | 7535 (27)<sup>4</sup> | 198 (13) | 252803 (72)<sup>5</sup> | 7535 (87) | 198 (91) | 252803 (91) | 5379 (31) | 155 (18) | 576827 (69) | 5379 (92) | 155 (88) | 576827 (92) |
| Two BNT162b2 | 5593 (20) | 136 (9) | 184987 (52) | 5593 (65) | 136 (63) | 184987 (67) | 4323 (25) | 123 (14) | 436106 (52) | 4323 (74) | 123 (70) | 436106 (70) |
| Two mRNA-1273 | 1408 (5) | 44 (3) | 46596 (13) | 1408 (16) | 44 (20) | 46596 (17) | 914 (5) | 25 (3) | 123986 (15) | 914 (16) | 25 (14) | 123986 (20) |
| Two mixed mRNA | 533 (2) | 18 (1) | 21208 (6) | 533 (6) | 18 (8) | 21208 (8) | 142 (1) | 7 (1) | 16735 (2) | 142 (2) | 7 (4) | 16735 (3) |
| Two ChAdOx1 | 652 (2) | 17 (1) | 8635 (2) | 652 (8) | 17 (8) | 8635 (3) | 236 (1) | 14 (2) | 18826 (2) | 236 (4) | 14 (8) | 18826 (3) |
| Two mixed ChAdOx1/mRNA | 443 (2) | 2 (<1) | 15034 (4) | 443 (5) | 2 (<1) | 15034 (5) | 253 (2) | 7 (1) | 29110 (3) | 253 (4) | 7 (4) | 29110 (5) |
| **Interval between doses<sup>3</sup>** |  |  |  |  |  |  |  |  |  |  |  |  |
| 21-34 days (3-4 weeks) | NA | NA | NA | 6261 (2) | 277 (3) | 5984 (2) | NA | NA | NA | 430 (7) | 19 (11) | 23300 (4) |
| 35-48 days (5-6 weeks) | NA | NA | NA | 18522 (7) | 782 (9) | 17740 (6) | NA | NA | NA | 484 (8) | 6 (3) | 35880 (6) |
| 49-62 days (7-8 weeks) | NA | NA | NA | 106284 (37) | 3286 (38) | 102998 (37) | NA | NA | NA | 1636 (28) | 15 (9) | 149696 (24) |
| 63-83 days (9-11 weeks) | NA | NA | NA | 103641 (36) | 2802 (32) | 100839 (36) | NA | NA | NA | 1808 (31) | 50 (28) | 204352 (33) |
| 84-111 days (12-15 weeks) | NA | NA | NA | 44228 (16) | 1284 (15) | 42944 (16) | NA | NA | NA | 973 (17) | 52 (30) | 130874 (21) |
| 112+ days (16+ weeks) | NA | NA | NA | 6166 (2) | 199 (2) | 5967 (2) | NA | NA | NA | 537 (9) | 34 (19) | 80661 (13) |
| **Time since second dose<sup>3</sup>** |  |  |  |  |  |  |  |  |  |  |  |  |
| 0-13 days (0-1 weeks) | NA | NA | NA | 986 (11) | 20 (9) | 24123 (9) | NA | NA | NA | 762 (13) | 13 (7) | 69880 (11) |
| 14-27 days (2-3 weeks) | NA | NA | NA | 397 (5) | 9 (4) | 29811 (11) | NA | NA | NA | 335 (6) | 16 (9) | 78087 (12) |
| 28-55 days (4-7 weeks) | NA | NA | NA | 1793 (21) | 30 (14) | 75307 (27) | NA | NA | NA | 1299 (22) | 27 (15) | 173375 (28) |
| 56-83 days (8-11 weeks) | NA | NA | NA | 2685 (31) | 62 (29) | 82806 (30) | NA | NA | NA | 1923 (33) | 46 (26) | 170639 (27) |
| 84-111 days (12-15 weeks) | NA | NA | NA | 1814 (21) | 74 (34) | 45276 (16) | NA | NA | NA | 1121 (19) | 57 (32) | 90232 (14) |
| 112-139 days (16-19 weeks) | NA | NA | NA | 390 (5) | 14 (6) | 9124 (3) | NA | NA | NA | 311 (5) | 13 (7) | 28130 (5) |
| 140-167 days (20-23 weeks) | NA | NA | NA | 89 (1) | 3 (1) | 2385 (1) | NA | NA | NA | 89 (2) | 3 (2) | 12454 (2) |
| 168-195 days (24-27 weeks) | NA | NA | NA | 130 (2) | 0 (0) | 2618 (1) | NA | NA | NA | 19 (<1) | 0 (0) | 1397 (<1) |
| 196+ days (28+ weeks) | NA | NA | NA | 346 (4) | 5 (2) | 5022 (2) | NA | NA | NA | 9 (<1) | 1 (1) | 569 (<1) |
| Median, Interquartile range | NA | NA | NA | 68 (43-90) | 76 (51-95) | 58 (34-82) | NA | NA | NA | 64 (38-85) | 79 (47-96) | 54 (29-79) |
| Range (days) | NA | NA | NA | 0-250 | 2-231 | 0-265 | NA | NA | NA | 0-240 | 1-210 | 0-256 |

NA = Not applicable
<sup>1</sup> Unless otherwise specified, displayed is the percentage of cases, hospitalized cases or controls who received a second vaccine dose by on or before specimen collection, by row category, regardless of time since second dose.
<sup>2</sup> Displayed is the percentage of cases who were hospitalized.
<sup>3</sup> All percentages displayed below this row are column %
<sup>4</sup> One case twice vaccinated with unspecified mRNA vaccines in British Columbia.
<sup>5</sup> Twelve controls twice vaccinated with unspecified mRNA vaccines in British Columbia

### Vaccination profiles

By week 39, >80% of controls in BC and Quebec had been vaccinated with >90% having received two mRNA doses, 3% ChAdOx1 and 5% mixed ChAdOx1/mRNA products (Table 1). Among mixed ChAdOx1/mRNA recipients, >99% in each province had received ChAdOx1 first. About two-thirds of vaccinated controls in BC and Quebec received BNT-162b; 11% and 20% respectively, received mRNA-1273; and 8% and 3%, respectively, received a mix of either mRNA product of whom 80% and 70%, respectively, received BNT162b2 first. Follow-up periods are shown in Table 1 and by vaccine type in Supplementary Tables 2 and 3. ChAdOx1 recipients in both provinces were generally older and with less follow-up time after their second dose.

### Vaccine effectiveness

#### By vaccine type and outcome, including mixed schedules and VOC

Two-dose mRNA VE against infection was 90% (95%CI: 89–90) in BC and 88% (95%CI: 88–89) in Quebec, similar among recipients of the same or mixed mRNA doses (Figure 1, Supplementary Table 4). VE was significantly lower among recipients of two doses of ChAdOx1 at 71% (95%CI: 69–74) in BC and 73% (95%CI: 69–77) in Quebec. However, among those who received a mixed schedule of ChAdOx1 and mRNA vaccination, VE was significantly improved at 90% (95%CI: 89–91) in BC and 87% (95%CI: 85–89) in Quebec.

**Figure 1.**
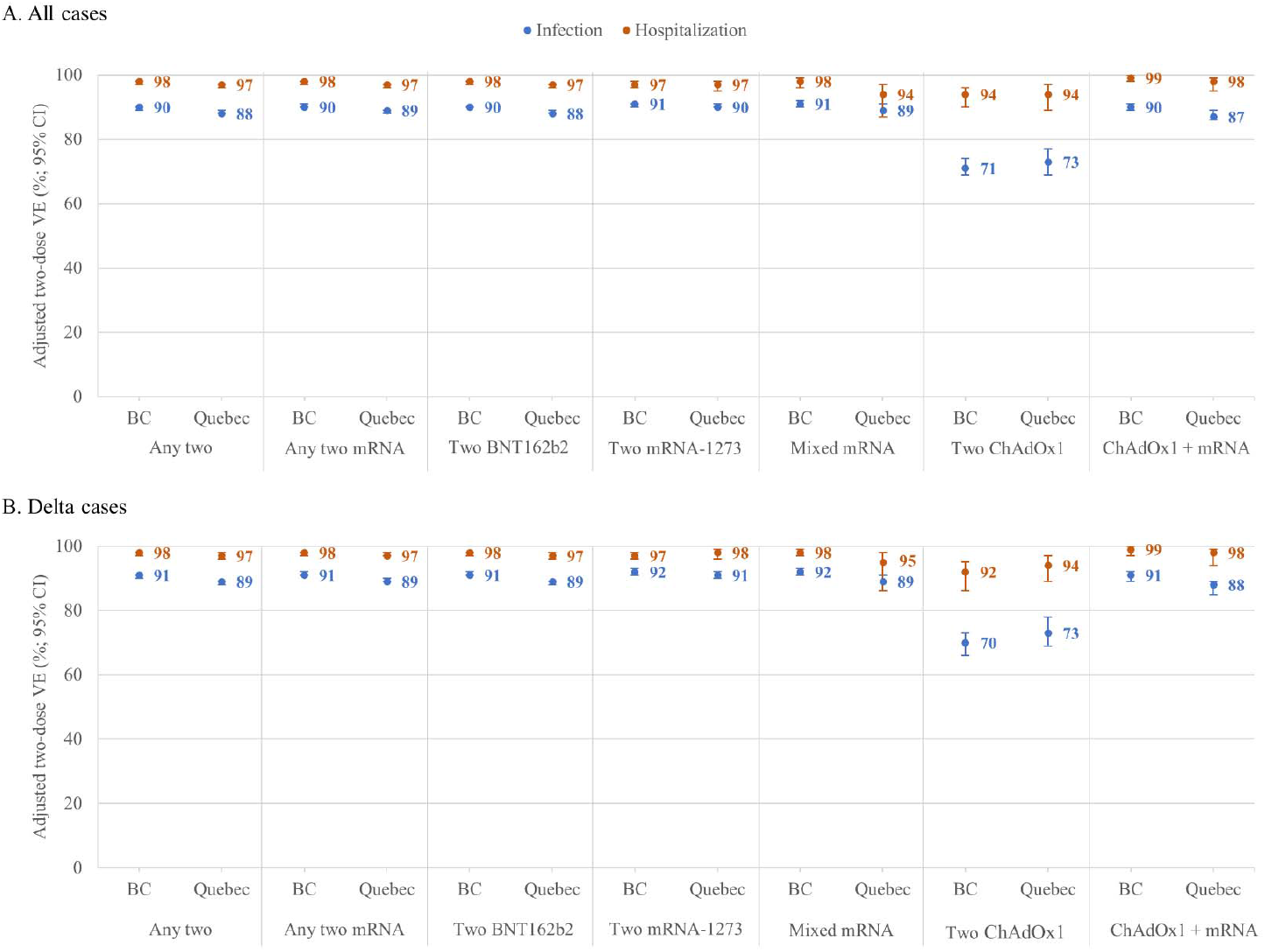
Adjusted two-dose vaccine effectiveness against infection and hospitalization, by vaccine type, ≥18-year-olds, British Columbia and Quebec, Canada. Shown are adjusted vaccine effectiveness (VE) estimates and 95% confidence intervals (CI) against infection (blue) and hospitalization (orange) ≥14 days after the second dose by vaccine type, overall (panel A) and for the Delta variant of concern (panel B) among adults ≥18 years old in the provinces of British Columbia (BC) and Quebec, Canada. All VE estimates are adjusted for age group (18-49, 50-69, 70-79, ≥80 years); sex (men, women); individual epidemiological week of the analysis period (weeks 22-39, categorical); and region of the province (5 in each province). In Quebec, VE against hospitalizations due to the Delta variant was assessed only between weeks 31-39 because no hospitalized Delta variant cases were identified prior to that period. For additional details including corresponding sample sizes and precise unadjusted and adjusted estimates and 95%CI, see Supplementary Table 4 (overall) and Supplementary Tables 8 and 9 (Delta and other variants of concern).

VE of two mRNA doses against hospitalization was 98% (95%CI: 97–98) in BC and 97% (95%CI: 96–97) in Quebec, and similar for two doses of ChAdOx1 at 94% (95%CI: 90–96) and 94% (95%CI: 89–97), respectively (Figure 1). VE against hospitalization was similar among recipients of mixed mRNA or mixed ChAdOx1/mRNA doses (Supplementary Table 4).

VE findings were similar by age group, notably including older adults ≥70 years (Figure 2, Supplementary Tables 5 and 6) and did not meaningfully differ by sex (Figure 2, Supplementary Table 7). VE against the Delta variant was almost identical to the overall analysis (Figure 1), and was similar for other VOC, recognizing smaller sample size (Supplementary Tables 8 and 9).

**Figure 2.**
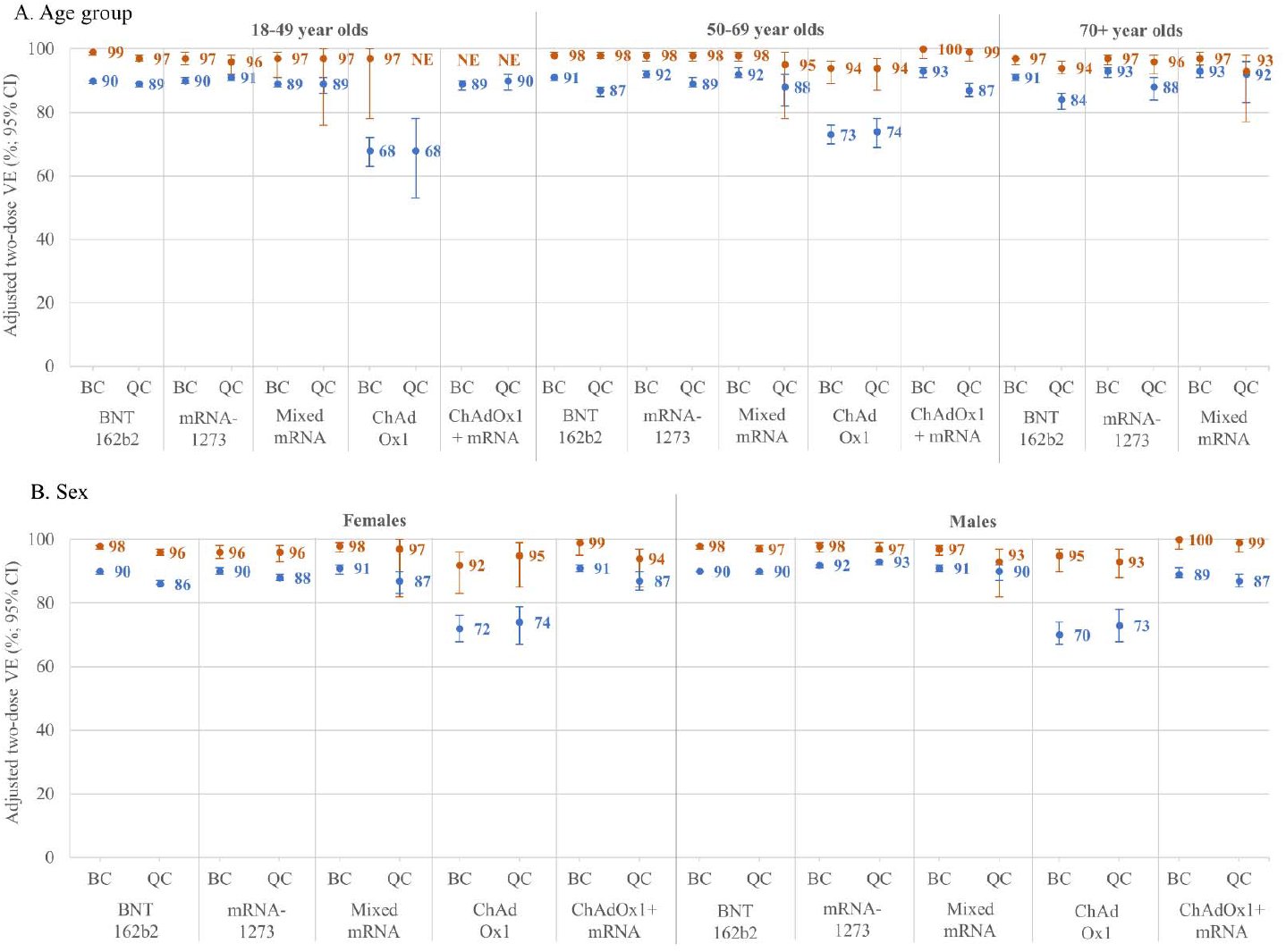
Adjusted two-dose vaccine effectiveness against infection and hospitalization, by age group, sex and vaccine type, ≥18-year-olds, British Columbia and Quebec, Canada. Shown are adjusted vaccine effectiveness (VE) estimates and 95% confidence intervals (CI) against infection (blue) and hospitalization (orange) ≥14 days after the second dose by age group (panel A) and sex (panel B) and by vaccine type in British Columbia (BC) and Quebec (QC), Canada. In panel A, VE estimates are adjusted for sex (men, women); individual epidemiological week of the analysis period (weeks 22-39, categorical); and region of the province (5 in each province). Among those ≥70 years, VE estimates are additionally adjusted for 70-79 and ≥80 years. In panel B, VE estimates are adjusted for the same covariates, omitting sex and including age group (18-49 years, 50-69 years, 70-79 years, ≥80 years). In Quebec, age-specific VE estimates against hospitalization in those 18-49 years of age were adjusted for calendar time bi-weekly and in ≥70-year-olds were adjusted tri-weekly owing to sample size. For additional details including corresponding sample sizes and precise unadjusted and adjusted estimates and 95%CI, see Supplementary Tables 5-7.

#### By time since vaccination

In both provinces, two-dose mRNA VE ≥95% against hospitalization was maintained through the seventh month post-vaccination (Figure 3; Supplementary Table 10). Two-dose mRNA VE against any infections peaked above 90% at 2–3 weeks post-vaccination, but remained about 80% or more through the eighth month. Given greater sample size, findings are most robust for BNT162b2 with similar pattern for mRNA-1273 (Supplementary Table 11) and mixed mRNA or ChAdOx1/mRNA recipients, recognizing limited follow-up beyond the fourth or fifth month (Supplementary Table 12). For homologous two-dose ChAdOx1 recipients, VE ≥70% was also maintained for at least the fourth month post-vaccination (Figure 3; Supplementary Table 10). There was no indication of greater decline in two-dose protection against Delta (Supplementary Tables 13–15). Among adults ≥70-years-old, mRNA VE was ≥80% against infection and ≥90% against hospitalization to at least the fifth month, with smaller sample size thereafter (Figure 4; Supplementary Table 16).

**Figure 3.**
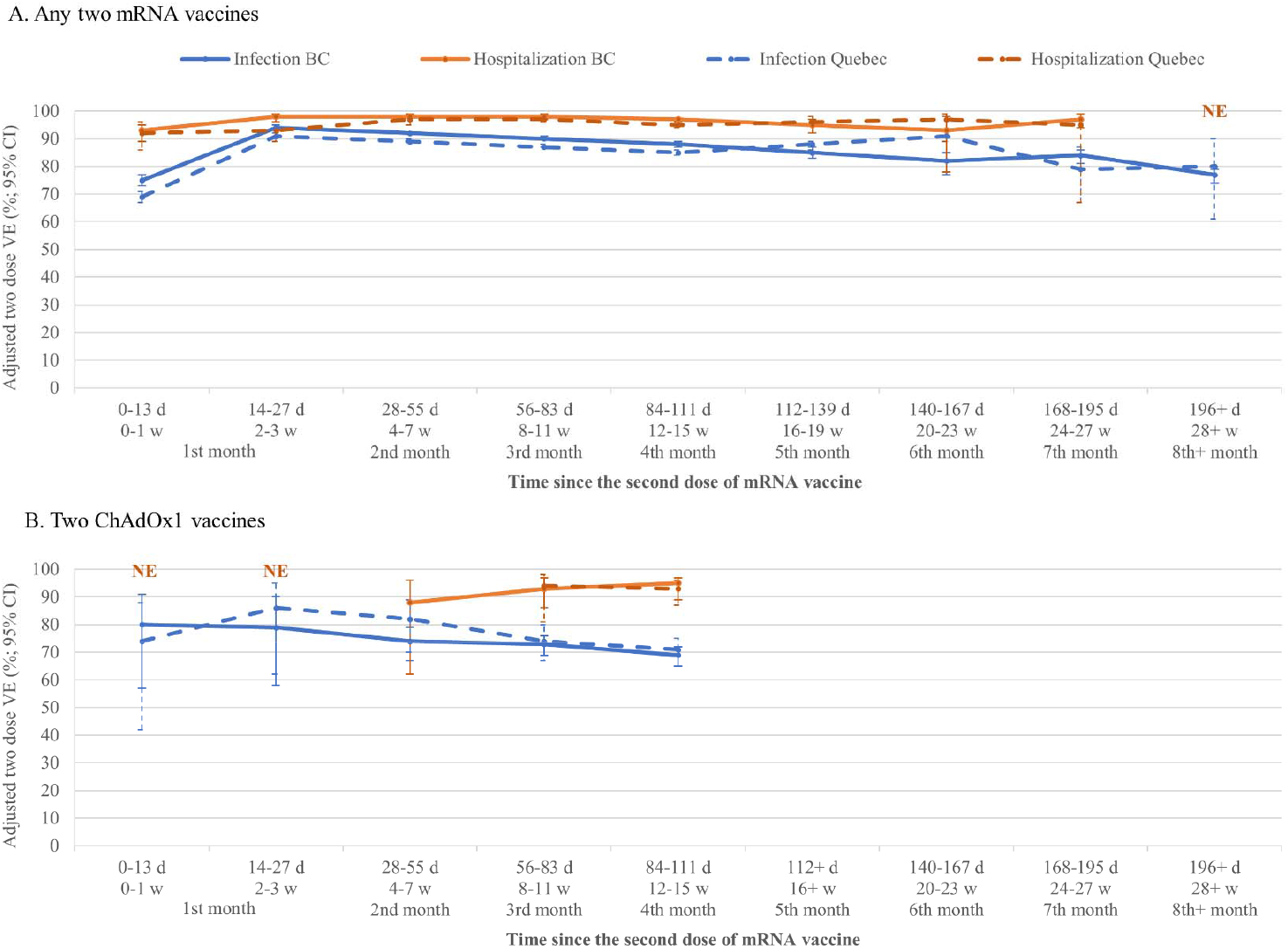
Adjusted two-dose vaccine effectiveness against infection and hospitalization, by time since vaccination, mRNA and ChAdOx1 vaccines, ≥18-year-olds, British Columbia and Quebec, Canada. Shown are adjusted vaccine effectiveness (VE) estimates and 95% confidence intervals (CI) against infection (blue) and hospitalization (orange) by time between receipt of the second dose and specimen collection, among adults ≥18 years old in British Columbia (BC) and Quebec, the latter displayed as dashed lines. Panel A displays estimates for those who received any two mRNA vaccines and panel B displays for those who received two ChAdOx1 vaccines. All VE estimates are adjusted for age group (18-49, 50-69, 70-79, ≥80 years); sex (men, women); individual epidemiological week of the analysis period (weeks 22-39, categorical); and region of the province (5 in each province). The final estimate displayed for mRNA vaccines against hospitalization is for ≥24 weeks since vaccination for both provinces. Estimates could not be displayed beyond 16-19 weeks for ChAdOx1 owing to sparse data. For additional details including corresponding sample sizes and precise unadjusted and adjusted estimates and 95%CI by vaccine type, see **Supplementary Tables 10-12**. The corresponding information is also displayed, Delta-specific, in **Supplementary Tables 13-15**.

**Figure 4.**
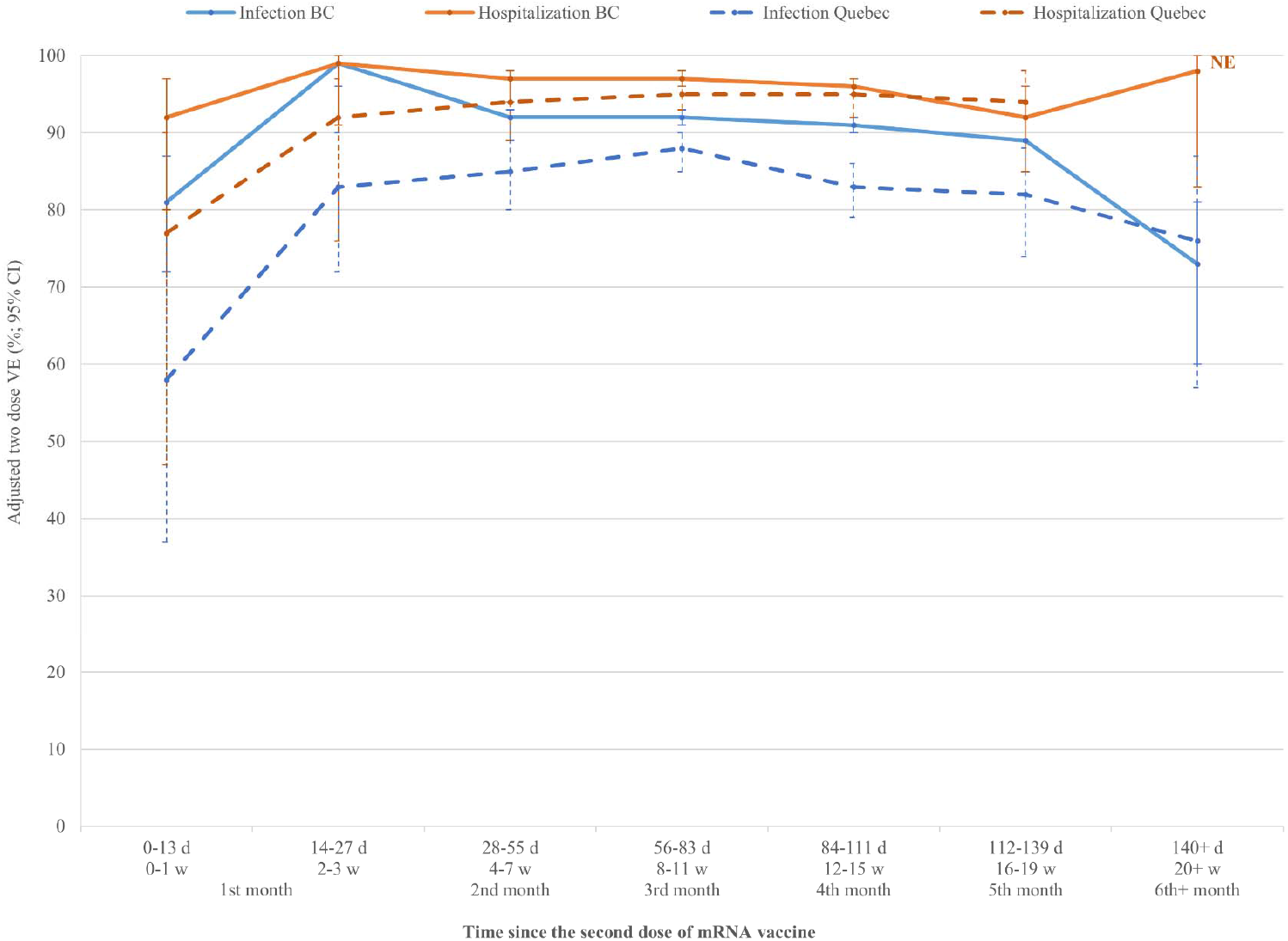
Adjusted two-dose vaccine effectiveness against infection and hospitalization, by time since vaccination, mRNA vaccines, ≥70-year-olds, British Columbia and Quebec, Canada. Shown are adjusted vaccine effectiveness (VE) estimates and 95% confidence intervals (CI) against infection (blue) and hospitalization (orange) by time between receipt of the second dose of any mRNA vaccine and specimen collection, among adults ≥70 years old in British Columbia (BC) and Quebec, the latter displayed as dashed lines. All VE estimates are adjusted for age group (70-79, ≥80 years); sex (men, women); individual epidemiological week of the analysis period (weeks 22-39, categorical); and region of the province (5 in each province) except in Quebec for which calendar time adjustment was tri-weekly for the hospitalization outcome owing to small sample size. For additional details including corresponding sample sizes and precise unadjusted and adjusted estimates and 95%CI, see Supplementary Table 16 (by mRNA vaccine type).

#### By interval between doses

Estimates of mRNA VE against infection improved with a longer interval between first and second doses. With the manufacturer-specified interval of 3–4 weeks, VE in BC was 85% (95%CI: 83–87) and in Quebec was 79% (95%CI: 76–81). At 7-8 weeks VE was significantly higher at 91% (95%CI: 91–91) and 89% (95%CI: 88–89), respectively, and remained relatively stable thereafter (Figure 5, Supplementary Table 17). A similar pattern was observed against hospitalizations with greater VE at 7–8-week interval (99%; 95%CI: 98–99 and 98%; 95%CI: 97– 99, respectively) vs. 3–4-week interval (93%; 95%CI: 87–96 and 87%; 95%CI: 79–92). Given larger sample size, findings are most robust for BNT162b2 (Supplementary Table 18). Confidence intervals were wide for ChAdOx1; in Quebec, but not BC, VE showed gradual increase with longer interval (Figure 5).

**Figure 5.**
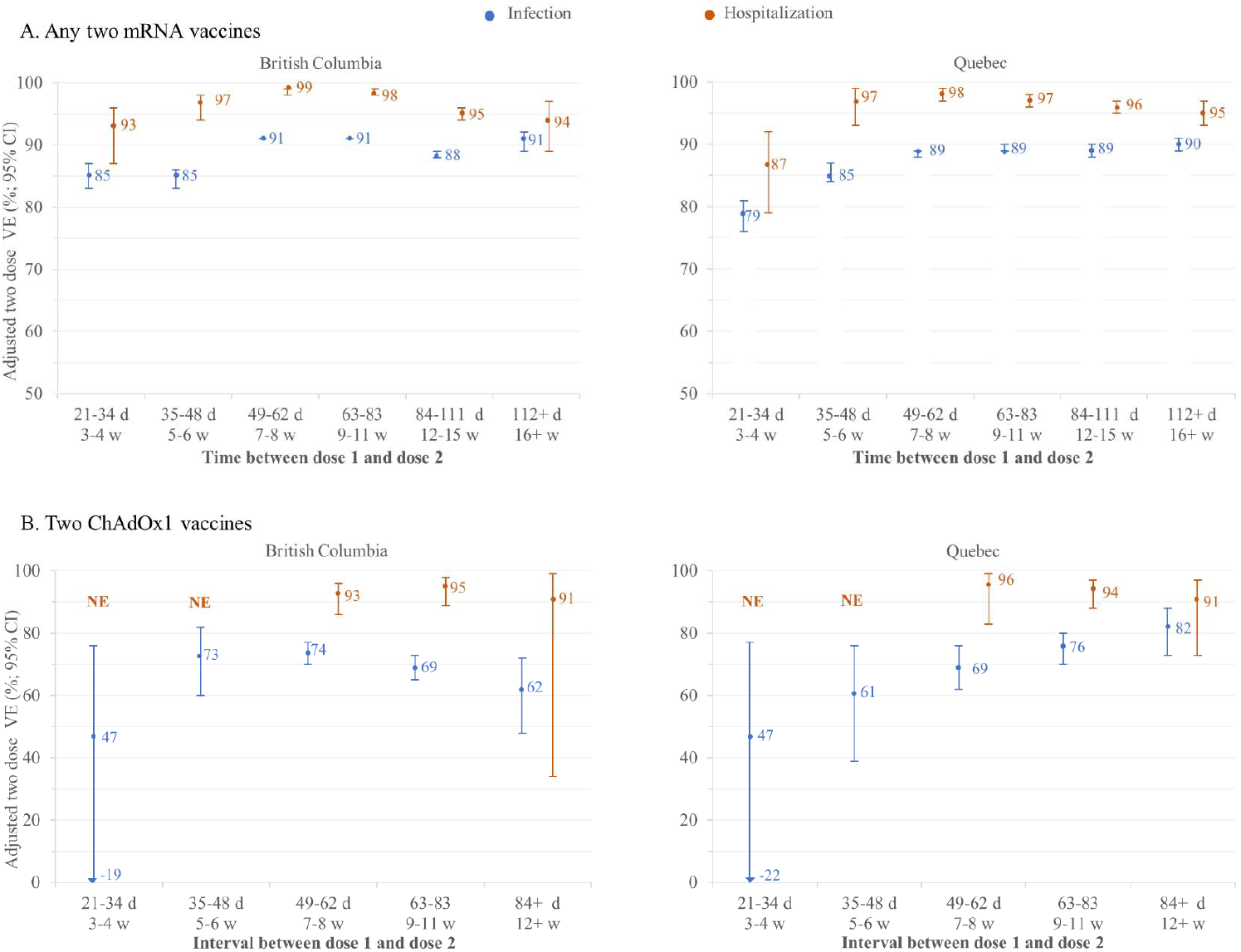
Adjusted two-dose vaccine effectiveness against infection and hospitalization, by interval between doses, mRNA and ChAdOx1 vaccines, ≥18-year-olds, British Columbia and Quebec, Canada. Shown are adjusted vaccine effectiveness (VE) estimates and 95% confidence intervals (CI) against infection (blue) and hospitalization (orange) ≥14 days after the second dose, by interval between the first and second dose among adults ≥18 years old who were vaccinated with any two mRNA vaccines or two ChAdOx1 vaccines in British Columbia (BC) and Quebec. All VE estimates are adjusted for age group (18-49, 50-69, 70-79, ≥80 years); sex (men, women); individual epidemiological week of the analysis period (weeks 22-39, categorical); and region of the province (5 in each province). For additional details including corresponding sample sizes and precise unadjusted and adjusted estimates and 95%CI, see Supplementary Table 17 (by vaccine type) and Supplementary Table 18 (by mRNA vaccine type).

Recognizing that shorter intervals between doses may have been associated with longer time since second dose, we also explored VE stratified simultaneously for period effects. This is shown for BNT162b2 in Figure 6 and for all vaccine types in Supplementary Table 19. The approximate 5-10% higher VE when the second dose was spaced ≥7-versus 3–4 weeks after the first, was maintained at all time points since the second dose. In each province the gap was comparable at 2– 3 weeks and ≥16 weeks post-vaccination. Similar pattern was observed for mRNA-1273 and ChAdOx1 but with greater variability given smaller sample sizes.

**Figure 6.**
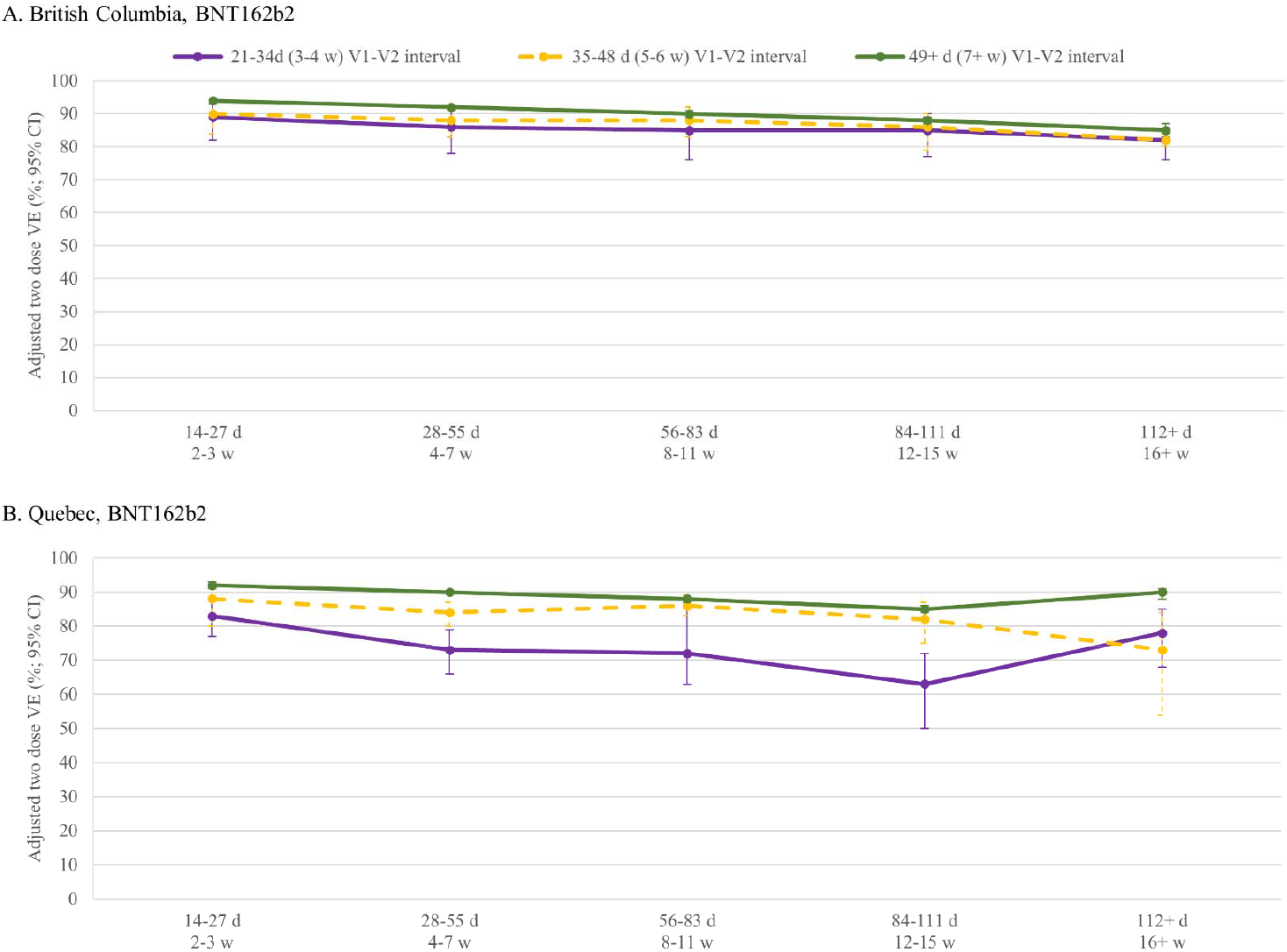
Adjusted two-dose vaccine effectiveness against infection by interval between doses and time since second dose, BNT162b2, ≥18-year-olds, British Columbia and Quebec, Canada. Shown are adjusted vaccine effectiveness (VE) estimates and 95% confidence intervals (CI) against infection (timed on specimen collection date) by interval between the first and second dose (3-4 weeks in purple; 5-6 weeks in dashed yellow; 7+ weeks in green) and time since the second dose among adults ≥18 years old who were vaccinated with two BNT162b2 vaccines in the provinces of British Columbia (BC) and Quebec. All VE estimates are adjusted for age group (18-49, 50-69, 70-79, ≥80 years); sex (men, women); individual epidemiological week of the analysis period (weeks 22-39, categorical); and region of the province (5 in each province). For additional details including corresponding sample sizes and precise unadjusted and adjusted estimates and 95%CI, see Supplementary Table 19 (by vaccine type, including mRNA combined and by mRNA vaccine type, as well as ChAdOx1).

## INTERPRETATION

From two of the larger provinces of Canada, located at nearly opposite ends of the country, we report concordant findings of two-dose SARS-CoV-2 VE against infection and hospitalization, including with mixed vaccines and extended dosing intervals. In both provinces, two homologous or heterologous doses were associated with about 95% reduction in the risk of hospitalization. VE also exceeded 90% against SARS-CoV-2 infection when at least one dose was an mRNA vaccine, but was lower at about 70% when both doses were ChAdOx1. The Delta variant was not associated with compromised VE. Vaccine protection was even better when the spacing between doses was longer than the 3-4 weeks recommended by manufacturers. Over the 5-7-month period after the second dose, mRNA VE was maintained at about 95% against hospitalizations, showing some decline but remaining about 80% or more against infections. With shorter follow-up time for the smaller two-dose ChAdOx1 subset, VE was also maintained to at least the fourth month post-vaccination.

To date there are no head-to-head RCT comparisons of homologous and heterologous SARS-CoV-2 vaccine efficacy. In immunogenicity studies, heterologous ChAdOx1 followed by mRNA vaccination induced antibody titers exceeding homologous vector vaccination and comparable to homologous mRNA vaccination.^11–13^ For pragmatic and immunologic reasons, some countries (e.g. France, Germany, Spain, the United Kingdom (UK)) have, like Canada, recommended and accepted mixed schedules as providing valid two-dose protection. Other countries strictly adhere to the homologous schedules submitted for regulatory approval by sponsor manufacturers. Modification of regulatory approval thereafter typically requires submission of updated data by the sponsor. However, to meet public health needs during a rapidly evolving crisis, decision-makers must be able to respond to emerging post-marketing evidence. In Canada, real-time expert committee recommendation, rather than regulatory review, allowed mixed schedules. By removing the requirement to maintain half of available doses in reserve for homologous series completion, this greatly simplified vaccine deployment and sped vaccine coverage. The findings presented here strongly reinforce vaccine interchangeability to complete the two-dose SARS-CoV-2 vaccine series. Mixed vaccine schedules could reduce logistical hurdles and support nimble vaccination campaigns in countries elsewhere that are still struggling with low vaccine supplies and/or coverage. Global recognition of homologous or heterologous doses as valid proof of vaccination could facilitate a more rapid end to the pandemic and more equitable opportunities for re-opening, travel and return to normal socio-economic interactions, everywhere.

Our estimates of two-dose VE against infection exceeding 90% for mRNA but lower at about 70% for ChAdOx1 vaccine are consistent with respective gold-standard RCT findings,^3–5^ and in particular with maximal ChAdOx1 two-dose efficacy against any infection of 66% in pooled RCT meta-analysis.^5^ In extended follow-up of participants in the pivotal BNT162b2 RCT, two-dose efficacy against clinical infection peaked at 96% during the first two months but remained >80% between four months and the end of follow-up, similar to the sustained protection we report.^14^ In the pivotal mRNA-1273 RCT, efficacy against COVID-19 illness was 93%, without indication of waning across a median of 5·2 months.^15^ Of note, following unblinding of the mRNA-1273 RCT, a two-year open-label study was initiated: participants who had originally received placebo were vaccinated (mRNA-1273p) and compared to participants who were earlier randomized to vaccination (mRNA-1273e).^16^ During the Delta surge in July and August 2021, the rate of COVID-19 and of severe COVID-19 was about 1·6 times greater among the mRNA-1273e group (median 13 months follow-up from the first dose) than the mRNA-1273p group (median follow-up 7·9 months). Applying this relative risk (RR) of 1·6 to the COVID-19 incidence in vaccinated individuals from the original RCT similarly translates into only minor drop in efficacy from 93% in the original trial to 89% with the longer time since vaccination.

Similar to our findings, other jurisdictions have also reported sustained two-dose vaccine protection against hospitalization, including due to the Delta variant.^17–23^ Studies from Israel, however, have reported greater risk of both infection and hospitalization with time since the second BNT162b2 dose.^24^ In the UK, VE against Delta hospitalization was 77% for ChAdOx1 and 93% for BNT162b2 by 20 weeks after the second dose but the decline in VE against symptomatic disease was greater at 47% and 70%, respectively.^21^ Observational studies from California and Qatar have reported even more rapid decline in mRNA VE against infection by five months after the second dose, to just 50% and 22%, respectively.^22,23^ The underlying reasons for these differences in the observed duration of protection are unclear. Despite limitations, surveillance data may provide useful reality check against which to balance some of the more alarming reports of declining VE. For example, the VE estimate of 50% from California corresponds to a RR of two for COVID-19 infection in unvaccinated compared to fully-vaccinated people. However, a RR of two is itself inconsistent with statewide surveillance data instead showing RR exceeding seven between September 26 and October 2, 2021,^25^ crudely corresponding to a VE of 87% five months after most fully-vaccinated Californians had received their second dose. Conversely, RR estimates based upon surveillance data from the UK seem more in line with their VE estimates.^26^ Likewise, in BC and Quebec, during the most recent four-week period available (spanning to early or mid-October, respectively), crude surveillance-based RR estimates of 10 and 7 for infection, respectively, and 52 and 24 for hospitalization, respectively, ^9,27^ correspond to VE estimates >85% against infection and >95% against hospitalization, reassuringly similar to the estimates we report here.

We observed improved VE with interval between first and second doses longer than the 3-4 weeks recommended by manufacturers. Immunogenicity studies have shown that longer intervals are conducive to more complete maturation of the immune response after the first dose, stronger response to the second dose, and ultimately higher and more durable SARS-CoV-2 antibody levels.^28,29^ In the UK, VE was consistently higher with an interval >45 days compared to 19–29 days between BNT162b2 doses, with more erratic findings for ChAdOx1.^28^ In the pooled-analysis of ChAdOx1 RCTS, however, efficacy was much higher with an interval >8 weeks.^5^ Also from the UK, Pouwels et al. found no evidence that BNT162b2 or ChAdOx1 effectiveness varied when comparing dichotomous intervals <9 or ≥9 weeks between doses;^30^ however, their broad categorization <9 weeks may have diluted the lower VE associated with shorter intervals. The optimal interval between doses ultimately represents a balance between rapid and enhanced protection. During a surging pandemic wave, rapid administration of the second dose may prevent some cases that would occur with a longer wait but otherwise, surveillance data in most countries suggest <1% of the unvaccinated population were infected during a given four-week period of the pandemic. With substantial single-dose protection against hospitalization, the absolute risk of severe outcome associated with waiting a few more weeks for the second dose would be small in that context. Conversely, the more durable immunity and approximate 5-10% increment in VE we observed with 7-8-week interval between doses could pay dividends into the future, ultimately preventing more cases and hospitalizations (depending upon evolving incidence and duration of protection). In most instances, therefore, an interval of 7-8 weeks between the first and second dose seems optimal for mRNA vaccines, not only to maximize single-dose coverage in the context of vaccine scarcity but also to optimize the second-dose booster response.

This study, based upon general laboratory submissions, surveillance registries and administrative data, has limitations. In particular, such data are subject to missing or incomplete information, misclassification and selection bias. The test-negative design partially standardizes for healthcare seeking behaviours, but testing indications for SARS-CoV-2 are broad and discretionary, and case ascertainment will have varied between population sub-groups and over time. With increasing vaccine coverage, the subset of remaining unvaccinated individuals may be less comparable in their likelihood of testing positive, with the direction of such bias unknown, and likely to vary with other public health measures. VE estimates were adjusted for calendar time (and age, sex, region of residence) but information pertaining to other potential confounders such as co-morbidity and socio-economic status were not readily available. We cannot rule out residual confounding. Higher-risk healthcare workers or immunocompromised individuals targeted for more rapid second-dose receipt may have contributed to the lower VE associated with shorter interval between doses; however, weighted by their small percentage of the population, such under-estimation would be minor. Further reduced sample size with additional stratification affects the precision of VE estimates, requiring cautious interpretation. Finally, to address differential likelihood of vaccination and exposure risk, our studies were conducted in community-dwelling adults; results may not be generalizable to residents of LTCFs and nursing homes.

In conclusion, two doses of mRNA and/or ChAdOx1 vaccines provided powerful and persistent protection against hospitalization, including due to the Delta variant, without sign of decline by 5-7 months post-vaccination among community-dwelling adults, including older adults. VE against infection declined from an earlier post-vaccination peak above 90% but still prevented 80% or more of infections by the seventh month post-vaccination. Extending the interval between first and second doses may have optimized booster dose protection in Canada. Given these findings, the need and timing of a third dose warrant serious reflection by decision-makers, especially since two-dose, or even one-dose, coverages still remain low in many areas of the world. Our findings support mixed SARS-CoV-2 vaccine schedules and extended intervals between doses, each of which may improve vaccine coverage and have health, equity and access implications globally.

## Data Availability

Sharing of de-identified data that underpin the vaccine effectiveness results reported in this article will be considered upon request with appropriate review and aggregation as required to comply with respective provincial/national privacy and confidentiality legislation and through secure file transfer protocols. Information regarding submitting proposals and accessing data may be obtained through the corresponding authors.

## Acknowledgments

We thank Shinhye Kim at the BC Centre for Disease Control for support in manuscript assembly and the summary tabulation of findings. Manish Sadarangani acknowledges general salary support provided to him by awards from the BC Children’s Hospital Foundation, the Canadian Child Health Clinician Scientist Program and the Michael Smith Foundation for Health Research. Denis Talbot was recipient of a Career Award from the Fond de recherche du Québec–Santé. Finally, we thank the many frontline, regional and provincial practitioners, including clinical, laboratory and public health providers, epidemiologists, Medical Health Officers, laboratory staff, vaccinators, participants and others who contributed to the epidemiological, virological and genetic characterization data underpinning these analyses.

