## Supplementary Material for "Two-dose SARS-CoV-2 vaccine effectiveness with mixed schedules and extended dosing intervals: test-negative design studies from British Columbia and Quebec, Canada"

**Supplementary Table 1.** Variant of concern detection overall and by epidemiological week..... 2

**Supplementary Table 2.** Profile of participants by vaccine type, British Columbia.................... 3

**Supplementary Table 3.** Profile of participants by vaccine type, Quebec .................................. 5

**Supplementary Table 4.** VE by vaccine type ............................................................................. 7

**Supplementary Table 5.** VE against infection, by age group and vaccine type........................... 8

**Supplementary Table 6.** VE against hospitalization, by age group and vaccine type................. 9

**Supplementary Table 7.** VE by sex and vaccine type ............................................................... 10

**Supplementary Table 8.** VE against Alpha and Delta variants, by vaccine type....................... 11

**Supplementary Table 9.** VE against Gamma variant, British Columbia, by vaccine type ........ 12

**Supplementary Table 10.** VE by time since vaccination (mRNA and ChAdOx1).................... 13

**Supplementary Table 11.** VE by time since vaccination (and mRNA vaccine type) ................ 14

**Supplementary Table 12.** VE by time since vaccination (mixed mRNA and mRNA/ChAdOx1) ....................................................................................................................................... 15

**Supplementary Table 13.** VE against Delta, by time since vaccination (mRNA and ChAdOx1) ....................................................................................................................................... 16

**Supplementary Table 14.** VE against Delta, by time since vaccination (and mRNA vaccine type) ............................................................................................................................... 17

**Supplementary Table 15.** VE against Delta, by time since vaccination (mixed mRNA and mRNA/ChAdOx1) ........................................................................................................... 18

**Supplementary Table 16**. VE by time since vaccination, (and mRNA vaccine type), ≥70 years ....................................................................................................................................... 19

**Supplementary Table 17.** VE by interval between doses (mRNA and ChAdOx1) .................. 21

**Supplementary Table 18.** VE by interval between doses (and mRNA vaccine type)................ 22

**Supplementary Table 19.** VE by interval between doses and time since second dose, by vaccine type............................................................................................................................. 23

**Supplementary Table 1. Variant of concern detection overall and by epidemiological week, ≥18-year-old study participants, British Columbia and Quebec, Canada**

In variant-specific analyses, cases were categorized as Alpha, Gamma, or Delta variants of concern (VOC)[1,2]. In both provinces, the methods and sampling frame for genetic characterization of viruses evolved in response to changing epidemic patterns, case load and laboratory capacity. In British Columbia (BC), genetic characterization of all case viruses was attempted by whole genome sequencing (WGS) between weeks 22 and 35 as previously described,[3,4,5,6] but with WGS applied to a subset only (10% random sample in addition to all hospitalized, vaccinated or outbreak-associated cases) between weeks 36-39 when the Delta variant almost solely contributed. In Québec,[7] between week 22 and week 35, diagnosis of Alpha was foremost by RT-PCR single-nucleotide polymorphism (SNP) screen for signature mutations, with a subset identified by WGS. After week 35, most Alpha diagnoses were by WGS. Detection of Delta by RT-PCR SNP screen was confirmed by WGS until week 24 and thereafter foremost by RT-PCR screen alone with a subset identified by WGS. The Gamma variant was detected in Quebec by WGS.

As shown in the table below, in each province about two-thirds of case viruses overall in the current study were genetically characterized. In BC, 91% of all participant case viruses were genetically characterized between weeks 22-35 with Alpha and Gamma variants comprising an approximately equal share (49% and 43%, respectively) between weeks 22-25, but <25% combined between weeks 26-30 and <1% between weeks 31-35 when Delta represented 77% and 99% of all characterized viruses, respectively. Although just 30% of participant case viruses were characterized in BC between weeks 36-39, Delta comprised virtually all of these. In Quebec, the Alpha variant predominated over Delta in weeks 22-25 (97%) and 26-30 (77%) but with Delta predominating over Alpha from August to October, including weeks 31-35 (89%) and 36-39 (99%). Quebec did not detect any Gamma or Beta variants and BC detected a single Beta variant [not displayed below] among participant cases during the analysis period.

| | British Columbia (BC) | | | | | | Quebec | | | | | |
|---|---|---|---|---|---|---|---|---|---|---|---|---|
| | **Total case viruses (N, column %)[8]** | **Genetically characterized case viruses (N, row %)[9]** | **Among genetically characterized viruses** | | | | **Total case viruses (N, column %)[8]** | **Genetically characterized case viruses (N, row %)[9]** | **Among genetically characterized viruses** | | | |
| | | | **Delta (n, row %)[10]** | **Alpha (n, row %)[10]** | **Gamma (n, row %)[10]** | **Non-VOC (n, row %)[10]** | | | **Delta (n, row %)[10]** | **Alpha (n, row %)[10]** | **Gamma (n, row %)[10]** | **Non-VOC (n, row %)[10]** |
| **Overall** | 27439 | 17690 (64%) | 16089 (91%) | 772 (4%) | 787 (4%) | 41 (<1%) | 17234 | 11830 (67%) | 10095 (85%) | 1735 (15%) | Not available | Not available |
| **By epidemiological week** | | | | | | | | | | | | |
| 22-25 (May 30-June 26) | 1493 (5) | 1245 (83%) | 88 (7) | 609 (49) | 532 (43) | 15 (1) | 1227 (7) | 819 (67) | 26 (3) | 793 (97) | NA | NA |
| 26-30 (June 27-July 31) | 1611 (6) | 1452 (90%) | 1114 (77) | 112 (8) | 213 (15) | 13 (1) | 1118 (6) | 589 (53) | 137 (23) | 452 (77) | NA | NA |
| 31-35 (August 1-Sept 4) | 12588 (46) | 11381 (90%) | 11278 (99) | 50 (<1) | 40 (<1) | 13 (<1) | 6551 (38) | 4065 (62) | 3609 (89) | 456 (11) | NA | NA |
| 36-39 (Sept 5-Oct 2) | 11747 (43) | 3612 (31%) | 3609 (>99) | 1 (<1) | 2 (<1) | 0 | 8338 (48) | 6357 (76) | 6323 (99) | 34 (1) | NA | NA |

NA=Not available

[1] World Health Organization. Tracking SARS-CoV-2 variants. [Accessed 16 October 2021]. Available: https://www.who.int/en/activities/tracking-SARS-CoV-2-variants/
[2] Government of Canada. SARS-CoV-2 variants: National definitions, classifications and public health actions. [Accessed 16 October 2021]. Available: https://www.canada.ca/en/public-health/services/diseases/2019-novel-coronavirus-infection/health-professionals/testing-diagnosing-case-reporting/sars-cov-2-variants-national-definitions-classifications-public-health-actions.html
[3] Skowronski DM, Setayeshgar S, Zou M, et al. Single-dose mRNA vaccine effectiveness against SARS-CoV-2, including Alpha and Gamma variants: a test-negative design in adults 70 years and older in British Columbia, Canada. Clin Infect Dis. 2021 Jul 9:ciab616. Doi: 10.1093/ciab616. Online ahead of print.
[4] Skowronski DM, Setayeshgar S, Zou M, et al. Comparative single-dose mRNA and ChAdOx1 vaccine effectiveness against SARS-CoV-2, including early variants of concern: a test-negative design, British Columbia, Canada. [Accessed 16 October 2021]. Available: https://www.medrxiv.org/content/10.1101/2021.09.20.21263875v1
[5] Hogan CA, Jassem AN, Sbihi H, et al. Rapid increase in SARS-CoV_2 P.1 lineage leading to codominance with B.1.1.7 lineage, British Columbia, Canada, January-April 2021. Emerging Infectious Diseases 2021;27: [early release]. Doi: 10.3201/eid2711.211190. Available: https://wwwnc.cdc.gov/eid/article/27/11/21-1190_article
[6] British Columbia Centre for Disease Control (BCCDC). COVID-19 VoC report. [Accessed 16 October 2021]. Available from: http://www.bccdc.ca/health-info/diseases-conditions/covid-19/data#variants
[7] Données sur les variants du SRAS-CoV-2 au Québec. Quebec City: INSPQ. [Accessed 16 October 2021]. Available from: https://www.inspq.qc.ca/covid-19/donnees/variants
[8] Percentage of cases viruses by epidemiological period (column %)
[9] Percentage of total case viruses genetically characterized overall and each epidemiological period (row %).
[10] Percentage of genetically characterized viruses that were the specified variant of concern (VOC) overall and by epidemiological period (row %).

**Supplementary Table 2. Profile of participants ≥18-year-olds, by vaccine type (regardless of time since vaccination), British Columbia, Canada**

| | Any two mRNA | | | BNT162b2 (Pfizer-BioNTech, Comirnaty) | | | mRNA-1273 (Moderna, Spikevax) | | | ChAdOx1 (AstraZeneca, Vaxzevria/COVISHIELD) | | |
|---|---|---|---|---|---|---|---|---|---|---|---|---|
| | **Cases (n, %)** | **Hosp (n, %)** | **Controls (n, %)** | **Cases (n, %)** | **Hosp (n, %)** | **Controls (n, %)** | **Cases (n, %)** | **Hosp (n, %)** | **Controls (n, %)** | **Cases (n, %)** | **Hosp (n, %)** | **Controls (n, %)** |
| **Total N** | 7535 | 198 | 252803 | 5593 | 136 | 184987 | 1408 | 44 | 46596 | 652 | 17 | 8635 |
| **Age group (years)** | | | | | | | | | | | | |
| 18-49 | 4958 (66) | 29 (15) | 148071 (59) | 3700 (66) | 16 (12) | 110302 (60) | 967 (69) | 10 (23) | 28821 (62) | 252 (39) | 1 (6) | 2794 (32) |
| 50-69 | 1795 (24) | 60 (30) | 61685 (24) | 1295 (23) | 37 (27) | 42794 (23) | 328 (23) | 17 (39) | 11716 (25) | 393 (60) | 14 (82) | 5704 (66) |
| 70-79 | 543 (7) | 44 (22) | 26476 (10) | 399 (7) | 34 (25) | 19451 (11) | 89 (6) | 5 (11) | 3774 (8) | 6 (1) | 2 (12) | 109 (1) |
| 80+ | 239 (3) | 65 (33) | 16571 (7) | 199 (4) | 49 (36) | 12440 (7) | 24 (2) | 12 (27) | 2285 (5) | 1 (0) | 0 (0) | 28 (0) |
| Median (Interquartile range) | 41 (30-57) | 72.5 (62-82) | 43 (31-63) | 40 (29-56) | 74 (64-84) | 43 (31-63) | 40 (30-54) | 65 (52-81) | 42 (31-59) | 54 (43-59) | 60 (55-63) | 57 (45-61) |
| **Sex** | | | | | | | | | | | | |
| Female | 4143 (55) | 84 (42) | 147458 (58) | 3075 (55) | 55 (40) | 108987 (59) | 772 (55) | 23 (52) | 26244 (56) | 261 (40) | 7 (41) | 3974 (46) |
| Male | 3392 (45) | 114 (58) | 105345 (42) | 2518 (45) | 81 (60) | 76000 (41) | 636 (45) | 21 (48) | 20352 (44) | 391 (60) | 10 (59) | 4661 (54) |
| **Epidemiological week** | | | | | | | | | | | | |
| 22-25 (May 30-June 26) | 75 (1) | 5 (3) | 10331 (4) | 65 (1) | 5 (4) | 8465 (5) | 9 (1) | 0 (0) | 1318 (3) | 3 (0) | 0 (0) | 452 (5) |
| 26-30 (June 27-July 31) | 303 (4) | 12 (6) | 31733 (13) | 256 (5) | 12 (9) | 23013 (12) | 28 (2) | 0 (0) | 4969 (11) | 28 (4) | 1 (6) | 1384 (16) |
| 31-35 (August 1-Sept 4) | 3064 (41) | 64 (32) | 97628 (39) | 2322 (42) | 39 (29) | 71807 (39) | 538 (38) | 17 (39) | 17827 (38) | 292 (45) | 9 (53) | 3209 (37) |
| 36-39 (Sept 5-Oct 2) | 4093 (54) | 117 (59) | 113111 (45) | 2950 (53) | 80 (59) | 81702 (44) | 833 (59) | 27 (61) | 22482 (48) | 329 (50) | 7 (41) | 3590 (42) |
| **Interval between doses** | | | | | | | | | | | | |
| 21-34 days (3-4 weeks) | 269 (4) | 12 (6) | 5907 (2) | 173 (3) | 6 (4) | 3642 (2) | 82 (6) | 5 (11) | 2027 (4) | 7 (1) | 0 | 50 (<1) |
| 35-48 days (5-6 weeks) | 748 (10) | 12 (6) | 17316 (7) | 494 (9) | 4 (3) | 11820 (6) | 223 (16) | 8 (18) | 4606 (10) | 25 (4) | 0 | 303 (4) |
| 49-62 days (7-8 weeks) | 2858 (38) | 29 (15) | 94445 (37) | 2184 (39) | 21 (15) | 68418 (37) | 447 (32) | 4 (9) | 18111 (39) | 309 (47) | 10 (59) | 4442 (51) |
| 63-83 days (9-11 weeks) | 2309 (31) | 63 (32) | 89169 (35) | 1732 (31) | 46 (34) | 65402 (35) | 391 (28) | 10 (23) | 15485 (33) | 265 (41) | 6 (35) | 3372 (39) |
| 84-111 days (12-15 weeks) | 1166 (15) | 68 (34) | 40215 (16) | 869(16) | 49 (36) | 31225 (17) | 234 (17) | 14 (32) | 5505 (12) | 45 (7) | 1 (6) | 444 (5) |
| 112+ days (16+ weeks) | 184 (2) | 14 (7) | 5739 (2) | 141(3) | 10 (7) | 4480 (2) | 31 (2) | 3 (7) | 862 (2) | 1 (<1) | 0 | 24 (<1) |
| **Time since second dose** | | | | | | | | | | | | |
| 0-13 d (0-1 w) | 940 (12) | 20 (10) | 22615 (9) | 717 (13) | 14 (10) | 16156 (9) | 199 (14) | 6 (14) | 4707 (10) | 7 (1) | 0 (0) | 558 (6) |
| 14-27 d (2-3 w) | 378 (5) | 8 (4) | 28148 (11) | 302 (5) | 6 (4) | 20709 (11) | 65 (5) | 1 (2) | 5512 (12) | 8 (1) | 0 (0) | 548 (6) |
| 28-55 d (4-7 w) | 1614 (21) | 26 (13) | 70047 (28) | 1268 (23) | 18 (13) | 51929 (28) | 248 (18) | 4 (9) | 13052 (28) | 77 (12) | 3 (18) | 1504 (17) |
| 56-83 d (8-11 w) | 2261 (30) | 55 (28) | 74147 (29) | 1667 (30) | 40 (29) | 53607 (29) | 356 (25) | 9 (20) | 12943 (28) | 248 (38) | 7 (41) | 2826 (33) |
| 84-111 d (12-15 w) | 1428 (19) | 67 (34) | 38993 (15) | 1009 (18) | 47 (35) | 27629 (15) | 261 (19) | 13 (30) | 6685 (14) | 272 (42) | 7 (41) | 2948 (34) |
| 112-139 d (16-19 w) | 350 (5) | 14 (7) | 8837 (4) | 234 (4) | 6 (4) | 6906 (4) | 112 (8) | 8 (18) | 1749 (4) | 39 (6) | 0 (0) | 242 (3) |
| 140-167 d (20-23 w) | 88 (1) | 3 (2) | 2376 (1) | 62 (1) | 2 (1) | 1811 (1) | 25 (2) | 1 (2) | 558 (1) | 1 (0) | 0 (0) | 9 (0) |
| 168-195 d (24-27 w) | 130 (2) | 0 (0) | 2618 (1) | 104 (2) | 0 (0) | 2069 (1) | 26 (2) | 0 (0) | 547 (1) | 0 (0) | 0 (0) | 0 (0) |
| 196+ d (28+ w) | 346 (5) | 5 (3) | 5022 (2) | 230 (4) | 3 (2) | 4171 (2) | 116 (8) | 2 (5) | 843 (2) | 0 (0) | 0 (0) | 0 (0) |
| Median | 66 | 76 | 57 | 64 | 74 | 57 | 71 | 90 | 56 | 82 | 77 | 74 |
| Range (days) | 0-252 | 2-231 | 0-265 | 0-252 | 2-214 | 0-265 | 0-247 | 2-231 | 0-253 | 0-144 | 41-99 | 0-157 |
| Interquartile range (days) | 40-90 | 51-99 | 33-81 | 39-88 | 51-95 | 33-81 | 40-101 | 55.5-111 | 31-80 | 66-98 | 67-90 | 49-93 |

Hosp = Hospitalized cases

**Supplementary Table 2 Cont'd. Profile of participants ≥18-year-olds, by vaccine type (regardless of time since vaccination), British Columbia, Canada**

| | **Mixed mRNA** | | | **Mixed ChAdOX1 plus mRNA** | | |
|---|---|---|---|---|---|---|
| | **Cases (n, %)** | **Hosp (n, %)** | **Controls (n, %)** | **Cases (n, %)** | **Hosp (n, %)** | **Controls (n, %)** |
| **Total N** | 533 | 18 | 21208 | 443 | 2 | 15034 |
| **Age group (years)** | | | | | | |
| 18-49 | 291 (55) | 3 (17) | 8941 (42) | 299 (67) | 0 (0) | 8445 (56) |
| 50-69 | 171 (32) | 6 (33) | 7173 (34) | 141 (32) | 1 (50) | 6475 (43) |
| 70-79 | 55 (10) | 5 (28) | 3248 (15) | 2 (0) | 1 (50) | 95 (1) |
| 80+ | 16 (3) | 4 (22) | 1846 (9) | 1 (0) | 0 (0) | 19 (0) |
| Median (Interquartile range) | 46 (33-62) | 69.5 (64-75) | 56 (37-69) | 44 (37-52) | 63 (53-73) | 47 (41-57) |
| **Sex** | | | | | | |
| Female | 295 (55) | 6 (33) | 12221 (58) | 191 (43) | 1 (50) | 7958 (53) |
| Male | 238 (45) | 12 (67) | 8987 (42) | 252 (57) | 1 (50) | 7076 (47) |
| **Epidemiological week** | | | | | | |
| 22-25 (May 30-June 26) | 1 (<1) | 0 | 548 (3) | 19 (4) | 0 (0) | 285 (2) |
| 26-30 (June 27-July 31) | 19 (4) | 0 | 3748 (18) | 15 (3) | 0 (0) | 1909 (13) |
| 31-35 (August 1-Sept 4) | 204 (38) | 8 (44) | 7990 (38) | 171 (39) | 2 (100) | 5810 (39) |
| 36-39 (Sept 5-Oct 2) | 309 (58) | 10 (55) | 8922 (42) | 238 (54) | 0 (0) | 7030 (47) |
| **Interval between doses** | | | | | | |
| 21-34 days (3-4 weeks) | 14 (3) | 1 (6) | 238 (1) | 1 (<1) | 0 | 25 (<1) |
| 35-48 days (5-6 weeks) | 31 (6) | 0 | 890 (4) | 9 (2) | 0 | 120 (<1) |
| 49-62 days (7-8 weeks) | 227 (43) | 4 (22) | 7916 (37) | 119 (27) | 0 | 4107 (27) |
| 63-83 days (9-11 weeks) | 186 (35) | 7 (39) | 8282 (39) | 227 (51) | 2 (100) | 8295 (55) |
| 84-111 days (12-15 weeks) | 63 (12) | 5 (28) | 3485 (16) | 73 (16) | 0 | 2285 (15) |
| 112+ days (16+ weeks) | 12 (2) | 1 (6) | 397 (2) | 14 (3) | 0 | 202 (1) |
| **Time since second dose** | | | 16947 (80) | | | |
| 0-13 d (0-1 w) | 24 (5) | 0 | 1750 (8) | 39 (9) | 0 (0) | 950 (6) |
| 14-27 d (2-3 w) | 11 (2) | 1 | 1925 (9) | 11 (2) | 1 (50) | 1115 (7) |
| 28-55 d (4-7 w) | 98 (18) | 4 | 5064 (24) | 102 (23) | 1 (50) | 3756 (25) |
| 56-83 d (8-11 w) | 238 (45) | 6 | 7594 (36) | 176 (40) | 0 (0) | 5833 (39) |
| 84-111 d (12-15 w) | 157 (29) | 7 | 4677 (22) | 114 (26) | 0 (0) | 3335 (22) |
| 112-139 d (16-19 w) | 4 (1) | 0 | 182 (1) | 1 (0) | 0 (0) | 45 (0) |
| 140-167 d (20-23 w) | 1 (<1) | 0 | 7 (<1) | 0 (0) | 0 (0) | 0 (0) |
| 168-195 d (24-27 w) | 0 | 0 | 1 (<1) | 0 (0) | 0 (0) | 0 (0) |
| 196+ d (28+ w) | 0 | 0 | 8 (<1) | 0 (0) | 0 (0) | 0 (0) |
| Median | 71 | 76 | 63 | 68 | 34.5 | 64 |
| Range (days) | 1-144 | 23-104 | 0-224 | 0-118 | 27-42 | 0-134 |
| Interquartile range (days) | 56-86 | 52-93 | 38-82 | 46-84 | 27-42 | 43-82 |

Hosp = Hospitalized cases

**Supplementary Table 3. Profile of participants ≥18-year-olds, by vaccine type (regardless of time since vaccination), Quebec, Canada**

| | Any two mRNA | | | BNT162b2 (Pfizer-BioNTech, Comirnaty) | | | mRNA-1273 (Moderna, Spikevax) | | | ChAdOx1 (AstraZeneca, Vaxzevria/COVISHIELD) | | |
|---|---|---|---|---|---|---|---|---|---|---|---|---|
| | Cases (n, %) | Hosp (n, %) | Controls (n, %) | Cases (n, %) | Hosp (n, %) | Controls (n, %) | Cases (n, %) | Hosp (n, %) | Controls (n, %) | Cases (n, %) | Hosp (n, %) | Controls (n, %) |
| **Total N** | 5379 | 155 | 576827 | 4323 | 123 | 436106 | 914 | 25 | 123986 | 236 | 14 | 18826 |
| **Age group (years)** | | | | | | | | | | | | |
| 18-49 | 3516 (65) | 31 (20) | 312770 (54) | 2782 (64) | 22 (18) | 231848 (53) | 630 (69) | 8 (32) | 71115 (57) | 28 (12) | 0 (0) | 930 (5) |
| 50-69 | 1298 (24) | 31 (20) | 159863 (28) | 1059 (24) | 23 (19) | 121129 (28) | 213 (23) | 6 (24) | 34872 (28) | 154 (65) | 6 (43) | 9905 (53) |
| 70-79 | 363 (7) | 40 (26) | 65616 (11) | 313 (7) | 35 (28) | 52470 (12) | 44 (5) | 4 (16) | 11321 (9) | 49 (21) | 5 (36) | 6053 (32) |
| 80+ | 202 (4) | 53 (34) | 38578 (7) | 169 (4) | 43 (35) | 30659 (7) | 27 (3) | 7 (28) | 6678 (5) | 5 (2) | 3 (21) | 1938 (10) |
| Median (Interquartile range) | 42 (32-57) | 73 (59-82) | 47 (34-65) | 43 (32-57) | 75 (62-82) | 47 (34-66) | 39 (30-54) | 61 (44-82) | 45 (32-62) | 58 (55-69) | 71 (67-79) | 67 (58-75) |
| **Sex** | | | | | | | | | | | | |
| Female | 2359 (44) | 86 (55) | 230659 (40) | 1917 (44) | 67 (54) | 171074 (39) | 371 (41) | 13 (52) | 52049 (42) | 150 (64) | 11 (79) | 9578 (51) |
| Male | 3020 (56) | 69 (45) | 346168 (60) | 2406 (56) | 56 (46) | 265032 (61) | 543 (59) | 12 (48) | 71937 (58) | 86 (36) | 3 (21) | 9248 (49) |
| **Epidemiological week** | | | | | | | | | | | | |
| 22-25 (May 30-June 26) | 69 (1) | 5 (3) | 43045 (8) | 62 (1) | 4 (4) | 34685 (8) | 6 (0) | 1 (4) | 7172 (5) | 7 (3) | 1 (7) | 2734 (15) |
| 26-30 (June 27-July 31) | 220 (4) | 6 (4) | 102224 (18) | 186 (5) | 5 (5) | 76716 (18) | 30 (3) | 1 (4) | 22017 (18) | 6 (2) | 0 (0) | 4492 (25) |
| 31-35 (August 1-Sept 4) | 1864 (35) | 52 (33) | 182100 (31) | 1482 (34) | 41 (34) | 136391 (31) | 326 (35) | 6 (24) | 40328 (32) | 90 (38) | 4 (28) | 5461 (29) |
| 36-39 (Sept 5-Oct 2) | 3226 (60) | 92 (59) | 249458 (42) | 2593 (60) | 73 (59) | 188314 (42) | 552 (61) | 17 (68) | 54469 (45) | 133 (57) | 9 (63) | 6139 (33) |
| **Interval between doses** | | | | | | | | | | | | |
| 21-34 days (3-4 weeks) | 423 (8) | 17 (11) | 23062 (4) | 313 (7) | 8 (7) | 13898 (3) | 94 (10) | 7 (28) | 8213 (7) | 6 (3) | 1 (7) | 180 (1) |
| 35-48 days (5-6 weeks) | 464 (9) | 6 (4) | 34960 (6) | 293 (7) | 5 (4) | 20439 (5) | 142 (16) | 1 (4) | 11924 (10) | 19 (8) | 0 (0) | 794 (4) |
| 49-62 days (7-8 weeks) | 1490 (28) | 13 (8) | 139531 (24) | 1193 (28) | 9 (7) | 101395 (23) | 260 (28) | 3 (12) | 33452 (27) | 84 (36) | 2 (14) | 4876 (26) |
| 63-83 days (9-11 weeks) | 1575 (29) | 41 (26) | 179057 (31) | 1283 (30) | 37 (30) | 134852 (31) | 262 (29) | 4 (16) | 39908 (32) | 105 (44) | 8 (57) | 10180 (54) |
| 84-111 days (12-15 weeks) | 908 (17) | 45 (29) | 122361 (21) | 766 (18) | 38 (31) | 97199 (22) | 122 (13) | 6 (24) | 22360 (18) | 21 (9) | 3 (21) | 2354 (13) |
| 112+ days (16+ weeks) | 519 (10) | 33 (21) | 77856 (13) | 475 (11) | 26 (21) | 68323 (16) | 34 (4) | 4 (16) | 8129 (7) | 1 (0) | 0 (0) | 442 (2) |
| **Time since second dose** | | | | | | | | | | | | |
| 0-13 days (0-1 weeks) | 745 (14) | 12 (8) | 64574 (11) | 616 (15) | 10 (8) | 48138 (11) | 118 (13) | 1 (4) | 14125 (12) | 6 (2) | 1 (7) | 1924 (11) |
| 14-27 days (2-3 weeks) | 324 (6) | 16 (11) | 72539 (12) | 256 (6) | 11 (9) | 55110 (12) | 63 (7) | 3 (12) | 15315 (12) | 4 (1) | 0 (0) | 2185 (12) |
| 28-55 days (4-7 weeks) | 1234 (24) | 26 (18) | 162071 (28) | 989 (23) | 20 (16) | 124151 (28) | 212 (23) | 5 (20) | 33187 (27) | 17 (8) | 0 (0) | 3970 (20) |
| 56-83 days (8-11 weeks) | 1743 (31) | 41 (26) | 156632 (27) | 1358 (32) | 29 (24) | 116747 (27) | 312 (34) | 9 (36) | 34862 (28) | 62 (26) | 3 (21) | 4313 (23) |
| 84-111 days (12-15 weeks) | 941 (17) | 48 (31) | 80130 (14) | 767 (18) | 45 (37) | 60368 (14) | 160 (17) | 3 (12) | 17915 (15) | 112 (47) | 5 (35) | 4872 (26) |
| 112-139 days (16-19 weeks) | 277 (5) | 9 (6) | 26489 (4) | 238 (5) | 7 (5) | 20682 (4) | 35 (5) | 2 (8) | 5286 (4) | 34 (14) | 4 (28) | 1540 (8) |
| 140-167 days (20-23 weeks) | 87 (1) | 2 (1) | 12433 (2) | 76 (1) | 1 (1) | 9721 (2) | 9 (1) | 1 (4) | 2537 (2) | 1 (0) | 1 (7) | 16 (0) |
| 168-195 days (24-27 weeks) | 19 (0) | 0 (0) | 1394 (0) | 16 (0) | 0 (0) | 782 (0) | 3 (0) | 0 (0) | 604 (0) | 0 (0) | 0 (0) | 2 (0) |
| 196+ days (28+ weeks) | 9 (0) | 1 (1) | 565 (0) | 7 (0) | 0 (0) | 407 (0) | 2 (0) | 1 (4) | 155 (0) | 0 (0) | 0 (0) | 4 (0) |
| Median | 62 | 74 | 54 | 62 | 78 | 54 | 61 | 70 | 55 | 91 | 105 | 65 |
| Range (days) | 0-240 | 1-210 | 0-256 | 0-240 | 3-143 | 0-256 | 0-219 | 1-210 | 0-238 | 2-154 | 6-154 | 0-216 |
| Interquartile range (days) | 36-83 | 40-95 | 29-79 | 36-84 | 46-96 | 29-79 | 36-81 | 46-84 | 29-80 | 76-104 | 82-116 | 32-93 |

Hosp = Hospitalized cases

**Supplementary Table 3 Cont'd. Profile of participants ≥18-year-olds, by vaccine type (regardless of time since vaccination), Quebec, Canada**

| | **Mixed mRNA** | | | **Mixed ChAdOX1 plus mRNA** | | |
|---|---|---|---|---|---|---|
| | **Cases (n, %)** | **Hosp (n, %)** | **Controls (n, %)** | **Cases (n, %)** | **Hosp (n, %)** | **Controls (n, %)** |
| **Total N** | 142 | 7 | 16735 | 253 | 7 | 29110 |
| **Age group (years)** | | | | | | |
| 18-49 | 104 (73) | 1 (14) | 9807 (59) | 62 (25) | 0 (0) | 5974 (21) |
| 50-69 | 26 (18) | 2 (29) | 3862 (23) | 170 (67) | 3 (43) | 19143 (66) |
| 70-79 | 6 (4) | 1 (14) | 1825 (11) | 17 (7) | 2 (29) | 3287 (11) |
| 80+ | 6 (4) | 3 (43) | 1241 (7) | 4 (2) | 2 (29) | 706 (2) |
| Median (Interquartile range) | 39 (32-51) | 70 (61-92) | 44 (31-65) | 55 (50-59) | 79 (61-87) | 57 (51-62) |
| **Sex** | | | | | | |
| Female | 71 (50) | 1 (14) | 9199 (55) | 164 (65) | 2 (29) | 15188 (52) |
| Male | 71 (50) | 6 (86) | 7536 (45) | 89 (35) | 5 (71) | 13922 (48) |
| **Epidemiological week** | | | | | | |
| 22-25 (May 30-June 26) | 1 (1) | 0 (0) | 1188 (7) | 0 (0) | 0 (0) | 762 (3) |
| 26-30 (June 27-July 31) | 4 (3) | 0 (0) | 3491 (21) | 8 (2) | 0 (0) | 6282 (22) |
| 31-35 (August 1-Sept 4) | 56 (39) | 5 (71) | 5381 (32) | 83 (33) | 2 (28) | 9599 (33) |
| 36-39 (Sept 5-Oct 2) | 81 (57) | 2 (29) | 6675 (40) | 162 (64) | 5 (72) | 12467 (42) |
| **Interval between doses** | | | | | | |
| 21-34 days (3-4 weeks) | 16 (11) | 2 (29) | 951 (6) | 1 (0) | 1 (14) | 58 (0) |
| 35-48 days (5-6 weeks) | 29 (20) | 0 (0) | 2597 (16) | 1 (0) | 0 (0) | 126 (0) |
| 49-62 days (7-8 weeks) | 37 (26) | 1 (14) | 4684 (28) | 62 (25) | 0 (0) | 5289 (18) |
| 63-83 days (9-11 weeks) | 30 (21) | 0 (0) | 4297 (26) | 128 (51) | 1 (14) | 15115 (52) |
| 84-111 days (12-15 weeks) | 20 (14) | 1 (14) | 2802 (17) | 44 (17) | 4 (57) | 6159 (21) |
| 112+ days (16+ weeks) | 10 (7) | 3 (43) | 1404 (8) | 17 (7) | 1 (14) | 2363 (8) |
| **Time since second dose** | | | | | | |
| 0-13 d (0-1 w) | 11 (8) | 1 (14) | 2311 (14) | 11 (4) | 0 (0) | 3382 (11) |
| 14-27 d (2-3 w) | 5 (4) | 2 (29) | 2114 (13) | 7 (3) | 0 (0) | 3363 (12) |
| 28-55 d (4-7 w) | 33 (23) | 1 (14) | 4733 (28) | 48 (19) | 1 (14) | 7334 (26) |
| 56-83 d (8-11 w) | 73 (51) | 3 (43) | 5023 (30) | 118 (46) | 2 (28) | 9694 (33) |
| 84-111 d (12-15 w) | 14 (10) | 0 (0) | 1847 (11) | 68 (27) | 4 (57) | 5230 (18) |
| 112-139 d (16-19 w) | 4 (3) | 0 (0) | 521 (3) | 0 (0) | 0 (0) | 101 (0) |
| 140-167 d (20-23 w) | 2 (1) | 0 (0) | 175 (1) | 1 (0) | 0 (0) | 5 (0) |
| 168-195 d (24-27 w) | 0 (0) | 0 (0) | 8 (0) | 0 (0) | 0 (0) | 1 (0) |
| 196+ d (28+ w) | 0 (0) | 0 (0) | 3 (0) | 0 (0) | 0 (0) | 0 (0) |
| Median | 64 | 48 | 51 | 71 | 84 | 57 |
| Range (days) | 0-148 | 2-73 | 0-236 | 2-143 | 52-107 | 0-188 |
| Interquartile range (days) | 48-77 | 15-69 | 26-74 | 55-84 | 76-104 | 30-78 |

Hosp = Hospitalized cases

**Supplementary Table 4. Two-dose vaccine effectiveness against infection and hospitalization (≥14 days post-vaccination), by vaccine type, ≥18-year-olds, British Columbia and Quebec, Canada**

| | British Columbia | | | | Quebec | | | |
|---|---|---|---|---|---|---|---|---|
| | Sample sizes | | VE (95% CI) | | Sample sizes | | VE (95% CI) | |
| | Case N | Control N | Crude | Adjusted[1] | Case N | Control N | Crude | Adjusted[1] |
| **Infection** | | | | | | | | |
| Two any vaccines | 7644 | 252349 | 88 (87, 88) | 90 (89, 90) | 5106 | 554883 | 83 (82, 83) | 88 (88, 89) |
| Two any mRNA vaccines | 6595 | 230188 | 88 (88, 89) | 90 (90, 91) | 4634 | 512253 | 83 (82, 84) | 89 (88, 89) |
| Two BNT162b2 (mRNA) (Pfizer-BioNTech, Comirnaty) | 4876 | 168831 | 88 (88, 89) | 90 (90, 90) | 3707 | 387968 | 82 (81, 83) | 88 (88, 89) |
| Two mRNA-1273 (mRNA) (Moderna, Spikevax) | 1209 | 41889 | 88 (88, 89) | 91 (90, 91) | 796 | 109861 | 86 (85, 87) | 90 (90, 91) |
| Two mixed mRNA doses | 509 | 19458 | 89 (88, 90) | 91 (90, 92) | 131 | 14424 | 83 (80, 86) | 89 (87, 91) |
| Two ChAdOx1 (AstraZeneca, Vaxzevria/COVISHIELD) | 645 | 8077 | 67 (65, 70) | 71 (69, 74) | 230 | 16902 | 75 (71, 78) | 73 (69, 77) |
| Two mixed ChAdOx1 + mRNA | 404 | 14084 | 88 (87, 89) | 90 (89, 91) | 242 | 25728 | 82 (80, 84) | 87 (85, 89) |
| Unvaccinated | 18809 | 76621 | Reference | | 11366 | 212918 | Reference | |
| **Hospitalization** | | | | | | | | |
| Two any vaccines | 197 | 252349 | 96 (95, 96) | 98 (97, 98) | 163 | 554883 | 91 (89, 92) | 97 (96, 97) |
| Two any mRNA vaccines | 178 | 230188 | 96 (95, 96) | 98 (97, 98) | 143 | 512253 | 92 (90, 93) | 97 (96, 97) |
| Two BNT162b2 (mRNA) (Pfizer-BioNTech, Comirnaty) | 122 | 168831 | 96 (95, 97) | 98 (97, 98) | 113 | 387968 | 91 (89, 93) | 97 (96, 97) |
| Two mRNA-1273 (mRNA) (Moderna, Spikevax) | 38 | 41889 | 95 (93, 96) | 97 (96, 98) | 24 | 109861 | 93 (90, 96) | 97 (95, 98) |
| Two mixed mRNA doses | 18 | 19458 | 95 (92, 97) | 98 (96, 99) | 6 | 14424 | 87 (72, 94) | 94 (87, 97) |
| Two ChAdOx1 (AstraZeneca, Vaxzevria/COVISHIELD) | 17 | 8077 | 88 (81, 93) | 94 (90, 96) | 13 | 16902 | 77 (60, 87) | 94 (89, 97) |
| Two mixed ChAdOx1 + mRNA | 2 | 14084 | 99 (97, 100) | 99 (98, 100) | 7 | 25728 | 92 (83, 96) | 98 (95, 99) |
| Unvaccinated | 1365 | 76621 | Reference | | 702 | 212918 | Reference | |

VE=vaccine effectiveness; 95% CI = 95% confidence interval

[1] Unless otherwise specified, all VE estimates adjusted for age group (18-49, 50-69, 70-79, ≥80 years); sex (male, female); individual epidemiological week of the analysis period (weeks 22-39, categorical); and region of the province (5 in each province).

**Supplementary Table 5. Two-dose vaccine effectiveness against infection (≥14 days post-vaccination), by age group and vaccine type, British Columbia and Quebec, Canada**

| | British Columbia | | | | Quebec | | | |
|---|---|---|---|---|---|---|---|---|
| | Sample sizes | | VE (95% CI) | | Sample sizes | | VE (95% CI) | |
| | Case N | Control N | Crude | Adjusted[1] | Case N | Control N | Crude | Adjusted[1] |
| **18-49 years** | | | | | | | | |
| Two any vaccines | 4693 | 145047 | 87 (87, 88) | 89 (89, 90) | 3007 | 283589 | 82 (81, 83) | 89 (89, 90) |
| Two any mRNA vaccines | 4171 | 134464 | 88 (87, 88) | 90 (90, 90) | 2922 | 277438 | 82 (82, 83) | 89 (89, 90) |
| Two BNT162b2 (mRNA) (Pfizer-BioNTech, Comirnaty) | 3103 | 100530 | 88 (87, 88) | 90 (89, 90) | 2298 | 206318 | 81 (80, 82) | 89 (88, 89) |
| Two mRNA-1273 (mRNA) (Moderna, Spikevax) | 795 | 25637 | 88 (87, 89) | 90 (89, 91) | 527 | 62597 | 86 (85, 87) | 91 (90, 92) |
| Two mixed mRNA doses | 273 | 8292 | 87 (85, 88) | 89 (88, 91) | 97 | 8523 | 81 (77, 84) | 89 (86, 91) |
| Two ChAdOx1 (AstraZeneca, Vaxzevria/COVISHIELD) | 250 | 2641 | 62 (57, 67) | 68 (63, 72) | 28 | 824 | 43 (17, 61) | 68 (53, 78) |
| Two mixed ChAdOx1 + mRNA | 272 | 7942 | 86 (85, 88) | 89 (87, 90) | 57 | 5327 | 82 (77, 86) | 90 (87, 92) |
| Unvaccinated | 14576 | 57847 | Reference | | 9464 | 159076 | Reference | |
| **50-69 years** | | | | | | | | |
| Two any vaccines | 2197 | 68407 | 87 (86, 88) | 90 (89, 91) | 1497 | 168612 | 77 (75, 78) | 86 (85, 87) |
| Two any mRNA vaccines | 1680 | 57041 | 88 (88, 89) | 91 (91, 92) | 1183 | 142701 | 78 (76, 80) | 87 (86, 88) |
| Two BNT162b2 (mRNA) (Pfizer-BioNTech, Comirnaty) | 1205 | 39629 | 88 (87, 89) | 91 (90, 91) | 957 | 108114 | 77 (75, 78) | 87 (85, 88) |
| Two mRNA-1273 (mRNA) (Moderna, Spikevax) | 307 | 10770 | 89 (87, 90) | 92 (91, 93) | 200 | 31261 | 83 (80, 85) | 89 (88, 91) |
| Two mixed mRNA doses | 167 | 6640 | 90 (88, 91) | 92 (91, 94) | 26 | 3326 | 79 (70, 86) | 88 (82, 92) |
| Two ChAdOx1 (AstraZeneca, Vaxzevria/COVISHIELD) | 388 | 5319 | 71 (67, 74) | 73 (70, 76) | 149 | 8889 | 56 (48, 63) | 74 (69, 78) |
| Two mixed ChAdOx1 + mRNA | 129 | 6047 | 91 (90, 93) | 93 (91, 94) | 165 | 17022 | 74 (70, 78) | 87 (85, 89) |
| Unvaccinated | 3463 | 13824 | Reference | | 1645 | 43380 | Reference | |
| **≥70 years[2]** | | | | | | | | |
| Two any vaccines | 754 | 38895 | 88 (86, 89) | 91 (90, 92) | 602 | 102682 | 76 (72, 79) | 85 (82, 87) |
| Two any mRNA vaccines | 744 | 38683 | 88 (86, 89) | 91 (90, 92) | 529 | 92114 | 77 (73, 80) | 85 (82, 87) |
| Two BNT162b2 (mRNA) (Pfizer-BioNTech, Comirnaty) | 568 | 28672 | 87 (86, 89) | 91 (90, 92) | 452 | 73536 | 75 (71, 79) | 84 (81, 86) |
| Two mRNA-1273 (mRNA) (Moderna, Spikevax) | 107 | 5482 | 87 (85, 90) | 93 (91, 94) | 69 | 16003 | 82 (77, 87) | 88 (84, 91) |
| Two mixed mRNA doses | 69 | 4526 | 90 (87, 92) | 93 (91, 95) | 8 | 2575 | 87 (74, 94) | 92 (83, 96) |
| Two ChAdOx1 (AstraZeneca, Vaxzevria/COVISHIELD) | 7 | 117 | 62 (17, 82) | 73 (42, 88) | 53 | 7189 | 70 (60, 78) | 81 (74, 86) |
| Two mixed ChAdOx1 + mRNA | 3 | 95 | 80 (36, 94) | 85 (52, 95) | 20 | 3379 | 76 (62, 85) | 88 (81, 93) |
| Unvaccinated | 770 | 4950 | Reference | | 257 | 10462 | Reference | |
| **≥80 years** | | | | | | | | |
| Two any vaccines | 221 | 14738 | 85 (81, 87) | 89 (86, 91) | 197 | 36296 | 68 (58, 75) | 77 (70, 83) |
| Two any mRNA vaccines | 219 | 14699 | 85 (82, 88) | 89 (86, 91) | 188 | 33974 | 67 (57, 75) | 77 (70, 83) |
| Two BNT162b2 (mRNA) (Pfizer-BioNTech, Comirnaty) | 183 | 11030 | 83 (79, 86) | 87 (84, 90) | 157 | 27080 | 65 (54, 74) | 76 (68, 82) |
| Two mRNA-1273 (mRNA) (Moderna, Spikevax) | 21 | 2061 | 90 (84, 93) | 93 (90, 96) | 27 | 5875 | 73 (57, 82) | 80 (68, 87) |
| Two mixed mRNA doses | 15 | 1608 | 90 (84, 94) | 93 (87, 96) | 4 | 1019 | 77 (36, 91) | 83 (52, 94) |
| Two ChAdOx1 (AstraZeneca, Vaxzevria/COVISHIELD) | 1 | 24 | NE | NE | 5 | 1733 | 83 (57, 93) | 85 (62, 94) |
| Two mixed ChAdOx1 + mRNA | 1 | 15 | NE | NE | 4 | 589 | 59 (-11, 85) | 74 (29, 91) |
| Unvaccinated | 205 | 2092 | Reference | | 76 | 4534 | Reference | |

VE=vaccine effectiveness; 95%CI=95% confidence interval; NE=Not estimable

[1] Unless otherwise specified, all VE estimates adjusted for sex (male, female); individual epidemiological week of the analysis period (weeks 22-39, categorical); and region of the province (5 in each province).
[2] Among those ≥70 years, VE additionally adjusted for 70-79 and ≥80 years.

**Supplementary Table 6. Two-dose vaccine effectiveness against hospitalization (≥14 days post-vaccination), by age group and vaccine type, British Columbia and Quebec, Canada**

| | British Columbia | | | | Quebec | | | |
|---|---|---|---|---|---|---|---|---|
| | Sample sizes | | VE (95% CI) | | Sample sizes | | VE (95% CI) | |
| | Case N | Control N | Crude | Adjusted[1] | Case N | Control N | Crude | Adjusted[1] |
| **18-49 years[2]** | | | | | | | | |
| Two any vaccines | 26 | 145047 | 98 (97, 99) | 99 (98, 99) | 28 | 283589 | 95 (93, 97) | 97 (96, 98) |
| Two any mRNA vaccines | 25 | 134464 | 98 (97, 99) | 98 (98, 99) | 28 | 277438 | 95 (93, 97) | 97 (96, 98) |
| Two BNT162b2 (mRNA) (Pfizer-BioNTech, Comirnaty) | 13 | 100530 | 99 (98, 99) | 99 (98, 99) | 20 | 206318 | 95 (92, 97) | 97 (96, 98) |
| Two mRNA-1273 (mRNA) (Moderna, Spikevax) | 9 | 25637 | 96 (93, 98) | 97 (95, 99) | 7 | 62597 | 94 (88, 97) | 96 (92, 98) |
| Two mixed mRNA doses | 3 | 8292 | 96 (88, 99) | 97 (91, 99) | 1 | 8523 | 94 (58, 99) | 97 (76, 100) |
| Two ChAdOx1 (AstraZeneca, Vaxzevria/COVISHIELD) | 1 | 2641 | 96 (72, 99) | 97 (78, 100) | 0 | 824 | NE | NE |
| Two mixed ChAdOx1 + mRNA | 0 | 7942 | NE | NE | 0 | 5327 | NE | NE |
| Unvaccinated | 558 | 57847 | Reference | | 317 | 159076 | Reference | |
| **50-69 years** | | | | | | | | |
| Two any vaccines | 65 | 68407 | 97 (97, 98) | 98 (97, 98) | 38 | 168612 | 96 (95, 97) | 98 (97, 99) |
| Two any mRNA vaccines | 50 | 57041 | 98 (97, 98) | 98 (98, 99) | 29 | 142701 | 97 (95, 98) | 98 (97, 99) |
| Two BNT162b2 (mRNA) (Pfizer-BioNTech, Comirnaty) | 31 | 39629 | 98 (97, 99) | 98 (98, 99) | 21 | 108114 | 97 (95, 98) | 98 (97, 99) |
| Two mRNA-1273 (mRNA) (Moderna, Spikevax) | 13 | 10770 | 97 (94, 98) | 98 (96, 99) | 6 | 31261 | 97 (93, 99) | 98 (96, 99) |
| Two mixed mRNA doses | 6 | 6640 | 98 (95, 99) | 98 (96, 99) | 2 | 3326 | 90 (61, 98) | 95 (78, 99) |
| Two ChAdOx1 (AstraZeneca, Vaxzevria/COVISHIELD) | 14 | 5319 | 93 (88, 96) | 94 (89, 96) | 6 | 8889 | 89 (76, 95) | 94 (87, 97) |
| Two mixed ChAdOx1 + mRNA | 1 | 6047 | 100 (97, 100) | 100 (97, 100) | 3 | 17022 | 97 (91, 99) | 99 (96, 100) |
| Unvaccinated | 523 | 13824 | Reference | | 271 | 43380 | Reference | |
| **≥70 years[3,4]** | | | | | | | | |
| Two any vaccines | 106 | 38895 | 95 (94, 96) | 97 (96, 97) | 97 | 102682 | 91 (89, 93) | 94 (93, 96) |
| Two any mRNA vaccines | 103 | 38683 | 95 (94, 96) | 97 (96, 97) | 86 | 92114 | 91 (89, 94) | 94 (93, 96) |
| Two BNT162b2 (mRNA) (Pfizer-BioNTech, Comirnaty) | 78 | 28672 | 95 (94, 96) | 97 (95, 97) | 72 | 73536 | 91 (88, 93) | 94 (92, 96) |
| Two mRNA-1273 (mRNA) (Moderna, Spikevax) | 16 | 5482 | 95 (92, 97) | 97 (95, 98) | 11 | 16003 | 94 (88, 97) | 96 (92, 98) |
| Two mixed mRNA doses | 9 | 4526 | 97 (93, 98) | 97 (95, 99) | 3 | 2575 | 89 (66, 97) | 93 (77, 98) |
| Two ChAdOx1 (AstraZeneca, Vaxzevria/COVISHIELD) | 2 | 117 | 70 (-21, 93) | 79 (14, 95) | 7 | 7189 | 91 (81, 96) | 94 (88, 97) |
| Two mixed ChAdOx1 + mRNA | 1 | 95 | 82 (-32, 97) | 86 (-2, 98) | 4 | 3379 | 89 (71, 96) | 95 (85, 98) |
| Unvaccinated | 284 | 4950 | Reference | | 114 | 10462 | Reference | |
| **≥80 years[5]** | | | | | | | | |
| Two any vaccines | 60 | 14738 | 91 (88, 94) | 94 (91, 96) | 52 | 36296 | 83 (75, 89) | 88 (82, 92) |
| Two any mRNA vaccines | 60 | 14699 | 91 (88, 94) | 94 (91, 96) | 47 | 33974 | 84 (75, 89) | 89 (83, 93) |
| Two BNT162b2 (mRNA) (Pfizer-BioNTech, Comirnaty) | 45 | 11030 | 91 (88, 94) | 94 (91, 96) | 38 | 27080 | 84 (74, 90) | 89 (82, 93) |
| Two mRNA-1273 (mRNA) (Moderna, Spikevax) | 11 | 2061 | 89 (79, 94) | 93 (86, 96) | 7 | 5875 | 86 (69, 94) | 90 (77, 96) |
| Two mixed mRNA doses | 4 | 1608 | 95 (86, 98) | 96 (89, 99) | 2 | 1019 | 77 (5, 95) | 82 (27, 96) |
| Two ChAdOx1 (AstraZeneca, Vaxzevria/COVISHIELD) | 0 | 24 | NE | NE | 3 | 1733 | 80 (35, 94) | 82 (43, 95) |
| Two mixed ChAdOx1 + mRNA | 0 | 15 | NE | NE | 2 | 589 | NE | NE |
| Unvaccinated | 98 | 2092 | Reference | | 39 | 4534 | Reference | |

VE=vaccine effectiveness; 95%CI=95% confidence interval; NE=Not estimable

[1] Unless otherwise specified, all VE estimates adjusted for sex (male, female); individual epidemiological week of the analysis period (weeks 22-39, categorical); and region of the province (5 in each province).
[2] In Quebec, adjusted VE estimates against hospitalization in 18-49-year-olds were adjusted for calendar time bi-weekly owing to sample size.
[3] Among those ≥70 years, VE additionally adjusted for 70-79 and ≥80 years in both provinces.
[4] In Quebec, adjusted VE estimates against hospitalization in ≥70-year-olds were adjusted for calendar time tri-weekly owing to sample size.
[5] In Quebec, adjusted VE estimates against hospitalization in ≥80-year-olds were adjusted for calendar time tri-weekly owing to sample size.

**Supplementary Table 7. Two-dose vaccine effectiveness against infection and hospitalization (≥14 days post-vaccination), by sex and vaccine type, ≥18-year-olds, British Columbia and Quebec, Canada**

| | British Columbia | | | | Quebec | | | |
|---|---|---|---|---|---|---|---|---|
| | Sample sizes | | VE (95% CI) | | Sample sizes | | VE (95% CI) | |
| | Case N | Control N | Crude | Adjusted[1] | Case N | Control N | Crude | Adjusted[1] |
| **Female, Infection** | | | | | | | | |
| Two any vaccines | 4149 | 146358 | 87 (87, 88) | 90 (89, 90) | 2818 | 330058 | 80 (79, 80) | 87 (86, 87) |
| Two any mRNA vaccines | 3716 | 135160 | 88 (87, 88) | 90 (89, 90) | 2650 | 309456 | 79 (78, 80) | 87 (86, 87) |
| Two BNT162b2 (mRNA) (Pfizer-BioNTech, Comirnaty) | 2733 | 100157 | 88 (87, 88) | 90 (89, 90) | 2112 | 237473 | 79 (78, 80) | 86 (86, 87) |
| Two mRNA-1273 (mRNA) (Moderna, Spikevax) | 695 | 23727 | 87 (86, 88) | 90 (89, 91) | 471 | 63990 | 82 (81, 84) | 88 (87, 89) |
| Two mixed mRNA doses | 287 | 11270 | 89 (87, 90) | 91 (89, 92) | 67 | 7993 | 80 (74, 84) | 87 (83, 90) |
| Two ChAdOx1 (AstraZeneca, Vaxzevria/COVISHIELD) | 258 | 3724 | 69 (64, 73) | 72 (68, 76) | 83 | 8300 | 76 (70, 81) | 74 (67, 79) |
| Two mixed ChAdOx1 + mRNA | 175 | 7474 | 89 (88, 91) | 91 (90, 92) | 85 | 12302 | 83 (79, 87) | 87 (84, 90) |
| Unvaccinated | 8718 | 39296 | Reference | | 5561 | 133526 | Reference | |
| **Female, Hospitalization** | | | | | | | | |
| Two any vaccines | 84 | 146358 | 96 (95, 97) | 98 (97, 98) | 71 | 330058 | 90 (87, 92) | 96 (95, 97) |
| Two any mRNA vaccines | 76 | 135160 | 96 (95, 97) | 98 (97, 98) | 63 | 309456 | 91 (88, 93) | 96 (95, 97) |
| Two BNT162b2 (mRNA) (Pfizer-BioNTech, Comirnaty) | 49 | 100157 | 97 (95, 97) | 98 (97, 98) | 50 | 237473 | 90 (87, 93) | 96 (95, 97) |
| Two mRNA-1273 (mRNA) (Moderna, Spikevax) | 21 | 23727 | 94 (90, 96) | 96 (94, 98) | 12 | 63990 | 92 (85, 95) | 96 (93, 98) |
| Two mixed mRNA doses | 6 | 11270 | 96 (92, 98) | 98 (96, 99) | 1 | 7993 | 94 (60, 99) | 97 (82, 100) |
| Two ChAdOx1 (AstraZeneca, Vaxzevria/COVISHIELD) | 7 | 3724 | 87 (72, 94) | 92 (83, 96) | 3 | 8300 | 84 (49, 95) | 95 (85, 99) |
| Two mixed ChAdOx1 + mRNA | 1 | 7474 | 99 (93, 100) | 99 (95, 100) | 5 | 12302 | 82 (55, 92) | 94 (85, 97) |
| Unvaccinated | 555 | 39296 | Reference | | 295 | 133526 | Reference | |
| **Male, Infection** | | | | | | | | |
| Two any vaccines | 3495 | 105991 | 88 (87, 88) | 90 (90, 90) | 2288 | 224825 | 86 (85, 87) | 90 (90, 91) |
| Two any mRNA vaccines | 2879 | 95028 | 89 (88, 89) | 91 (90, 91) | 1984 | 202797 | 87 (86, 87) | 91 (90, 91) |
| Two BNT162b2 (mRNA) (Pfizer-BioNTech, Comirnaty) | 2143 | 68674 | 88 (88, 89) | 90 (90, 91) | 1595 | 150495 | 86 (85, 86) | 90 (89, 90) |
| Two mRNA-1273 (mRNA) (Moderna, Spikevax) | 514 | 18162 | 90 (89, 90) | 92 (91, 92) | 325 | 45871 | 90 (89, 91) | 93 (92, 93) |
| Two mixed mRNA doses | 222 | 8188 | 90 (89, 91) | 91 (90, 92) | 64 | 6431 | 86 (83, 89) | 90 (87, 92) |
| Two ChAdOx1 (AstraZeneca, Vaxzevria/COVISHIELD) | 387 | 4353 | 67 (63, 70) | 70 (67, 74) | 147 | 8602 | 77 (72, 80) | 73 (68, 78) |
| Two mixed ChAdOx1 + mRNA | 229 | 6610 | 87 (85, 89) | 89 (88, 91) | 157 | 13426 | 84 (81, 86) | 87 (85, 89) |
| Unvaccinated | 10091 | 37325 | Reference | | 5805 | 79392 | Reference | |
| **Male, Hospitalization** | | | | | | | | |
| Two any vaccines | 113 | 105991 | 95 (94, 96) | 98 (97, 98) | 92 | 224825 | 92 (90, 94) | 97 (96, 98) |
| Two any mRNA vaccines | 102 | 95028 | 95 (94, 96) | 98 (97, 98) | 80 | 202797 | 92 (90, 94) | 97 (96, 98) |
| Two BNT162b2 (mRNA) (Pfizer-BioNTech, Comirnaty) | 73 | 68674 | 95 (94, 96) | 98 (97, 98) | 63 | 150495 | 92 (89, 94) | 97 (96, 98) |
| Two mRNA-1273 (mRNA) (Moderna, Spikevax) | 17 | 18162 | 96 (93, 97) | 98 (96, 99) | 12 | 45871 | 95 (91, 97) | 97 (96, 99) |
| Two mixed mRNA doses | 12 | 8188 | 93 (88, 96) | 97 (95, 98) | 5 | 6431 | 85 (63, 94) | 93 (82, 97) |
| Two ChAdOx1 (AstraZeneca, Vaxzevria/COVISHIELD) | 10 | 4353 | 89 (80, 94) | 95 (90, 97) | 10 | 8602 | 77 (58, 88) | 93 (88, 97) |
| Two mixed ChAdOx1 + mRNA | 1 | 6610 | 99 (95, 100) | 100 (97, 100) | 2 | 13426 | 97 (88, 99) | 99 (96, 100) |
| Unvaccinated | 810 | 37325 | Reference | | 407 | 79392 | Reference | |

VE=vaccine effectiveness; 95%CI=95% confidence interval

[1] Unless otherwise specified, all VE estimates adjusted for age group (18-49, 50-69, 70-79, ≥80 years); individual epidemiological week of the analysis period (weeks 22-39, categorical); and region of the province (5 in each province).

**Supplementary Table 8. Two-dose vaccine effectiveness (VE) against infection and hospitalization (≥14 days post-vaccination), Alpha and Delta variants, by vaccine type ≥18-year-olds, British Columbia and Quebec, Canada**

| | British Columbia | | | | Quebec | | | |
|---|---|---|---|---|---|---|---|---|
| | Sample sizes | | VE (95% CI) | | Sample sizes | | VE (95% CI) | |
| | Case N | Control N | Crude | Adjusted[1] | Case N | Control N | Crude | Adjusted[1] |
| **Delta Infection** | | | | | | | | |
| Two any vaccines | 3911 | 252349 | 90 (89, 90) | 91 (90, 91) | 3422 | 554883 | 79 (78, 80) | 89 (88, 89) |
| Two any mRNA vaccines | 3320 | 230188 | 90 (90, 91) | 91 (91, 92) | 3093 | 512253 | 80 (79, 81) | 89 (89, 90) |
| Two BNT162b2 (mRNA) (Pfizer-BioNTech, Comirnaty) | 2501 | 168831 | 90 (90, 91) | 91 (91, 92) | 2489 | 387968 | 78 (77, 79) | 89 (88, 89) |
| Two mRNA-1273 (mRNA) (Moderna, Spikevax) | 573 | 41889 | 91 (90, 92) | 92 (91, 93) | 517 | 109861 | 84 (83, 86) | 91 (90, 92) |
| Two mixed mRNA doses | 245 | 19458 | 92 (90, 93) | 92 (91, 93) | 87 | 14424 | 80 (75, 84) | 89 (86, 91) |
| Two ChAdOx1 (AstraZeneca, Vaxzevria/COVISHIELD) | 375 | 8077 | 69 (66, 72) | 70 (66, 73) | 162 | 16902 | 68 (62, 73) | 73 (69, 78) |
| Two mixed ChAdOx1 + mRNA | 216 | 14084 | 90 (88, 91) | 91 (89, 92) | 167 | 25728 | 78 (75, 81) | 88 (85, 89) |
| Unvaccinated | 11500 | 76621 | Reference | | 6349 | 212918 | Reference | |
| **Delta Hospitalization[2]** | | | | | | | | |
| Two any vaccines | 124 | 252349 | 96 (95, 96) | 98 (97, 98) | 99 | 445235 | 91 (89, 93) | 97 (96, 98) |
| Two any mRNA vaccines | 108 | 230188 | 96 (95, 97) | 98 (97, 98) | 85 | 411947 | 96 (94, 96) | 97 (97, 98) |
| Two BNT162b2 (mRNA) (Pfizer-BioNTech, Comirnaty) | 76 | 168831 | 96 (95, 97) | 98 (97, 98) | 70 | 310111 | 95 (94, 96) | 97 (96, 98) |
| Two mRNA-1273 (mRNA) (Moderna, Spikevax) | 24 | 41889 | 95 (92, 97) | 97 (96, 98) | 11 | 90108 | 97 (95, 99) | 98 (96, 99) |
| Two mixed mRNA doses | 8 | 19458 | 96 (93, 98) | 98 (97, 99) | 4 | 11728 | 93 (80, 97) | 95 (86, 98) |
| Two ChAdOx1 (AstraZeneca, Vaxzevria/COVISHIELD) | 14 | 8077 | 85 (74, 91) | 92 (86, 95) | 9 | 11532 | 83 (67, 91) | 94 (89, 97) |
| Two mixed ChAdOx1 + mRNA | 2 | 14084 | 99 (95, 100) | 99 (97, 100) | 5 | 21756 | 95 (88, 98) | 98 (94, 99) |
| Unvaccinated | 873 | 76621 | Reference | | 438 | 94254 | Reference | |
| **Alpha Infection[3]** | | | | | | | | |
| Two any vaccines | 18 | 252349 | 99 (99, 100) | 95 (93, 97) | 47 | 554883 | 99 (98, 99) | 97 (97, 98) |
| Two any mRNA vaccines | 14 | 230188 | 99 (99, 100) | 96 (93, 98) | 42 | 512253 | 99 (99, 99) | 98 (97, 98) |
| Two BNT162b2 (mRNA) (Pfizer-BioNTech, Comirnaty) | 10 | 168831 | 99 (99, 100) | 96 (93, 98) | 32 | 387968 | 99 (98, 99) | 98 (97, 98) |
| Two mRNA-1273 (mRNA) (Moderna, Spikevax) | 3 | 41889 | 99 (98, 100) | 95 (85, 98) | 9 | 109861 | 99 (98, 99) | 97 (95, 99) |
| Two mixed mRNA doses | 1 | 19458 | 99 (96, 100) | 96 (73, 99) | 1 | 14424 | 99 (93, 100) | 98 (87, 100) |
| Two ChAdOx1 (AstraZeneca, Vaxzevria/COVISHIELD) | 4 | 8077 | 95 (86, 98) | 74 (29, 90) | 3 | 16902 | 98 (93, 99) | 94 (80, 98) |
| Two mixed ChAdOx1 + mRNA | 0 | 14084 | NE | NE | 2 | 25728 | 99 (96, 100) | 98 (90, 99) |
| Unvaccinated | 728 | 76621 | Reference | | 1589 | 212918 | Reference | |
| **Alpha Hospitalization[4,5]** | | | | | | | | |
| Two any vaccines | 2 | 252349 | 99 (96, 100) | 97 (88, 99) | 4 | 477261 | 98 (93, 99) | 97 (92, 99) |
| Two any mRNA vaccines | 2 | 230188 | 99 (96, 100) | 97 (87, 99) | 4 | 439590 | 97 (93, 99) | 97 (91, 99) |
| Two BNT162b2 (mRNA) (Pfizer-BioNTech, Comirnaty) | 2 | 168831 | 99 (94, 100) | 96 (83, 99) | 1 | 332460 | 99 (94, 100) | 99 (93, 100) |
| Two mRNA-1273 (mRNA) (Moderna, Spikevax) | 0 | 41889 | NE | NE | 3 | 94626 | 91 (71, 97) | 87 (56, 96) |
| Two mixed mRNA doses | 0 | 19458 | NE | NE | 0 | 12504 | NE | NE |
| Two ChAdOx1 (AstraZeneca, Vaxzevria/COVISHIELD) | 0 | 8077 | NE | NE | 0 | 15273 | NE | NE |
| Two mixed ChAdOx1 + mRNA | 0 | 14084 | NE | NE | 0 | 22398 | NE | NE |
| Unvaccinated | 65 | 76621 | Reference | | 64 | 183043 | Reference | |

VE=vaccine effectiveness; 95%CI=95% confidence interval; NE=Not estimable

[1] Unless otherwise specified, all VE estimates adjusted for age group (18-49, 50-69, 70-79, ≥80 years); sex (male, female); individual epidemiological week of the analysis period (weeks 22-39, categorical); and region of the province (5 in each province).

[2] In Quebec, VE against hospitalizations due to Delta variant assessed only between weeks 31-39 because no hospitalized Delta variant cases were identified prior to that period.

[3] In BC, adjusted VE against Alpha infections adjusted for calendar time as individual epidemiological weeks 22-35, but regrouping weeks 36-39 together owing to sample size considerations.

[4] In BC, adjusted VE against Alpha hospitalizations adjusted for calendar time as individual epidemiological week 22-33, regrouping weeks 34-39 owing to sample size considerations.

[5] In Quebec, VE against Alpha hospitalizations assessed only between weeks 23-38, and adjusted for calendar time by four epidemiological week periods.

**Supplementary Table 9. Two-dose vaccine effectiveness (VE) against infection and hospitalization (≥14 days post-vaccination), Gamma variant, by vaccine type, ≥18-year-olds, British Columbia, Canada**

| | British Columbia | | | |
|---|---|---|---|---|
| | **Sample sizes** | | **VE (95% CI)** | |
| | Case N | Control N | Crude | **Adjusted** |
| **Gamma Infection**[1] | | | | |
| Two any vaccines | 32 | 252349 | 99 (98, 99) | 93 (90, 95) |
| Two any mRNA vaccines | 29 | 230188 | 99 (98, 99) | 93 (90, 96) |
| Two BNT162b2 (mRNA) (Pfizer-BioNTech, Comirnaty) | 24 | 168831 | 98 (98, 99) | 93 (89, 95) |
| Two mRNA-1273 (mRNA) (Moderna, Spikevax) | 3 | 41889 | 99 (98, 100) | 95 (85, 99) |
| Two mixed mRNA doses | 2 | 19458 | 99 (96, 100) | 94 (75, 99) |
| Two ChAdOx1 (AstraZeneca, Vaxzevria/COVISHIELD) | 2 | 8077 | 97 (89, 99) | 90 (61, 98) |
| Two mixed ChAdOx1 + mRNA | 1 | 14084 | 99 (95, 100) | 96 (70, 99) |
| Unvaccinated | 708 | 76621 | Reference | |
| **Gamma Hospitalization**[2] | | | | |
| Two any vaccines | 4 | 252349 | 98 (93, 99) | 95 (87, 98) |
| Two any mRNA vaccines | 4 | 230188 | 97 (93, 99) | 95 (85, 98) |
| Two BNT162b2 (mRNA) (Pfizer-BioNTech, Comirnaty) | 3 | 168831 | 97 (91, 99) | 95 (83, 99) |
| Two mRNA-1273 (mRNA) (Moderna, Spikevax) | 0 | 41889 | NE | NE |
| Two mixed mRNA doses | 1 | 19458 | 92 (44, 99) | NE |
| Two ChAdOx1 (AstraZeneca, Vaxzevria/COVISHIELD) | 0 | 8077 | NE | NE |
| Two mixed ChAdOx1 + mRNA | 0 | 14084 | NE | NE |
| Unvaccinated | 51 | 76621 | Reference | |

VE=vaccine effectiveness; 95%CI=95% confidence interval; NE=Not estimable

[1] In BC, adjusted VE against Gamma infections adjusted for age group (18-49, 50-69, 70-79, ≥80 years); sex (male, female); and region of the province (5) and for calendar time as individual epidemiological weeks 22-35, but regrouping weeks 36-39 together owing to sample size considerations.

[2] In BC, adjusted VE against Gamma hospitalizations adjusted for age group (18-49, 50-69, 70-79, ≥80 years); sex (male, female); and region of the province (5) and for calendar time as individual epidemiological week 22-33, regrouping weeks 34-39 owing to sample size considerations.

**Supplementary Table 10. Two-dose vaccine effectiveness (VE) against infection and hospitalization, by time since vaccination, (mRNA and ChAdOx1), ≥18-year-olds, British Columbia, and Quebec, Canada**

| | British Columbia, Canada | | | | Quebec, Canada | | | |
|---|---|---|---|---|---|---|---|---|
| | Sample sizes | | VE (95% CI) | | Sample sizes | | VE (95% CI) | |
| | Case N | Control N | Crude | Adjusted[1] | Case N | Control N | Crude | Adjusted[1] |
| **Any two mRNA vaccines, Infection** | | | | | | | | |
| 0-13 days (0-1 week) | 940 | 22615 | 83 (82, 84) | 75 (73, 77) | 745 | 64574 | 78 (77, 80) | 69 (67, 71) |
| 14-27 days (2-3 weeks) | 378 | 28148 | 95 (94, 95) | 94 (93, 95) | 324 | 72539 | 92 (91, 93) | 91 (90, 92) |
| 28-55 days (4-7 weeks, 2nd month) | 1614 | 70047 | 91 (90, 91) | 92 (92, 92) | 1234 | 162071 | 86 (85, 87) | 89 (89, 90) |
| 56-83 days (8-11 weeks, 3rd month) | 2261 | 74147 | 88 (87, 88) | 90 (90, 91) | 1743 | 156632 | 79 (78, 80) | 87 (87, 88) |
| 84-111 days (12-15 weeks, 4th month) | 1428 | 38993 | 85 (84, 86) | 88 (87, 89) | 941 | 80130 | 78 (76, 79) | 85 (84, 86) |
| 112-139 days (16-19 weeks, 5th month) | 350 | 8837 | 84 (82, 86) | 85 (83, 87) | 277 | 26489 | 80 (78, 83) | 88 (86, 89) |
| 140-167 days (20-23 weeks, 6th month) | 88 | 2376 | 85 (81, 88) | 82 (77, 85) | 87 | 12433 | 87 (84, 89) | 91 (89, 93) |
| 168-195 days (24-27 weeks, 7th month) | 130 | 2618 | 80 (76, 83) | 84 (81, 86) | 19 | 1394 | 74 (60, 84) | 79 (67, 87) |
| 196+ days (28+ weeks, 8th+ month) | 346 | 5022 | 72 (69, 75) | 77 (74, 79) | 9 | 565 | 70 (42, 84) | 80 (61, 90) |
| Unvaccinated | 18809 | 76621 | Reference | | 11366 | 212918 | Reference | |
| **Any two mRNA vaccines, Hospitalization** | | | | | | | | |
| 0-13 days (0-1 week) | 20 | 22615 | 95 (92, 97) | 93 (89, 95) | 12 | 64574 | 94 (90, 97) | 92 (86, 96) |
| 14-27 days (2-3 weeks) | 8 | 28148 | 98 (97, 99) | 98 (96, 99) | 16 | 72539 | 93 (89, 96) | 93 (89, 96) |
| 28-55 days (4-7 weeks, 2nd month) | 26 | 70047 | 98 (97, 99) | 98 (98, 99) | 26 | 162071 | 95 (93, 97) | 97 (95, 98) |
| 56-83 days (8-11 weeks, 3rd month) | 55 | 74147 | 96 (95, 97) | 98 (97, 99) | 41 | 156632 | 92 (89, 94) | 97 (96, 98) |
| 84-111 days (12-15 weeks, 4th month) | 67 | 38993 | 90 (88, 92) | 97 (96, 97) | 48 | 80130 | 82 (76, 86) | 95 (94, 97) |
| 112-139 days (16-19 weeks, 5th month) | 14 | 8837 | 91 (85, 95) | 95 (92, 97) | 9 | 26489 | 90 (80, 95) | 96 (92, 98) |
| 140-167 days (20-23 weeks, 6th month) | 3 | 2376 | 93 (78, 98) | 93 (78, 98) | 2 | 12433 | 95 (80, 99) | 97 (89, 99) |
| 168+ days (24+ weeks, 7th+ month) | 5 | 7640 | 96 (91, 98) | 97 (94, 99) | 1 | 1959 | 84 (-10, 98) | 95 (67, 99) |
| Unvaccinated | 1365 | 76621 | Reference | | 702 | 212918 | Reference | |
| **Two ChAdOx1 vaccines, Infection** | | | | | | | | |
| 0-13 days (0-1 week) | 7 | 558 | 95 (89, 98) | 80 (57, 91) | 6 | 1924 | 94 (87, 97) | 74 (42, 88) |
| 14-27 days (2-3 weeks) | 8 | 548 | 94 (88, 97) | 79 (58, 90) | 4 | 2185 | 97 (91, 99) | 86 (62, 95) |
| 28-55 days (4-7 weeks, 2nd month) | 77 | 1504 | 79 (74, 83) | 74 (67, 79) | 17 | 3970 | 92 (87, 95) | 82 (70, 89) |
| 56-83 days (8-11 weeks, 3rd month) | 248 | 2826 | 64 (59, 69) | 73 (69, 76) | 62 | 4313 | 73 (65, 79) | 74 (67, 80) |
| 84+ days (12+ weeks, 4th+ month) | 312 | 3199 | 60 (55, 65) | 69 (64, 72) | 147 | 6434 | 57 (50, 64) | 71 (65, 75) |
| Unvaccinated | 18809 | 76621 | Reference | | 11366 | 212918 | Reference | |
| **Two ChAdOx1 vaccines, Hospitalization** | | | | | | | | |
| 0-13 days (0-1 week) | 0 | 558 | NE | NE | 1 | 1924 | 84 (-12, 98) | 78 (-59, 97) |
| 14-27 days (2-3 weeks) | 0 | 548 | NE | NE | 0 | 2185 | NE | NE |
| 28-55 days (4-7 weeks, 2nd month) | 3 | 1504 | 89 (65, 96) | 88 (62, 96) | 0 | 3970 | NE | NE |
| 56-83 days (8-11 weeks, 3rd month) | 7 | 2826 | 86 (71, 93) | 93 (86, 97) | 3 | 4313 | 79 (34, 93) | 94 (81, 98) |
| 84+ days (12+ weeks, 4th+ month) | 7 | 3199 | 88 (74, 94) | 95 (89, 98) | 10 | 6434 | 53 (12, 75) | 93 (87, 96) |
| Unvaccinated | 1365 | 76621 | Reference | | 702 | 212918 | Reference | |

VE=vaccine effectiveness; 95%CI=95% confidence interval; NE=Not estimable

[1] Unless otherwise specified, all VE estimates adjusted for age group (18-49, 50-69, 70-79, ≥80 years); sex (male, female); individual epidemiological week of the analysis period (weeks 22-39, categorical); and region of the province (5 in each province).

**Supplementary Table 11. Two-dose vaccine effectiveness (VE) against infection and hospitalization, by time since vaccination, (and mRNA vaccine type), ≥18-year-olds, British Columbia, and Quebec, Canada**

| | British Columbia, Canada | | | | Quebec, Canada | | | |
|---|---|---|---|---|---|---|---|---|
| | Sample sizes | | VE (95% CI) | | Sample sizes | | VE (95% CI) | |
| | Case N | Control N | Crude | Adjusted[1] | Case N | Control N | Crude | Adjusted[1] |
| **Two BNT162b2 (Pfizer-BioNTech), Infection** | | | | | | | | |
| 0-13 days (0-1 week) | 717 | 16156 | 82 (80, 83) | 72 (70, 74) | 616 | 48138 | 76 (74, 78) | 66 (63, 68) |
| 14-27 days (2-3 weeks) | 302 | 20709 | 94 (93, 95) | 93 (93, 94) | 256 | 55110 | 91 (90, 92) | 91 (90, 92) |
| 28-55 days (4-7 weeks, 2nd month) | 1268 | 51929 | 90 (89, 91) | 92 (91, 92) | 989 | 124151 | 85 (84, 86) | 89 (88, 90) |
| 56-83 days (8-11 weeks, 3rd month) | 1667 | 53607 | 87 (87, 88) | 90 (89, 90) | 1358 | 116747 | 78 (77, 79) | 87 (86, 88) |
| 84-111 days (12-15 weeks, 4th month) | 1009 | 27629 | 85 (84, 86) | 88 (87, 89) | 767 | 60368 | 76 (74, 78) | 84 (83, 85) |
| 112-139 days (16-19 weeks, 5th month) | 234 | 6906 | 86 (84, 88) | 86 (84, 88) | 238 | 20682 | 78 (75, 81) | 88 (86, 89) |
| 140-167 days (20-23 weeks, 6th month) | 62 | 1811 | 86 (82, 89) | 81 (75, 85) | 76 | 9721 | 85 (82, 88) | 91 (89, 93) |
| 168-195 days (24-27 weeks, 7th month) | 104 | 2069 | 80 (75, 83) | 83 (79, 86) | 16 | 782 | 62 (37, 77) | 75 (59, 85) |
| 196+ days (28+ weeks, 8th+ month) | 230 | 4171 | 78 (74, 80) | 81 (78, 83) | 7 | 407 | 68 (32, 85) | 80 (59, 91) |
| Unvaccinated | 18809 | 76621 | Reference | | 11366 | 212918 | Reference | |
| **Two BNT162b2 (Pfizer-BioNTech), Hospitalization** | | | | | | | | |
| 0-13 days (0-1 week) | 14 | 16156 | 95 (92, 97) | 93 (87, 96) | 10 | 48138 | 94 (88, 97) | 92 (84, 95) |
| 14-27 days (2-3 weeks) | 6 | 20709 | 98 (96, 99) | 98 (95, 99) | 11 | 55110 | 94 (89, 97) | 94 (89, 97) |
| 28-55 days (4-7 weeks, 2nd month) | 18 | 51929 | 98 (97, 99) | 98 (98, 99) | 20 | 124151 | 95 (92, 97) | 97 (95, 98) |
| 56-83 days (8-11 weeks, 3rd month) | 40 | 53607 | 96 (94, 97) | 98 (97, 99) | 29 | 116747 | 92 (89, 95) | 98 (96, 98) |
| 84-111 days (12-15 weeks, 4th month) | 47 | 27629 | 90 (87, 93) | 97 (96, 98) | 45 | 60368 | 77 (69, 83) | 95 (93, 96) |
| 112-139 days (16-19 weeks, 5th month) | 6 | 6906 | 95 (89, 98) | 97 (94, 99) | 7 | 20682 | 90 (78, 95) | 96 (92, 98) |
| 140-167 days (20-23 weeks, 6th month) | 2 | 1811 | 94 (75, 98) | 92 (69, 98) | 1 | 9721 | 97 (78, 100) | 98 (89, 100) |
| 168+ days (24+ weeks, 7th+ month) | 3 | 6240 | 97 (92, 99) | 98 (94, 99) | 0 | 1189 | NE | NE |
| Unvaccinated | 1365 | 76621 | Reference | | 702 | 212918 | Reference | |
| **Two mRNA-1273 (Moderna), Infection** | | | | | | | | |
| 0-13 days (0-1 week) | 199 | 4707 | 83 (80, 85) | 81 (77, 83) | 118 | 14125 | 84 (81, 87) | 79 (75, 82) |
| 14-27 days (2-3 weeks) | 65 | 5512 | 95 (94, 96) | 95 (94, 96) | 63 | 15315 | 92 (90, 94) | 92 (90, 94) |
| 28-55 days (4-7 weeks, 2nd month) | 248 | 13052 | 92 (91, 93) | 94 (93, 94) | 212 | 33187 | 88 (86, 90) | 91 (90, 92) |
| 56-83 days (8-11 weeks, 3rd month) | 356 | 12943 | 89 (88, 90) | 91 (90, 92) | 312 | 34862 | 83 (81, 85) | 90 (89, 91) |
| 84-111 days (12-15 weeks, 4th month) | 261 | 6685 | 84 (82, 86) | 88 (87, 90) | 160 | 17915 | 83 (80, 86) | 87 (85, 89) |
| 112-139 days (16-19 weeks, 5th month) | 112 | 1749 | 74 (68, 78) | 82 (78, 85) | 35 | 5286 | 88 (83, 91) | 87 (82, 91) |
| 140-167 days (20-23 weeks, 6th month) | 25 | 558 | 82 (73, 88) | 84 (76, 89) | 9 | 2537 | 84 (81, 87) | 79 (75, 82) |
| 168+ days (24+ weeks, 7th+ month) | 334 | 6240 | 58 (51, 65) | 71 (65, 75) | 5 | 759 | 88 (70, 95) | 86 (67, 94) |
| Unvaccinated | 18809 | 76621 | Reference | | 11366 | 212918 | Reference | |
| **Two mRNA-1273 (Moderna), Hospitalization** | | | | | | | | |
| 0-13 days (0-1 week) | 6 | 4707 | 93 (84, 97) | 91 (81, 96) | 1 | 14125 | 98 (85, 100) | 97 (79, 100) |
| 14-27 days (2-3 weeks) | 1 | 5512 | 99 (93, 100) | 99 (92, 100) | 3 | 15315 | 94 (82, 98) | 94 (81, 98) |
| 28-55 days (4-7 weeks, 2nd month) | 4 | 13052 | 98 (95, 99) | 99 (96, 100) | 5 | 33187 | 95 (89, 98) | 97 (93, 99) |
| 56-83 days (8-11 weeks, 3rd month) | 9 | 12943 | 96 (92, 98) | 98 (96, 99) | 9 | 34862 | 92 (85, 96) | 97 (94, 98) |
| 84-111 days (12-15 weeks, 4th month) | 13 | 6685 | 89 (81, 94) | 96 (93, 98) | 3 | 17915 | 95 (84, 98) | 98 (94, 99) |
| 112-139 days (16-19 weeks, 5th month) | 8 | 1749 | 74 (48, 87) | 90 (79, 95) | 2 | 5286 | 89 (54, 97) | 93 (72, 98) |
| 140-167 days (20-23 weeks, 6th month) | 1 | 558 | 90 (28, 99) | 94 (56, 99) | 1 | 2537 | 88 (15, 98) | 92 (44, 99) |
| 168+ days (24+ weeks, 7th+ month) | 3 | 6240 | 92 (68, 98) | 96 (83, 99) | 1 | 759 | NE | NE |
| Unvaccinated | 1365 | 76621 | Reference | | 702 | 212918 | Reference | |

VE=vaccine effectiveness; 95%CI=95% confidence interval; NE=Not estimable

[1] Unless otherwise specified, all VE estimates adjusted for age group (18-49, 50-69, 70-79, ≥80 years); sex (male, female); individual epidemiological week of the analysis period (weeks 22-39, categorical); and region of the province (5 in each province).

**Supplementary Table 12. Two-dose vaccine effectiveness (VE) against infection and hospitalization, by time since vaccination, (mixed mRNA and mRNA/ChAdOx1), ≥18-year-olds, British Columbia, and Quebec, Canada**

| | British Columbia, Canada | | | | Quebec, Canada | | | |
|---|---|---|---|---|---|---|---|---|
| | Sample sizes | | VE (95% CI) | | Sample sizes | | VE (95% CI) | |
| | Case N | Control N | Crude | **Adjusted**[1] | Case N | Control N | Crude | **Adjusted**[1] |
| **Two mixed mRNA, Infection** | | | | | | | | |
| 0-13 days (0-1 week) | 24 | 1750 | 94 (92, 96) | 83 (74, 89) | 11 | 2311 | 91 (84, 95) | 79 (61, 88) |
| 14-27 days (2-3 weeks) | 11 | 1925 | 98 (96, 99) | 95 (91, 97) | 5 | 2114 | 96 (89, 98) | 93 (84, 97) |
| 28-55 days (4-7 weeks, 2nd month) | 98 | 5064 | 92 (90, 94) | 93 (91, 94) | 33 | 4733 | 87 (82, 91) | 90 (86, 93) |
| 56-83 days (8-11 weeks, 3rd month) | 238 | 7594 | 87 (85, 89) | 91 (89, 92) | 73 | 5023 | 73 (66, 78) | 85 (81, 88) |
| 84-111 days (12-15 weeks, 4th month) | 157 | 4677 | 86 (84, 88) | 88 (86, 90) | 14 | 1847 | 86 (76, 92) | 90 (83, 94) |
| 112-139 days (16-19 weeks, 5th month) | 4 | 182 | 91 (76, 97) | 93 (80, 97) | 4 | 521 | 86 (61, 95) | 91 (76, 97) |
| 140-167 days (20-23 weeks, 6th month) | 1 | 7 | NE | NE | 2 | 175 | 79 (14, 95) | 85 (40, 96) |
| 168-195 days (24-27 weeks, 7th month) | 0 | 1 | NE | NE | 0 | 8 | NE | NE |
| 196+ days (28+ weeks, 8th+ month) | 0 | 8 | NE | NE | 0 | 3 | NE | NE |
| Unvaccinated | 18809 | 76621 | Reference | | 11366 | 212918 | Reference | |
| **Two mixed mRNA, Hospitalization** | | | | | | | | |
| 0-13 days (0-1 week) | 0 | 1750 | NE | NE | 1 | 2311 | 87 (7, 98) | NE |
| 14-27 days (2-3 weeks) | 1 | 1925 | 97 (79, 100) | 95 (64, 99) | 2 | 2114 | NE | NE |
| 28-55 days (4-7 weeks, 2nd month) | 4 | 5064 | 96 (88, 98) | 97 (92, 100) | 1 | 4733 | 94 (54, 99) | 96 (70, 99) |
| 56-83 days (8-11 weeks, 3rd month) | 6 | 7594 | 96 (90, 98) | 98 (96, 99) | 3 | 5023 | 82 (44, 94) | 93 (78, 98) |
| 84-111 days (12-15 weeks, 4th month) | 7 | 4677 | 92 (82, 96) | 97 (94, 99) | 0 | 1847 | NE | NE |
| 112-139 days (16-19 weeks, 5th month) | 0 | 182 | NE | NE | 0 | 521 | NE | NE |
| 140-167 days (20-23 weeks, 6th month) | 0 | 7 | NE | NE | 0 | 175 | NE | NE |
| 168+ days (24+ weeks, 7th+ month) | 0 | 1 | NE | NE | 0 | 8 | NE | NE |
| Unvaccinated | 1365 | 76621 | Reference | | 702 | 212918 | Reference | |
| **Two mixed ChadOx1 + mRNA, Infection** | | | | | | | | |
| 0-13 days (0-1 week) | 39 | 950 | 83 (77, 88) | 45 (23, 60) | 11 | 3382 | 94 (89, 97) | 78 (60, 88) |
| 14-27 days (2-3 weeks) | 11 | 1115 | 96 (93, 98) | 93 (87, 96) | 7 | 3363 | 96 (92, 98) | 91 (82, 96) |
| 28-55 days (4-7 weeks, 2nd month) | 102 | 3756 | 89 (87, 91) | 91 (89, 92) | 48 | 7334 | 88 (84, 91) | 88 (84, 91) |
| 56-83 days (8-11 weeks, 3rd month) | 176 | 5833 | 88 (86, 89) | 91 (89, 92) | 18 | 9694 | 77 (73, 81) | 87 (84, 89) |
| 84-111 days (12-15 weeks, 4th month) | 114 | 3335 | 86 (83, 88) | 88 (86, 90) | 68 | 5230 | 76 (69, 81) | 84 (79, 87) |
| 112-139 days (16-19 weeks, 5th month) | 1 | 45 | 91 (34, 99) | 92 (44, 99) | 0 | 101 | NE | NE |
| 140-167 days (20-23 weeks, 6th month) | 0 | 0 | NE | NE | 1 | 5 | NE | NE |
| 168+ days (24+ weeks, 7th+ month) | 0 | 0 | NE | NE | 0 | 1 | NE | NE |
| Unvaccinated | 18809 | 76621 | Reference | | 11366 | 212918 | Reference | |
| **Two mixed ChadOx1 + mRNA, Hospitalization** | | | | | | | | |
| 0-13 days (0-1 week) | 0 | 950 | NE | NE | 0 | 3382 | NE | NE |
| 14-27 days (2-3 weeks) | 1 | 1115 | 95 (64, 99) | 91 (34, 99) | 0 | 3363 | NE | NE |
| 28-55 days (4-7 weeks, 2nd month) | 1 | 3756 | 99 (89, 100) | 99 (91, 100) | 1 | 7334 | 96 (71, 99) | 98 (87, 100) |
| 56-83 days (8-11 weeks, 3rd month) | 0 | 5833 | NE | NE | 2 | 9694 | 94 (75, 98) | 99 (94, 100) |
| 84-111 days (12-15 weeks, 4th month) | 0 | 3335 | NE | NE | 4 | 5230 | 77 (38, 91) | 94 (84, 98) |
| 112-139 days (16-19 weeks, 5th month) | 0 | 45 | NE | NE | 0 | 101 | NE | NE |
| 140-167 days (20-23 weeks, 6th month) | 0 | 0 | NE | NE | 0 | 5 | NE | NE |
| 168+ days (24+ weeks, 7th+ month) | 0 | 0 | NE | NE | 0 | 1 | NE | NE |
| Unvaccinated | 1365 | 76621 | Reference | | 702 | 212918 | 212918 | |

VE=vaccine effectiveness; 95%CI=95% confidence interval; NE=Not estimable

[1] Unless otherwise specified, all VE estimates adjusted for age group (18-49, 50-69, 70-79, ≥80 years); sex (male, female); individual epidemiological week of the analysis period (weeks 22-39, categorical); and region of the province (5 in each province).

**Supplementary Table 13. Two-dose vaccine effectiveness (VE) against Delta-specific infection and hospitalization, by time since vaccination, (mRNA and ChAdOx1), ≥18-year-olds, British Columbia, and Quebec, Canada**

| | British Columbia, Canada | | | | Quebec, Canada | | | |
|---|---|---|---|---|---|---|---|---|
| | Sample sizes | | VE (95% CI) | | Sample sizes | | VE (95% CI) | |
| | Case N | Control N | Crude | Adjusted[1] | Case N | Control N | Crude | Adjusted[1] |
| **Any two mRNA vaccines, Delta infection** | | | | | | | | |
| 0-13 days (0-1 week) | 667 | 22615 | 80 (79, 82) | 74 (72, 76) | 322 | 64574 | 83 (81, 85) | 73 (70, 76) |
| 14-27 days (2-3 weeks) | 260 | 28148 | 94 (93, 95) | 95 (94, 95) | 162 | 72539 | 93 (91, 94) | 92 (91, 93) |
| 28-55 days (4-7 weeks, 2nd month) | 1113 | 70047 | 89 (89, 90) | 93 (92, 93) | 744 | 162071 | 85 (83, 86) | 90 (90, 91) |
| 56-83 days (8-11 weeks, 3rd month) | 1136 | 74147 | 90 (89, 90) | 91 (90, 91) | 1223 | 156632 | 74 (72, 75) | 88 (88, 89) |
| 84-111 days (12-15 weeks, 4th month) | 464 | 38993 | 92 (91, 93) | 89 (88, 90) | 682 | 80130 | 71 (69, 74) | 86 (85, 87) |
| 112-139 days (16-19 weeks, 5th month) | 85 | 8837 | 94 (92, 95) | 85 (81, 88) | 199 | 26489 | 75 (71, 78) | 89 (87, 90) |
| 140-167 days (20-23 weeks, 6th month) | 41 | 2376 | 89 (84, 92) | 80 (73, 86) | 61 | 12433 | 84 (79, 87) | 92 (90, 94) |
| 168-195 days (24-27 weeks, 7th month) | 110 | 2618 | 72 (66, 77) | 83 (80, 86) | 14 | 1394 | 66 (43, 80) | 79 (65, 88) |
| 196+ days (28+ weeks, 8th+ month) | 111 | 5022 | 85 (82, 88) | 79 (75, 83) | 8 | 565 | 53 (5, 76) | 78 (55, 89) |
| Unvaccinated | 11500 | 76621 | Reference | | 6349 | 212918 | Reference | |
| **Any two mRNA vaccines, Delta Hospitalization[2]** | | | | | | | | |
| 0-13 days (0-1 week) | 10 | 22615 | 96 (93, 98) | 92 (86, 96) | 5 | 19611 | 95 (87, 98) | 93 (83, 97) |
| 14-27 days (2-3 weeks) | 5 | 28148 | 98 (96, 99) | 98 (95, 99) | 7 | 34394 | 96 (91, 98) | 94 (88, 97) |
| 28-55 days (4-7 weeks, 2nd month) | 17 | 70047 | 98 (97, 99) | 99 (98, 99) | 9 | 118284 | 98 (97, 99) | 98 (97, 99) |
| 56-83 days (8-11 weeks, 3rd month) | 46 | 74147 | 95 (93, 96) | 98 (97, 98) | 25 | 142975 | 96 (94, 97) | 98 (97, 99) |
| 84-111 days (12-15 weeks, 4th month) | 30 | 38993 | 93 (90, 95) | 97 (96, 98) | 36 | 76325 | 90 (86, 93) | 96 (94, 97) |
| 112-139 days (16-19 weeks, 5th month) | 7 | 8837 | 93 (85, 97) | 93 (85, 97) | 7 | 25841 | 94 (88, 97) | 96 (91, 98) |
| 140-167 days (20-23 weeks, 6th month) | 1 | 2376 | 96 (74, 99) | 95 (65, 99) | 1 | 12199 | 98 (87, 100) | 98 (89, 100) |
| 168+ days (24+ weeks, 7th+ month) | 2 | 7640 | 98 (91, 99) | 98 (93, 100) | 0 | 1929 | NE | NE |
| Unvaccinated | 873 | 76621 | Reference | | 438 | 94524 | Reference | |
| **Two ChAdOx1 vaccines, Delta infection** | | | | | | | | |
| 0-13 days (0-1 week) | 2 | 558 | 98 (90, 99) | 76 (1, 94) | 0 | 1924 | NE | NE |
| 14-27 days (2-3 weeks) | 4 | 548 | 95 (87, 98) | 77 (38, 92) | 2 | 2185 | 97 (88, 99) | 74 (-7, 94) |
| 28-55 days (4-7 weeks, 2nd month) | 74 | 1504 | 67 (59, 74) | 68 (60, 75) | 9 | 3970 | 92 (85, 96) | 78 (58, 89) |
| 56-83 days (8-11 weeks, 3rd month) | 199 | 2826 | 53 (46, 59) | 72 (67, 76) | 36 | 4313 | 72 (61, 80) | 76 (67, 83) |
| 84+ days (12+ weeks, 4th+ month) | 98 | 3199 | 80 (75, 83) | 65 (57, 72) | 115 | 6434 | 40 (28, 50) | 72 (66, 77) |
| Unvaccinated | 11500 | 76621 | Reference | | 6349 | 212918 | Reference | |
| **Two ChAdOx1 vaccines, Delta Hospitalization[2]** | | | | | | | | |
| 0-13 days (0-1 week) | 0 | 558 | NE | NE | 0 | 68 | NE | NE |
| 14-27 days (2-3 weeks) | 0 | 548 | NE | NE | 0 | 149 | NE | NE |
| 28-55 days (4-7 weeks, 2nd month) | 3 | 1504 | 82 (46, 94) | 84 (51, 95) | 0 | 989 | NE | NE |
| 56-83 days (8-11 weeks, 3rd month) | 6 | 2826 | 81 (58, 92) | 93 (84, 97) | 1 | 3962 | 95 (61, 99) | 97 (78, 100) |
| 84+ days (12+ weeks, 4th+ month) | 5 | 3199 | 86 (67, 94) | 92 (81, 97) | 8 | 6432 | 73 (46, 87) | 93 (86, 97) |
| Unvaccinated | 873 | 76621 | Reference | | 438 | 94524 | Reference | |

VE=vaccine effectiveness; 95%CI=95% confidence interval; NE=Not estimable

[1] Unless otherwise specified, all VE estimates adjusted for age group (18-49, 50-69, 70-79, ≥80 years); sex (male, female); individual epidemiological week of the analysis period (weeks 22-39, categorical); and region of the province (5 in each province).

[2] In Quebec, VE against hospitalizations due to Delta variant assessed only between weeks 31-39 because no hospitalized Delta variant cases identified prior to that period.

**Supplementary Table 14. Two-dose vaccine effectiveness (VE) against Delta-specific infection and hospitalization, by time since vaccination, (and mRNA vaccine type), ≥18-year-olds, British Columbia, and Quebec, Canada**

| | British Columbia, Canada | | | | Quebec, Canada | | | |
|---|---|---|---|---|---|---|---|---|
| | Sample sizes | | VE (95% CI) | | Sample sizes | | VE (95% CI) | |
| | Case N | Control N | Crude | Adjusted[1] | Case N | Control N | Crude | Adjusted[1] |
| **Two BNT162b2 (Pfizer-BioNTech), Infection** | | | | | | | | |
| 0-13 days (0-1 week) | 517 | 16156 | 79 (77, 81) | 71 (69, 74) | 260 | 48138 | 82 (79, 84) | 70 (66, 74) |
| 14-27 days (2-3 weeks) | 215 | 20709 | 93 (92, 94) | 94 (93, 95) | 131 | 55110 | 92 (91, 93) | 92 (90, 93) |
| 28-55 days (4-7 weeks, 2nd month) | 883 | 51929 | 89 (88, 89) | 92 (92, 93) | 598 | 124151 | 84 (82, 85) | 90 (89, 91) |
| 56-83 days (8-11 weeks, 3rd month) | 819 | 53607 | 90 (89, 91) | 90 (90, 91) | 956 | 116747 | 73 (71, 74) | 88 (87, 89) |
| 84-111 days (12-15 weeks, 4th month) | 330 | 27629 | 92 (91, 93) | 89 (88, 90) | 562 | 60368 | 69 (66, 71) | 85 (84, 86) |
| 112-139 days (16-19 weeks, 5th month) | 48 | 6906 | 95 (94, 97) | 86 (81, 89) | 171 | 20682 | 72 (68, 76) | 89 (87, 90) |
| 140-167 days (20-23 weeks, 6th month) | 34 | 1811 | 87 (82, 91) | 77 (67, 84) | 53 | 9721 | 82 (76, 86) | 92 (89, 94) |
| 168-195 days (24-27 weeks, 7th month) | 85 | 2069 | 73 (66, 78) | 83 (79, 86) | 11 | 782 | 53 (14, 74) | 76 (57, 87) |
| 196+ days (28+ weeks, 8th+ month) | 87 | 4171 | 86 (83, 89) | 80 (76, 84) | 7 | 407 | 42 (-22, 73) | 76 (48, 88) |
| Unvaccinated | 11500 | 76621 | Reference | | 6349 | 212918 | Reference | |
| **Two BNT162b2 (Pfizer-BioNTech), Hospitalization[2]** | | | | | | | | |
| 0-13 days (0-1 week) | 8 | 16156 | 96 (91, 98) | 91 (82, 95) | 3 | 14594 | 96 (86, 99) | 94 (82, 98) |
| 14-27 days (2-3 weeks) | 4 | 20709 | 98 (95, 99) | 98 (94, 99) | 6 | 25973 | 95 (89, 98) | 94 (86, 97) |
| 28-55 days (4-7 weeks, 2nd month) | 12 | 51929 | 98 (96, 99) | 99 (98, 99) | 6 | 88853 | 99 (97, 99) | 99 (97, 99) |
| 56-83 days (8-11 weeks, 3rd month) | 34 | 53607 | 94 (92, 96) | 98 (97, 98) | 18 | 106378 | 96 (94, 98) | 98 (97, 99) |
| 84-111 days (12-15 weeks, 4th month) | 22 | 27629 | 93 (89, 95) | 97 (96, 98) | 34 | 57834 | 87 (82, 91) | 95 (93, 97) |
| 112-139 days (16-19 weeks, 5th month) | 1 | 6906 | 99 (91, 100) | 98 (88, 100) | 5 | 20332 | 95 (87, 98) | 97 (92, 99) |
| 140-167 days (20-23 weeks, 6th month) | 1 | 1811 | 95 (66, 99) | 92 (41, 99) | 1 | 9578 | 98 (84, 100) | 98 (87, 100) |
| 168+ days (24+ weeks, 7th+ month) | 2 | 6240 | 97 (89, 99) | 98 (91, 99) | 0 | 1163 | NE | NE |
| Unvaccinated | 873 | 76621 | Reference | | 438 | 94524 | Reference | |
| **Two mRNA-1273 (Moderna), Infection** | | | | | | | | |
| 0-13 days (0-1 week) | 140 | 4707 | 80 (77, 83) | 80 (76, 83) | 55 | 14125 | 87 (83, 90) | 81 (76, 86) |
| 14-27 days (2-3 weeks) | 40 | 5512 | 95 (93, 96) | 96 (94, 97) | 29 | 15315 | 94 (91, 96) | 94 (91, 96) |
| 28-55 days (4-7 weeks, 2nd month) | 150 | 13052 | 92 (91, 93) | 94 (93, 95) | 123 | 33187 | 88 (85, 90) | 92 (91, 93) |
| 56-83 days (8-11 weeks, 3rd month) | 190 | 12943 | 90 (89, 92) | 91 (90, 93) | 216 | 34862 | 79 (76, 82) | 91 (89, 92) |
| 84-111 days (12-15 weeks, 4th month) | 100 | 6685 | 90 (88, 92) | 88 (86, 90) | 112 | 17915 | 79 (75, 83) | 88 (86, 90) |
| 112-139 days (16-19 weeks, 5th month) | 37 | 1749 | 86 (80, 90) | 83 (76, 88) | 25 | 5286 | 84 (76, 89) | 87 (81, 91) |
| 140-167 days (20-23 weeks, 6th month) | 7 | 558 | 92 (82, 96) | 89 (76, 95) | 8 | 2537 | 89 (79, 95) | 91 (81, 95) |
| 168+ days (24+ weeks, 7th+ month) | 49 | 1390 | 77 (69, 82) | 80 (73, 85) | 4 | 759 | 82 (53, 93) | 85 (61, 95) |
| Unvaccinated | 11500 | 76621 | Reference | | 6349 | 212918 | Reference | |
| **Two mRNA-1273 (Moderna), Hospitalization[2]** | | | | | | | | |
| 0-13 days (0-1 week) | 2 | 4707 | 96 (85, 99) | 95 (80, 99) | 1 | 4689 | 95 (67, 99) | 94 (59, 99) |
| 14-27 days (2-3 weeks) | 1 | 5512 | 98 (89, 100) | 98 (89, 100) | 0 | 7620 | NE | NE |
| 28-55 days (4-7 weeks, 2nd month) | 2 | 13052 | 99 (95, 100) | 99 (96, 100) | 2 | 25763 | 98 (93, 100) | 98 (93, 100) |
| 56-83 days (8-11 weeks, 3rd month) | 8 | 12943 | 95 (89, 97) | 98 (95, 99) | 5 | 31846 | 97 (92, 99) | 98 (95, 99) |
| 84-111 days (12-15 weeks, 4th month) | 7 | 6685 | 91 (81, 96) | 96 (92, 98) | 2 | 16686 | 97 (90, 99) | 99 (94, 100) |
| 112-139 days (16-19 weeks, 5th month) | 6 | 1749 | 70 (33, 87) | 84 (63, 93) | 2 | 4990 | 91 (65, 98) | 92 (66, 98) |
| 140-167 days (20-23 weeks, 6th month) | 0 | 558 | NE | NE | 0 | 2448 | NE | NE |
| 168+ days (24+ weeks, 7th+ month) | 0 | 1390 | NE | NE | 0 | 755 | NE | NE |
| Unvaccinated | 873 | 76621 | Reference | | 438 | 94524 | Reference | |

VE=vaccine effectiveness; 95%CI=95% confidence interval; NE=Not estimable

[1] Unless otherwise specified, all VE estimates adjusted for age group (18-49, 50-69, 70-79, ≥80 years); sex (male, female); individual epidemiological week of the analysis period (weeks 22-39, categorical); and region of the province (5 in each province).
[2] In Quebec, VE against hospitalizations due to Delta variant assessed only between weeks 31-39 because no hospitalized Delta variant cases identified prior to that period.

**Supplementary Table 15. Two-dose vaccine effectiveness (VE) against Delta-specific infection and hospitalization, by time since vaccination, (mixed mRNA and mRNA/ChAdOx1), ≥18-year-olds, British Columbia, and Quebec, Canada**

| | British Columbia, Canada | | | | Quebec, Canada | | | |
|---|---|---|---|---|---|---|---|---|
| | Sample sizes | | VE (95% CI) | | Sample sizes | | VE (95% CI) | |
| | Case N | Control N | Crude | Adjusted[1] | Case N | Control N | Crude | Adjusted[1] |
| **Two mixed mRNA, Delta Infection** | | | | | | | | |
| 0-13 days (0-1 week) | 10 | 1750 | 96 (93, 98) | 85 (73, 92) | 7 | 2311 | 90 (79, 95) | 60 (16, 81) |
| 14-27 days (2-3 weeks) | 5 | 1925 | 98 (96, 99) | 97 (92, 99) | 2 | 2114 | 97 (87, 99) | 94 (75, 98) |
| 28-55 days (4-7 weeks, 2nd month) | 80 | 5064 | 89 (90, 92) | 93 (91, 94) | 23 | 4733 | 84 (75, 89) | 89 (84, 93) |
| 56-83 days (8-11 weeks, 3rd month) | 127 | 7594 | 89 (87, 91) | 92 (90, 93) | 51 | 5023 | 66 (55, 74) | 87 (82, 90) |
| 84-111 days (12-15 weeks, 4th month) | 33 | 4677 | 95 (93, 97) | 88 (82, 91) | 8 | 1847 | 85 (71, 93) | 93 (86, 97) |
| 112-139 days (16-19 weeks, 5th month) | 0 | 182 | NE | NE | 3 | 521 | 81 (40, 94) | 91 (73, 97) |
| 140-167 days (20-23 weeks, 6th month) | 0 | 7 | NE | NE | 0 | 175 | NE | NE |
| 168-195 days (24-27 weeks, 7th month) | 0 | 1 | NE | NE | 0 | 8 | NE | NE |
| 196+ days (28+ weeks, 8th+ month) | | 8 | NE | NE | 0 | 3 | NE | NE |
| Unvaccinated | 11500 | 76621 | Reference | | 6349 | 212918 | Reference | |
| **Two mixed mRNA, Delta Hospitalization [2]** | | | | | | | | |
| 0-13 days (0-1 week) | 0 | 1750 | NE | NE | 1 | 328 | NE | NE |
| 14-27 days (2-3 weeks) | 0 | 1925 | NE | NE | 1 | 801 | NE | NE |
| 28-55 days (4-7 weeks, 2nd month) | 3 | 5064 | 95 (84, 98) | 96 (88, 99) | 1 | 3668 | 94 (58, 99) | 94 (55, 99) |
| 56-83 days (8-11 weeks, 3rd month) | 4 | 7594 | 95 (88, 98) | 98 (94, 99) | 2 | 4751 | 91 (64, 98) | 94 (77, 99) |
| 84-111 days (12-15 weeks, 4th month) | 1 | 4677 | 98 (87, 100) | 98 (85, 100) | 0 | 1805 | NE | NE |
| 112-139 days (16-19 weeks, 5th month) | 0 | 182 | NE | NE | 0 | 519 | NE | NE |
| 140-167 days (20-23 weeks, 6th month) | 0 | 7 | NE | NE | 0 | 173 | NE | NE |
| 168+ days (24+ weeks, 7th+ month) | 0 | 1 | NE | NE | 0 | 11 | NE | NE |
| Unvaccinated | 873 | 76621 | Reference | | 438 | 94524 | Reference | |
| **Two mixed ChadOx1 + mRNA, Delta Infection** | | | | | | | | |
| 0-13 days (0-1 week) | 9 | 950 | 94 (88, 97) | 73 (48, 86) | 2 | 3382 | 98 (92, 100) | 86 (42, 96) |
| 14-27 days (2-3 weeks) | 8 | 1115 | 95 (90, 98) | 94 (87, 97) | 4 | 3363 | 96 (89, 99) | 87 (66, 95) |
| 28-55 days (4-7 weeks, 2nd month) | 89 | 3756 | 84 (80, 87) | 90 (88, 92) | 27 | 7334 | 88 (82, 92) | 89 (84, 92) |
| 56-83 days (8-11 weeks, 3rd month) | 97 | 5833 | 89 (86, 91) | 91 (89, 93) | 85 | 9694 | 71 (64, 76) | 88 (85, 90) |
| 84-111 days (12-15 weeks, 4th month) | 22 | 3335 | 96 (93, 97) | 85 (77, 90) | 50 | 5230 | 68 (58, 76) | 86 (81, 89) |
| 112-139 days (16-19 weeks, 5th month) | 0 | 45 | NE | NE | 0 | 101 | NE | NE |
| 140-167 days (20-23 weeks, 6th month) | 0 | 0 | NE | NE | 0 | 5 | NE | NE |
| 168+ days (24+ weeks, 7th+ month) | 0 | 0 | NE | NE | 0 | 1 | NE | NE |
| Unvaccinated | 11500 | 76621 | Reference | | 6349 | 212918 | Reference | |
| **Two mixed ChadOx1 + mRNA, Delta Hospitalization [2]** | | | | | | | | |
| 0-13 days (0-1 week) | 0 | 950 | NE | NE | 0 | 310 | NE | NE |
| 14-27 days (2-3 weeks) | 1 | 1115 | 92 (44, 99) | NE | 0 | 795 | NE | NE |
| 28-55 days (4-7 weeks, 2nd month) | 1 | 3756 | 98 (83, 100) | 99 (90, 100) | 1 | 5957 | 96 (74, 99) | 97 (81, 100) |
| 56-83 days (8-11 weeks, 3rd month) | 0 | 5833 | NE | NE | 1 | 9669 | 88 (62, 96) | 95 (85, 98) |
| 84-111 days (12-15 weeks, 4th month) | 0 | 3335 | NE | NE | 3 | 5228 | NE | NE |
| 112-139 days (16-19 weeks, 5th month) | 0 | 45 | NE | NE | 0 | 101 | NE | NE |
| 140-167 days (20-23 weeks, 6th month) | 0 | 0 | NE | NE | 0 | 5 | NE | NE |
| 168+ days (24+ weeks, 7th+ month) | 0 | 0 | NE | NE | 0 | 1 | NE | NE |
| Unvaccinated | 873 | 76621 | Reference | | 438 | 94524 | Reference | |

VE=vaccine effectiveness; 95%CI=95% confidence interval; NE=Not estimable

[1] Unless otherwise specified, all VE estimates adjusted for age group (18-49, 50-69, 70-79, ≥80 years); sex (male, female); individual epidemiological week of the analysis period (weeks 22-39, categorical); and region of the province (5 in each province).

[2] In Quebec, VE against hospitalizations due to Delta variant assessed only between weeks 31-39 because no hospitalized Delta variant cases identified prior to that period.

**Supplementary Table 16. Two-dose vaccine effectiveness against infection and hospitalization, by time since vaccination, (and mRNA vaccine type), ≥70-year-olds, British Columbia and Quebec, Canada**

| | British Columbia (BC), Canada | | | | Quebec, Canada | | | |
|---|---|---|---|---|---|---|---|---|
| | Sample sizes | | VE (95% CI) | | Sample sizes | | VE (95% CI) | |
| | Case N | Control N | Crude | Adjusted[1] | Case N | Control N | Crude | Adjusted[1] |
| **Two any mRNA, Infection** | | | | | | | | |
| 0-13 days (0-1 week) | 38 | 4364 | 94 (92, 96) | 81 (72, 87) | 36 | 12080 | 88 (83, 91) | 58 (37, 72) |
| 14-27 days (2-3 weeks) | 3 | 4438 | 100 (99, 100) | 99 (96, 100) | 19 | 13312 | 94 (91, 96) | 83 (72, 90) |
| 28-55 days (4-7 weeks, 2nd month) | 79 | 9556 | 95 (93, 96) | 92 (89, 93) | 74 | 26101 | 88 (85, 91) | 85 (80, 89) |
| 56-83 days (8-11 weeks, 3rd month) | 268 | 12123 | 86 (84, 88) | 92 (91, 93) | 158 | 26853 | 76 (71, 80) | 88 (85, 90) |
| 84-111 days (12-15 weeks, 4th month) | 311 | 10711 | 81 (79, 84) | 91 (90, 92) | 235 | 21499 | 56 (47, 63) | 83 (79, 86) |
| 112-139 days (16-19 weeks, 5th month) | 48 | 1453 | 79 (71, 84) | 89 (85, 92) | 30 | 3144 | 61 (43, 73) | 82 (74, 88) |
| 140+ (20+ weeks, 6th+ month) | 35 | 402 | 44 (20, 61) | 73 (60, 81) | 13 | 1205 | 56 (23, 75) | 76 (57, 87) |
| Unvaccinated | 770 | 4950 | Reference | | 257 | 10462 | Reference | |
| **Two any mRNA, Hospitalizations** | | | | | | | | |
| 0-13 days (0-1 week) | 6 | 4364 | 98 (95, 99) | 92 (80, 97) | 7 | 12080 | 95 (89, 98) | 77 (47, 90) |
| 14-27 days (2-3 weeks) | 1 | 4438 | 100 (97, 100) | 99 (91, 100) | 4 | 13312 | 97 (93, 99) | 92 (76, 97) |
| 28-55 days (4-7 weeks, 2nd month) | 11 | 9556 | 98 (96, 99) | 97 (94, 98) | 12 | 26101 | 96 (92, 98) | 94 (89, 97) |
| 56-83 days (8-11 weeks, 3rd month) | 32 | 12123 | 95 (93, 97) | 97 (96, 98) | 28 | 26853 | 90 (86, 94) | 95 (93, 97) |
| 84-111 days (12-15 weeks, 4th month) | 48 | 10711 | 92 (89, 94) | 96 (95, 97) | 36 | 21499 | 85 (78, 89) | 95 (92, 96) |
| 112-139 days (16-19 weeks, 5th month) | 10 | 1453 | 88 (77, 94) | 92 (85, 96) | 5 | 3144 | 85 (64, 94) | 94 (85, 98) |
| 140+ days (20+ weeks, 6th+ month) | 1 | 402 | 96 (69, 99) | 98 (83, 100) | 1 | 1205 | 92 (45, 99) | NE |
| Unvaccinated | 284 | 4950 | Reference | | 114 | 10462 | 10462 | |
| **Two BNT162b2 (Pfizer-BioNTech), Infection** | | | | | | | | |
| 0-13 days (0-1 week) | 30 | 3219 | 94 (91, 96) | 79 (68, 86) | 30 | 9593 | 87 (81, 91) | 56 (31, 72) |
| 14-27 days (2-3 weeks) | 2 | 3253 | 100 (98, 100) | 99 (94, 100) | 17 | 10725 | 94 (89, 96) | 80 (66, 88) |
| 28-55 days (4-7 weeks, 2nd month) | 60 | 7072 | 95 (93, 96) | 91 (88, 93) | 63 | 20789 | 88 (84, 91) | 84 (78, 88) |
| 56-83 days (8-11 weeks, 3rd month) | 213 | 8904 | 85 (82, 87) | 91 (89, 92) | 127 | 21438 | 76 (70, 81) | 88 (85, 90) |
| 84-111 days (12-15 weeks, 4th month) | 243 | 8057 | 81 (7, 83) | 91 (89, 92) | 207 | 17733 | 52 (43, 60) | 82 (77, 85) |
| 112-139 days (16-19 weeks, 5th month) | 32 | 1123 | 82 (74, 87) | 91 (86, 94) | 27 | 2229 | 51 (27, 67) | 80 (69, 87) |
| 140+ (20+ weeks, 6th+ month) | 18 | 263 | 56 (29, 73) | 72 (54, 83) | 11 | 622 | 28 (-32, 61) | 68 (40, 83) |
| Unvaccinated | 770 | 4950 | Reference | | 257 | 10462 | Reference | |
| **Two BNT162b2 (Pfizer-BioNTech), Hospitalization[2]** | | | | | | | | |
| 0-13 days (0-1 week) | 5 | 3219 | 97 (93, 99) | 91 (76, 96) | 6 | 9593 | 94 (87, 97) | 75 (38, 90) |
| 14-27 days (2-3 weeks) | 1 | 3253 | 99 (96, 100) | 98 (86, 100) | 3 | 10725 | 97 (92, 99) | 92 (72, 97) |
| 28-55 days (4-7 weeks, 2nd month) | 9 | 7072 | 98 (96, 99) | 96 (93, 98) | 10 | 20789 | 96 (92, 98) | 94 (88, 97) |
| 56-83 days (8-11 weeks, 3rd month) | 26 | 8904 | 95 (92, 97) | 97 (96, 98) | 21 | 21438 | 91 (86, 94) | 95 (93, 97) |
| 84-111 days (12-15 weeks, 4th month) | 37 | 8057 | 92 (89, 94) | 96 (94, 97) | 34 | 17733 | 82 (74, 88) | 94 (91, 96) |
| 112-139 days (16-19 weeks, 5th month) | 4 | 1123 | 94 (83, 98) | 96 (89, 99) | 4 | 2229 | 84 (55, 94) | 94 (84, 98) |
| 140+ days (20+ weeks, 6th+ month) | 1 | 263 | 96 (69, 99) | 98 (83, 100) | 0 | 622 | NE | NE |
| Unvaccinated | 284 | 4950 | Reference | | 114 | 10462 | Reference | |

VE=vaccine effectiveness; 95%CI=95% confidence interval; NE=Not estimable

[1] Unless otherwise specified, all VE estimates adjusted for age group (70-79, ≥80 years); sex (male, female); individual epidemiological week of the analysis period (weeks 22-39, categorical); and region of the province (5 in each province).
[2] In Quebec, adjusted VE estimates against hospitalization in ≥70-year-olds were adjusted for calendar time tri-weekly owing to sample size.

**Supplementary Table 16 Contin'd. Two-dose vaccine effectiveness against infection and hospitalization, by time since vaccination, (and mRNA vaccine type), ≥70-year-olds, British Columbia and Quebec, Canada**

| | British Columbia (BC), Canada | | | | Quebec, Canada | | | |
|---|---|---|---|---|---|---|---|---|
| | Sample sizes | | VE (95% CI) | | Sample sizes | | VE (95% CI) | |
| | Case N | Control N | Crude | Adjusted[1] | Case N | Control N | Crude | Adjusted[1] |
| **Two mRNA-1273 (Moderna), Infection** | | | | | | | | |
| 0-13 days (0-1 week) | 6 | 577 | 93 (85, 97) | 85 (66, 94) | 2 | 1996 | 96 (84, 99) | 87 (48, 97) |
| 14-27 days (2-3 weeks) | 0 | 606 | NE | NE | 2 | 2149 | 96 (85, 99) | 90 (59, 98) |
| 28-55 days (4-7 weeks, 2nd month) | 8 | 1342 | 96 (92, 98) | 96 (91, 98) | 9 | 4520 | 92 (84, 96) | 90 (80, 95) |
| 56-83 days (8-11 weeks, 3rd month) | 27 | 1630 | 89 (84, 93) | 94 (92, 96) | 26 | 4683 | 77 (66, 85) | 89 (83, 93) |
| 84-111 days (12-15 weeks, 4th month) | 39 | 1488 | 83 (77, 88) | 93 (90, 95) | 27 | 3282 | 67 (50, 78) | 85 (78, 90) |
| 112-139 days (16-19 weeks, 5th month) | 16 | 278 | 63 (38, 78) | 85 (75, 91) | 3 | 800 | 85 (52, 95) | 90 (67, 97) |
| 140+ (20+ weeks, 6th+ month) | 17 | 138 | 21 (-32, 52) | 72 (51, 84) | 2 | 569 | 86 (42, 96) | 90 (59, 98) |
| Unvaccinated | 770 | 4950 | Reference | | 257 | 10462 | Reference | |
| **Two mRNA-1273 (Moderna), Hospitalization[2]** | | | | | | | | |
| 0-13 days (0-1 week) | 1 | 577 | 97 (78, 100) | 93 (47, 99) | 0 | 1996 | NE | NE |
| 14-27 days (2-3 weeks) | 0 | 606 | NE | NE | 1 | 2149 | 96 (69, 99) | 89 (17, 98) |
| 28-55 days (4-7 weeks, 2nd month) | 0 | 1342 | NE | NE | 1 | 4520 | 98 (85, 100) | 97 (81, 100) |
| 56-83 days (8-11 weeks, 3rd month) | 3 | 1630 | 97 (90, 99) | 98 (95, 99) | 5 | 4683 | 90 (76, 96) | NE |
| 84-111 days (12-15 weeks, 4th month) | 7 | 1488 | 92 (83, 96) | 96 (92, 98) | 2 | 3282 | 94 (77, 99) | NE |
| 112-139 days (16-19 weeks, 5th month) | 6 | 278 | 62 (15, 83) | 81 (56, 92) | 1 | 800 | 89 (18, 98) | 93 (45, 99) |
| 140+ days (20+ weeks, 6th+ month) | 0 | 138 | NE | NE | 1 | 569 | 84 (-16, 98) | NE |
| Unvaccinated | 284 | 4950 | Reference | | 114 | 10462 | Reference | |

VE=vaccine effectiveness; 95%CI=95% confidence interval; NE=Not estimable

[1] Unless otherwise specified, all VE estimates adjusted for age group (70-79, ≥80 years); sex (male, female); individual epidemiological week of the analysis period (weeks 22-39, categorical); and region of the province (5 in each province).

[2] In Quebec, adjusted VE estimates against hospitalization in ≥70-year-olds were adjusted for calendar time tri-weekly owing to sample size.

**Supplementary Table 17. Two-dose vaccine effectiveness against infection and hospitalization (≥14 days post-vaccination), by interval between doses, (mRNA and ChAdOx1), ≥18-year-olds, British Columbia and Quebec, Canada**

| | British Columbia, Canada | | | | Quebec, Canada | | | |
|---|---|---|---|---|---|---|---|---|
| | Sample sizes | | VE (95% CI) | | Sample sizes | | VE (95% CI) | |
| | Case N | Control N | Crude | Adjusted[1] | Case N | Control N | Crude | Adjusted[1] |
| **Any two mRNA vaccines, Infection** | | | | | | | | |
| 21-34 days (3-4 weeks) | 211 | 4854 | 82 (80, 85) | 85 (83, 87) | 323 | 17679 | 66 (62, 69) | 79 (76, 81) |
| 35-48 days (5-6 weeks) | 662 | 15844 | 83 (82, 84) | 85 (83, 86) | 419 | 30195 | 74 (71, 76) | 85 (84, 87) |
| 49-62 days (7-8 weeks) | 2509 | 86580 | 88 (88, 89) | 91 (91, 91) | 1361 | 124758 | 80 (78, 81) | 89 (88, 89) |
| 63-83 days (9-11 weeks) | 2049 | 81384 | 90 (89, 90) | 91 (91, 91) | 1355 | 159120 | 84 (83, 85) | 89 (89, 90) |
| 84-111 days (12-15 weeks) | 1038 | 36602 | 88 (88, 89) | 88 (88, 89) | 735 | 109238 | 87 (86, 88) | 89 (88, 90) |
| 112+ days (16+ weeks) | 126 | 4924 | 90 (88, 91) | 91 (89, 92) | 441 | 71263 | 88 (87, 89) | 90 (89, 91) |
| 49+ days (7+ weeks) | 5722 | 209490 | 89 (89, 89) | 91 (90, 91) | 3892 | 464379 | 84 (84, 85) | 89 (89, 90) |
| Unvaccinated | 18809 | 76621 | Reference | | 11366 | 212918 | Reference | |
| **Any two mRNA vaccines, Hospitalization** | | | | | | | | |
| 21-34 days (3-4 weeks) | 9 | 4854 | 90 (80, 95) | 93 (87, 96) | 15 | 17679 | 74 (57, 85) | 87 (79, 92) |
| 35-48 days (5-6 weeks) | 11 | 15844 | 96 (93, 98) | 97 (94, 98) | 5 | 30195 | 95 (88, 98) | 97 (93, 99) |
| 49-62 days (7-8 weeks) | 24 | 86580 | 98 (98, 99) | 99 (98, 99) | 12 | 124758 | 97 (95, 98) | 98 (97, 99) |
| 63-83 days (9-11 weeks) | 59 | 81384 | 96 (95, 97) | 98 (98, 99) | 38 | 159120 | 93 (90, 95) | 97 (96, 98) |
| 84-111 days (12-15 weeks) | 66 | 36602 | 90 (87, 92) | 95 (94, 96) | 43 | 109238 | 88 (84, 91) | 96 (95, 97) |
| 112+ days (16+ weeks) | 9 | 4924 | 90 (80, 95) | 94 (89, 97) | 30 | 71263 | 87 (82, 91) | 95 (93, 97) |
| 49+ days (7+ weeks) | 158 | 209490 | 96 (95, 96) | 98 (97, 98) | 123 | 464379 | 92 (90, 93) | 97 (96, 97) |
| Unvaccinated | 1365 | 76621 | Reference | | 702 | 212918 | Reference | |
| **Two ChAdOx1 vaccines, Infection** | | | | | | | | |
| 21-34 days (3-4 weeks) | 7 | 46 | 38 (-37, 72) | 47 (-19, 76) | 6 | 165 | 32 (-54, 70) | 47 (-22, 77) |
| 35-48 days (5-6 weeks) | 25 | 293 | 65 (48, 77) | 73 (60, 82) | 19 | 734 | 52 (24, 69) | 61 (39, 76) |
| 49-62 days (7-8 weeks) | 307 | 4170 | 70 (66, 73) | 74 (70, 77) | 82 | 4390 | 65 (56, 72) | 69 (62, 76) |
| 63-83 days (9-11 weeks) | 261 | 3139 | 66 (62, 70) | 69 (65, 73) | 101 | 9174 | 79 (75, 83) | 76 (70, 80) |
| 84+ days (12+ weeks) | 45 | 429 | 57 (42, 69) | 62 (48, 72) | 22 | 2439 | 83 (74, 89) | 82 (73, 88) |
| 49+ days (7+ weeks) | 613 | 7738 | 68 (65, 70) | 71 (69, 74) | 205 | 16003 | 76 (72, 79) | 75 (71, 78) |
| Unvaccinated | 18809 | 76621 | Reference | | 11366 | 212918 | Reference | |
| **Two ChAdOx1 vaccines, Hospitalization** | | | | | | | | |
| 21-34 days (3-4 weeks) | 0 | 46 | NE | NE | 1 | 165 | NE | NE |
| 35-48 days (5-6 weeks) | 0 | 293 | NE | NE | 0 | 734 | NE | NE |
| 49-62 days (7-8 weeks) | 10 | 4170 | 87 (75, 93) | 93 (86, 96) | 2 | 4390 | 86 (45, 97) | 96 (83, 99) |
| 63-83 days (9-11 weeks) | 6 | 3139 | 89 (76, 95) | 95 (89, 98) | 7 | 9174 | 77 (51, 89) | 94 (88, 97) |
| 84+ days (12+ weeks) | 1 | 429 | 87 (7, 98) | 91 (34, 99) | 3 | 2439 | 63 (-16, 88) | 91 (73, 97) |
| 49+ days (7+ weeks) | 17 | 7738 | 88 (80, 92) | 94 (90, 96) | 12 | 16003 | 77 (60, 87) | 94 (90, 97) |
| Unvaccinated | 1365 | 76621 | Reference | | 702 | 212918 | Reference | |

VE=vaccine effectiveness; 95%CI=95% confidence interval

[1] Unless otherwise specified, all VE estimates adjusted for age group (18-49, 50-69, 70-79, 80+ years); sex (male, female); individual epidemiological week (22-39); and region of the province.

**Supplementary Table 18. Two-dose vaccine effectiveness against infection and hospitalization (≥14 days post-vaccination), by interval between doses, (and mRNA vaccine type), ≥18-year-olds, British Columbia and Quebec, Canada**

| | British Columbia, Canada | | | | Quebec, Canada | | | |
|---|---|---|---|---|---|---|---|---|
| | Sample sizes | | VE (95% CI) | | Sample sizes | | VE (95% CI) | |
| | Case N | Control N | Crude | Adjusted[1] | Case N | Control N | Crude | Adjusted[1] |
| **Two BNT162b2 (Pfizer-BioNTech) vaccines, Infection** | | | | | | | | |
| 21-34 days (3-4 weeks) | 133 | 3094 | 82 (79, 85) | 85 (82, 87) | 233 | 10384 | 58 (52, 63) | 74 (71, 78) |
| 35-48 days (5-6 weeks) | 450 | 11113 | 84 (82, 85) | 84 (83, 86) | 266 | 17651 | 72 (68, 75) | 85 (83, 86) |
| 49-62 days (7-8 weeks) | 1907 | 62727 | 88 (87, 88) | 91 (90, 91) | 1080 | 90646 | 78 (76, 79) | 88 (87, 89) |
| 63-83 days (9-11 weeks) | 1527 | 59605 | 90 (89, 90) | 91 (90, 91) | 1099 | 119473 | 83 (82, 84) | 89 (88, 89) |
| 84-111 days (12-15 weeks) | 762 | 28435 | 89 (88, 90) | 89 (88, 90) | 622 | 86829 | 87 (85, 88) | 89 (88, 89) |
| 112+ days (16+ weeks) | 97 | 3857 | 90 (87, 92) | 91 (88, 92) | 407 | 62985 | 88 (87, 89) | 90 (89, 91) |
| 49+ days (7+ weeks) | 4293 | 154624 | 89 (88, 89) | 91 (90, 91) | 3208 | 359933 | 83 (83, 84) | 89 (88, 89) |
| Unvaccinated | 18809 | 76621 | Reference | | 11366 | 212918 | Reference | |
| **Two BNT162b2 (Pfizer-BioNTech) vaccines, Hospitalization** | | | | | | | | |
| 21-34 days (3-4 weeks) | 4 | 3094 | 93 (81, 97) | 95 (86, 98) | 6 | 10384 | 82 (61, 92) | 92 (82, 96) |
| 35-48 days (5-6 weeks) | 4 | 11113 | 98 (95, 99) | 98 (95, 99) | 4 | 17651 | 93 (82, 97) | 96 (89, 99) |
| 49-62 days (7-8 weeks) | 18 | 62727 | 98 (97, 99) | 99 (98, 99) | 9 | 90646 | 97 (94, 98) | 98 (97, 99) |
| 63-83 days (9-11 weeks) | 42 | 59605 | 96 (95, 97) | 98 (98, 99) | 34 | 119473 | 91 (88, 94) | 97 (95, 98) |
| 84-111 days (12-15 weeks) | 47 | 28435 | 91 (88, 93) | 95 (93, 96) | 36 | 86829 | 87 (82, 91) | 96 (94, 97) |
| 112+ days (16+ weeks) | 7 | 3857 | 90 (79, 95) | 94 (88, 97) | 24 | 62985 | 88 (83, 92) | 96 (93, 97) |
| 49+ days (7+ weeks) | 114 | 154624 | 96 (95, 97) | 98 (97, 98) | 103 | 359933 | 91 (89, 93) | 97 (96, 97) |
| Unvaccinated | 1365 | 76621 | Reference | | 702 | 212918 | Reference | |
| **Two mRNA-1273 (Moderna) vaccines, Infection** | | | | | | | | |
| 21-34 days (3-4 weeks) | 65 | 1565 | 83 (78, 87) | 87 (84, 90) | 75 | 6531 | 78 (73, 83) | 86 (82, 89) |
| 35-48 days (5-6 weeks) | 182 | 3935 | 81 (78, 84) | 85 (82, 87) | 125 | 10376 | 77 (73, 81) | 87 (84, 89) |
| 49-62 days (7-8 weeks) | 384 | 16484 | 91 (89, 91) | 92 (92, 93) | 245 | 29984 | 85 (83, 87) | 91 (90, 92) |
| 63-83 days (9-11 weeks) | 339 | 14137 | 90 (89, 91) | 92 (91, 93) | 227 | 35943 | 88 (87, 90) | 91 (90, 92) |
| 84-111 days (12-15 weeks) | 218 | 5047 | 82 (80, 85) | 86 (84, 88) | 98 | 19989 | 91 (89, 92) | 90 (88, 92) |
| 112+ days (16+ weeks) | 21 | 721 | 88 (82, 92) | 91 (86, 94) | 26 | 7038 | 93 (90, 95) | 92 (89, 95) |
| 49+ days (7+ weeks) | 962 | 36389 | 89 (88, 90) | 91 (91, 92) | 596 | 92954 | 88 (87, 89) | 91 (90, 92) |
| Unvaccinated | 18809 | 76621 | Reference | | 11366 | 212918 | Reference | |
| **Two mRNA-1273 (Moderna) vaccines, Hospitalization** | | | | | | | | |
| 21-34 days (3-4 weeks) | 4 | 1565 | 86 (62, 95) | 91 (77, 97) | 7 | 6531 | 67 (32, 85) | 82 (63, 92) |
| 35-48 days (5-6 weeks) | 7 | 3935 | 90 (79, 95) | 93 (85, 97) | 1 | 10376 | 97 (79, 100) | 98 (87, 100) |
| 49-62 days (7-8 weeks) | 2 | 16484 | 99 (97, 100) | 100 (98, 100) | 2 | 29984 | 98 (92, 99) | 99 (95, 100) |
| 63-83 days (9-11 weeks) | 10 | 14137 | 96 (93, 98) | 98 (97, 99) | 4 | 35943 | 97 (91, 99) | 98 (96, 99) |
| 84-111 days (12-15 weeks) | 14 | 5047 | 84 (74, 91) | 94 (90, 96) | 6 | 19989 | 91 (80, 96) | 96 (92, 98) |
| 112+ days (16+ weeks) | 1 | 721 | 92 (45, 99) | 97 (76, 100) | 4 | 7038 | 83 (54, 94) | 95 (86, 98) |
| 49+ days (7+ weeks) | 27 | 36389 | 96 (94, 97) | 98 (97, 98) | 16 | 92954 | 95 (91, 97) | 98 (96, 99) |
| Unvaccinated | 1365 | 76621 | Reference | | 702 | 212918 | Reference | |

VE=vaccine effectiveness; 95%CI=95% confidence interval

[1] Unless otherwise specified, all VE estimates adjusted for age group (18-49, 50-69, 70-79, 80+ years); sex (male, female); individual epidemiological week (22-39); and region of the province.

**Supplementary Table 19. Two-dose vaccine effectiveness against infection, by interval between doses and time since second dose, by vaccine type, ≥18-year-olds, British Columbia and Quebec, Canada**

| | British Columbia, Canada | | | | Quebec, Canada | | | |
|---|---|---|---|---|---|---|---|---|
| | Sample sizes | | VE (95% CI) | | Sample sizes | | VE (95% CI) | |
| | Case N | Control N | Crude | Adjusted[1] | Case N | Control N | Crude | Adjusted[1] |
| **Two mRNA vaccines** | | | | | | | | |
| **14-27 days (2-3 weeks) since second dose** | | | | | | | | |
| 21-34 day (3-4 week) interval between doses | 28 | 902 | 87 (82, 91) | 90 (85, 93) | 45 | 4099 | 79 (72, 85) | 87 (82, 90) |
| 35-48 day (5-6 week) interval between doses | 30 | 1391 | 91 (87, 94) | 93 (90, 95) | 27 | 4675 | 89 (84, 93) | 89 (84, 92) |
| 49-62 day (7+ week) interval between doses | 320 | 25855 | 95 (94, 95) | 94 (94, 95) | 252 | 63765 | 93 (92, 93) | 92 (91, 93) |
| **28-55 days (4-7 weeks) since second dose** | | | | | | | | |
| 21-34 day (3-4 week) interval between doses | 35 | 1068 | 87 (81, 90) | 89 (84, 92) | 95 | 5166 | 65 (58, 72) | 78 (74, 82) |
| 35-48 day (5-6 week) interval between doses | 67 | 2232 | 88 (84, 90) | 91 (88, 93) | 133 | 9720 | 74 (70, 78) | 85 (82, 88) |
| 49-62 day (7+ week) interval between doses | 1512 | 66747 | 91 (90, 91) | 92 (92, 93) | 1006 | 147185 | 87 (86, 88) | 91 (90, 91) |
| **56-83 days (8-11 weeks) since second dose** | | | | | | | | |
| 21-34 day (3-4 week) interval between doses | 30 | 698 | 82 (75, 88) | 85 (78, 90) | 81 | 3329 | 54 (43, 63) | 74 (68, 79) |
| 35-48 day (5-6 week) interval between doses | 58 | 1421 | 83 (78, 87) | 89 (85, 91) | 179 | 10893 | 69 (64, 73) | 86 (84, 88) |
| 49-62 day (7+ week) interval between doses | 2173 | 72028 | 88 (87, 88) | 90 (90, 91) | 1483 | 142410 | 80 (79, 82) | 88 (88, 89) |
| **84-111 days (12-15 weeks) since second dose** | | | | | | | | |
| 21-34 day (3-4 week) interval between doses | 36 | 719 | 80 (71, 85) | 84 (77, 88) | 62 | 2221 | 48 (33, 59) | 70 (62, 77) |
| 35-48 day (5-6 week) interval between doses | 48 | 1393 | 86 (81, 89) | 85 (81, 89) | 55 | 3425 | 70 (61, 77) | 85 (80, 88) |
| 49-62 day (7+ week) interval between doses | 1344 | 36881 | 85 (84, 86) | 88 (87, 89) | 824 | 74484 | 79 (78, 81) | 86 (85, 87) |
| **112+ days (16+ weeks) since second dose** | | | | | | | | |
| 21-34 day (3-4 week) interval between doses | 82 | 1467 | 77 (72, 82) | 81 (76, 85) | 40 | 2864 | 74 (64, 81) | 81 (74, 86) |
| 35-48 day (5-6 week) interval between doses | 459 | 9407 | 80 (78, 82) | 80 (78, 82) | 25 | 1482 | 68 (53, 79) | 78 (68, 85) |
| 49-62 day (7+ week) interval between doses | 373 | 7979 | 81 (79, 83) | 84 (82, 86) | 327 | 36535 | 83 (81, 85) | 90 (88, 91) |
| Unvaccinated | 18809 | 76621 | Reference | | 11366 | 212918 | Reference | |
| **Two BNT162b2 (Pfizer-BioNTech, Comirnaty)** | | | | | | | | |
| **14-27 days (2-3 weeks) since second dose** | | | | | | | | |
| 21-34 day (3-4 week) interval between doses | 14 | 452 | 87 (79, 93) | 89 (82, 94) | 35 | 2532 | 74 (64, 81) | 83 (77, 88) |
| 35-48 day (5-6 week) interval between doses | 18 | 631 | 88 (81, 93) | 90 (84, 94) | 18 | 2799 | 88 (81, 92) | 88 (80, 92) |
| 49-62 day (7+ week) interval between doses | 270 | 19626 | 94 (94, 95) | 94 (93, 94) | 203 | 49779 | 92 (91, 93) | 92 (91, 93) |
| **28-55 days (4-7 weeks) since second dose** | | | | | | | | |
| 21-34 day (3-4 week) interval between doses | 22 | 575 | 84 (76, 90) | 86 (78, 91) | 66 | 2870 | 57 (45, 66) | 73 (66, 79) |
| 35-48 day (5-6 week) interval between doses | 39 | 1019 | 84 (79, 89) | 88 (83, 91) | 91 | 5873 | 71 (64, 76) | 84 (80, 87) |
| 49-62 day (7+ week) interval between doses | 1207 | 50335 | 90 (90, 91) | 92 (91, 92) | 832 | 115408 | 86 (86, 87) | 90 (89, 91) |
| **56-83 days (8-11 weeks) since second dose** | | | | | | | | |
| 21-34 day (3-4 week) interval between doses | 21 | 488 | 82 (73, 89) | 85 (76, 90) | 56 | 2007 | 48 (32, 60) | 72 (63, 78) |
| 35-48 day (5-6 week) interval between doses | 35 | 819 | 83 (76, 88) | 88 (83, 92) | 108 | 6588 | 69 (63, 75) | 86 (83, 89) |
| 49-62 day (7+ week) interval between doses | 1611 | 52300 | 87 (87, 88) | 90 (89, 91) | 1194 | 108152 | 79 (78, 81) | 88 (87, 88) |
| **84-111 days (12-15 weeks) since second dose** | | | | | | | | |
| 21-34 day (3-4 week) interval between doses | 24 | 512 | 81 (71, 87) | 85 (77, 90) | 48 | 1285 | 30 (7, 48) | 63 (50, 72) |
| 35-48 day (5-6 week) interval between doses | 29 | 956 | 88 (82, 91) | 86 (79, 90) | 35 | 1818 | 64 (50, 74) | 82 (75, 87) |
| 49-62 day (7+ week) interval between doses | 956 | 26161 | 85 (84, 86) | 88 (87, 89) | 684 | 57265 | 78 (76, 79) | 85 (84, 86) |
| **112+ days (16+ weeks) since second dose** | | | | | | | | |
| 21-34 day (3-4 week) interval between doses | 52 | 1067 | 80 (74, 85) | 82 (76, 87) | 28 | 1690 | 69 (55, 79) | 78 (68, 85) |
| 35-48 day (5-6 week) interval between doses | 329 | 7688 | 83 (81, 84) | 82 (80, 84) | 14 | 573 | 54 (22, 73) | 73 (54, 84) |
| 49-62 day (7+ week) interval between doses | 249 | 6202 | 84 (81, 86) | 85 (83, 87) | 295 | 29329 | 81 (79, 83) | 90 (88, 91) |
| Unvaccinated | 18809 | 76621 | Reference | | 11366 | 212918 | Reference | |

VE=vaccine effectiveness; 95%CI=95% confidence interval

[1] Unless otherwise specified, all VE estimates adjusted for age group (18-49, 50-69, 70-79, 80+ years); sex (male, female); individual epidemiological week (22-39); and region of the province.

**Supplementary Table 19 Cont'd. Two-dose vaccine effectiveness against infection, by interval between doses and time since second dose, by vaccine type, ≥18-year-olds, British Columbia and Quebec, Canada**

| | British Columbia, Canada | | | | Quebec, Canada | | | |
|---|---|---|---|---|---|---|---|---|
| | Sample sizes | | VE (95% CI) | | Sample sizes | | VE (95% CI) | |
| | Case N | Control N | Crude | Adjusted[1] | Case N | Control N | Crude | Adjusted[1] |
| **Two mRNA-1273 (Moderna, Spikevax)** | | | | | | | | |
| **14-27 days (2-3 weeks) since second dose** | | | | | | | | |
| 21-34 day (3-4 week) interval between doses | 13 | 413 | 87 (78, 93) | 91 (84, 95) | 8 | 1430 | 90 (79, 95) | 93 (87, 97) |
| 35-48 day (5-6 week) interval between doses | 11 | 660 | 93 (88, 96) | 95 (91, 97) | 9 | 1546 | 89 (79, 94) | 90 (80, 95) |
| 49-62 day (7+ week) interval between doses | 41 | 4439 | 96 (95, 97) | 96 (94, 97) | 46 | 12339 | 93 (91, 95) | 92 (90, 94) |
| **28-55 days (4-7 weeks) since second dose** | | | | | | | | |
| 21-34 day (3-4 week) interval between doses | 11 | 440 | 90 (81, 94) | 92 (86, 96) | 25 | 2034 | 77 (66, 84) | 86 (79, 90) |
| 35-48 day (5-6 week) interval between doses | 21 | 980 | 91 (87, 94) | 93 (90, 96) | 34 | 3167 | 80 (72, 86) | 88 (83, 91) |
| 49-62 day (7+ week) interval between doses | 216 | 11632 | 92 (91, 93) | 94 (93, 94) | 153 | 27986 | 90 (88, 91) | 92 (91, 94) |
| **56-83 days (8-11 weeks) since second dose** | | | | | | | | |
| 21-34 day (3-4 week) interval between doses | 3 | 149 | 92 (74, 97) | 92 (76, 98) | 19 | 1110 | 68 (49, 80) | 79 (67, 87) |
| 35-48 day (5-6 week) interval between doses | 7 | 305 | 91 (80, 96) | 93 (86, 97) | 52 | 3423 | 72 (63, 78) | 86 (82, 90) |
| 49-62 day (7+ week) interval between doses | 346 | 12489 | 89 (87, 90) | 91 (90, 92) | 241 | 30329 | 85 (83, 87) | 91 (90, 92) |
| **84-111 days (12-15 weeks) since second dose** | | | | | | | | |
| 21-34 day (3-4 week) interval between doses | 8 | 167 | 80 (60, 90) | 84 (68, 92) | 11 | 833 | 75 (55, 86) | 84 (72, 91) |
| 35-48 day (5-6 week) interval between doses | 13 | 285 | 81 (68, 89) | 81 (67, 89) | 19 | 1393 | 74 (60, 84) | 86 (78, 91) |
| 49-62 day (7+ week) interval between doses | 240 | 6233 | 84 (82, 86) | 89 (87, 90) | 130 | 15689 | 84 (82, 87) | 88 (85, 90) |
| **112+ days (16+ weeks) since second dose** | | | | | | | | |
| 21-34 day (3-4 week) interval between doses | 30 | 396 | 69 (55, 79) | 78 (68, 85) | 12 | 1124 | 80 (65, 89) | 85 (73, 92) |
| 35-48 day (5-6 week) interval between doses | 130 | 1705 | 69 (63, 74) | 74 (69, 78) | 11 | 847 | 76 (56, 87) | 81 (66, 90) |
| 49-62 day (7+ week) interval between doses | 119 | 1596 | 70 (63, 75) | 81 (77, 84) | 26 | 6611 | 93 (89, 95) | 91 (87, 94) |
| Unvaccinated | 18809 | 76621 | Reference | | 11366 | 212918 | Reference | |
| **Two ChAdOx1 (AstraZeneca, Vaxzevria/COVISHIELD)** | | | | | | | | |
| **14-27 days (2-3 weeks) since second dose** | | | | | | | | |
| 21-34 day (3-4 week) interval between doses | 0 | 10 | NE | NE | 17 | 103 | NE | NE |
| 35-48 day (5-6 week) interval between doses | 1 | 16 | NE | NE | 4 | 2065 | NE | NE |
| 49-62 day (7+ week) interval between doses | 7 | 522 | 95 (88, 97) | 80 (57, 90) | 26 | 2 | 96 (90, 99) | 85 (60, 94) |
| **28-55 days (4-7 weeks) since second dose** | | | | | | | | |
| 21-34 day (3-4 week) interval between doses | 2 | 8 | NE | NE | 139 | 15 | NE | NE |
| 35-48 day (5-6 week) interval between doses | 7 | 59 | 51 (-6, 78) | 51 (-8, 78) | 3805 | 43 | NE | NE |
| 49-62 day (7+ week) interval between doses | 68 | 1437 | 81 (75, 85) | 75 (68, 81) | 2 | 242 | 93 (88, 96) | 84 (73, 90) |
| **56-83 days (8-11 weeks) since second dose** | | | | | | | | |
| 21-34 day (3-4 week) interval between doses | 3 | 13 | NE | NE | 4119 | 37 | NE | NE |
| 35-48 day (5-6 week) interval between doses | 9 | 92 | 60 (21, 80) | 75 (51, 88) | 9 | 215 | 52 (-29, 82) | 58 (-14, 85) |
| 49-62 day (7+ week) interval between doses | 236 | 2721 | 65 (60, 69) | 73 (69, 77) | 103 | 4620 | 74 (66, 80) | 75 (68, 81) |
| **84-111 days (12-15 weeks) since second dose** | | | | | | | | |
| 21-34 day (3-4 week) interval between doses | 2 | 5 | NE | NE | 6 | 48 | NE | NE |
| 35-48 day (5-6 week) interval between doses | 6 | 104 | 76 (46, 90) | 83 (61, 92) | 4 | 120 | 22 (-53, 60) | 65 (31, 82) |
| 49-62 day (7+ week) interval between doses | 264 | 2839 | 62 (57, 67) | 70 (65, 73) | 25 | 1394 | 58 (49, 66) | 72 (66, 77) |
| **84+ days (12+ weeks) since second dose** | | | | | | | | |
| 21-34 day (3-4 week) interval between doses | 2 | 15 | NE | NE | 6 | 85 | NE | NE |
| 35-48 day (5-6 week) interval between doses | 8 | 126 | 74 (47, 87) | 81 (61, 91) | 13 | 335 | 27 (-27, 58) | 64 (37, 79) |
| 49-62 day (7+ week) interval between doses | 302 | 3058 | 60 (55, 64) | 68 (64, 72) | 128 | 6014 | 60 (52, 67) | 72 (66, 77) |
| Unvaccinated | 18809 | 76621 | Reference | | 11366 | 212918 | Reference | |

VE=vaccine effectiveness; 95%CI=95% confidence interval; NE=Not estimable

[1] Unless otherwise specified, all VE estimates adjusted for age group (18-49, 50-69, 70-79, 80+ years); sex (male, female); individual epidemiological week (22-39); and region of the province.
